# Baseline drug treatments and long-term outcomes in COVID-19-hospitalized patients: results of the 2020 AUTCOV study

**DOI:** 10.1101/2024.08.22.24312424

**Authors:** Alexandra Christine Graf, Berthold Reichardt, Christine Wagenlechner, Pavla Krotka, Denise Traxler-Weidenauer, Michael Mildner, Julia Mascherbauer, Clemens Aigner, Johann Auer, Ralph Wendt, Hendrik Jan Ankersmit

**Author notes:** contributed equally (Alexandra Christine Graf, Ralph Wendt, Hendrik Jan Ankersmit).

## Abstract

Limited data are available on long-term morbidity and mortality after COVID-19 hospitalization. In this population-based study, we investigated the long-term mortality and morbidity after COVID-19 hospitalization and associations with baseline drug treatments. Data were provided on hospitalized COVID-19 patients in 2020 and matched controls by the Austrian Health Insurance Funds. The primary outcome was all-cause mortality. Secondary outcomes were all-cause mortality conditional on COVID-hospital survival and re-hospitalization due to any reason. The median follow-up was 600 days. 22 571 patients aged >18 years were hospitalized in Austria in 2020 due to COVID-19. The risk of all-cause mortality was significantly higher with polypharmacy. With the exception of the youngest age group (19-40 years), antiepileptics, antipsychotics and the medicament group of iron supplements, erythropoietic stimulating agents, Vitamin B12, and folic acid were significantly associated with a higher risk of death (all p<0,001). For Non-steroidal anti-inflammatory drugs and other anti-inflammatory drugs, significantly increased survival was observed (all p<0,001). Patients had a higher drug prescription load than the control population. Long-term mortality and the risk of re-hospitalization due to any reason were also significantly greater in the patients. Antipsychotics are assumed to be an underrecognized medication group linked to worse outcomes after COVID-19 hospitalization.

## Introduction

The outbreak of Coronavirus Disease 2019 (COVID-19) marked the start of a global pandemic and has been associated with substantial morbidity and mortality.

Age and gender are well-established risk factors for severe outcomes.^1-5^ Numerous reports have discussed that the presence of comorbidities may increase the risk of COVID-19-related death.^1,6-9^ In addition, an increased risk of readmission and mortality after hospital discharge has been observed up to 1 year after COVID-19 hospitalization.^10-13^ Patients who require hospitalization for COVID-19 have a greater comorbidity burden and are expected to have worse short-term outcomes.^12^ Differences in the prevalence of drug use and polypharmacy regimens were observed when compared to the general population.^14^ Studies have investigated possible associations of polypharmacy with increased morbidity and mortality among patients with COVID-19.^15,16^ Visser et al.^17^ investigated the impact of polypharmacy on COVID-19-related mortality in nursing home residents and found a significant positive association between the total number of medications and 30-day COVID-related adjusted mortality. However, published studies focusing on the drug profiles of hospitalized COVID-19 patients and long-term outcomes are still scarce.

Here, we present data on COVID-19-hospitalized patients and a matched control group provided by the Austrian Health Insurance Funds with a median follow-up of 600 days (maximum 880 days). The primary aim of this study was to analyze the long-term follow-up of patients hospitalized due to COVID-19 in Austria in 2020 in order to evaluate the association between prescribed medications and mortality or readmission after COVID-19 hospitalization. We also compared the characteristics and outcomes of the hospitalized patients with an age-, sex-, and region-matched control group, presenting a real-world picture.

## Methods

### Study Design and Cohorts

This retrospective, national population-based study complied with the Declaration of Helsinki and was approved by the ethics committee of Lower Austria (GS1-EK-4/747-2021). The data on both the patient and control cohorts were available from the Austrian Health Insurance Funds. Approximately 98% of the Austrian population is registered in the public health insurance system. Health care in Austria is a national system with good access to care, is regulated by the social insurance law and mainly financed by social insurance contributions.

Patients >18 years of age hospitalized in Austria due to the main diagnosis COVID-19 (ICD-10 Codes U071, U072, U049) from 1 January 2020 to 31 December 2020 were included in this study. For all patients age, sex, region, and medication were obtained from 1 year before hospitalization until study cut-off.

An age-, sex-, and region-matched control group (approximately 10 controls for each patient) consisted of individuals not hospitalized due to COVID-19 in the year 2020 were randomly chosen from the population registered in the Austrian Health Insurance Fund, representing the Austrian population. Data on the control group were available from 1 year before the first patient was hospitalized until study cut-off. The data set includes 22 571 patients and 217 295 controls.

### Study Outcomes

The primary outcome was all-cause mortality. The secondary outcome was hospitalization due to any reason. Hospitalization was defined based on billing information (MEL codes). For patients, we used the first readmission after the index COVID-19 hospital stay. For controls, hospitalization was defined as the first hospital admission after the index COVID hospital stay of the matched patient (Table S1). For controls, time to death was evaluated from the index COVID-hospital admission date of the matched patient.

### Statistical Analysis

For each patient, age, sex, region, and Anatomical Therapeutic Chemical Classification-Codes (ATC) describing prescribed medications were (for details on the statistical analyses plan see supplement). All analyses were performed separately for four age subgroups: 19-40 years, 41-64 years, 65-74 years, ≥75 years (Table S2). ATC codes before hospitalization were summarized in medication groups (Table S3). A binary variable was defined for each medication group, which was set to 1 if a drug of the corresponding ICD10-codes was prescribed at least once 1 year before the index COVID-19 hospitalization and 0 if no drug was prescribed. Twenty medication groups were used for statistical modeling (MG1 to MG20, see Table S3). For controls, a similar medication profile was generated using the drugs prescribed 1 year before the index COVID-hospital stay of the matched patient. These 20 medication groups were also used to define polypharmacy.

Numbers and percentages were used to summarize categorical variables, medians and interquartile ranges for continuous variables.

To evaluate the association between polypharmacy and all-cause death, a simple Cox regression model was calculated accounting for sex, age, half-year, and polypharmacy with clustering variable region. Polypharmacy was defined as the number of medication groups in which a patient received prescribed medication, categorized into four groups (0-1, 2-5, 6-10, >11). To evaluate the association between polypharmacy and re-hospitalization due to any reason, competing risk models (with competing risk death) were calculated accounting for sex, age, half-year, and polypharmacy with clustering variable region. Furthermore, the association between several medication groups and all-cause death, a Cox regression model was calculated accounting for sex, age, half-year, polypharmacy, and the 20 medication groups with clustering variable region. For re-hospitalization, a competing risk model was calculated using the same co-variables as described in the model for all-cause death.

Only 20 of the 32 medication groups were evaluated for the statistical models due to the underrepresentation.

As patients hospitalized due to COVID-19 had potentially more serious co-morbidities compared the Austrian population, we attempted to account for this imbalance using propensity score matching for age, sex, region, and medication groups MG1 to MG20. To evaluate the difference between patients and controls in the risk of all-cause death, a Cox regression model was calculated accounting for group, sex, age, half-year, polypharmacy, and the 20 medication groups with clustering variable region. For hospitalization due to any reason, a competing risk model was calculated using the same co-variables as described in the model for all-cause death.

Schönfeld residuals were used to evaluate the proportional hazard assumption and variance inflation factors to evaluate multicollinearity. For all models, hazard ratios (HR) and confidence intervals (CI) are provided. Due to the large sample size and large number of investigated co-variables (i.e., 25), p<0,002 (=0,05/25 applying conservative Bonferroni correction) was considered significant. All analyses were performed using R, release 4.2.2.^18^

## Results

### Characteristics of the Patients and Controls

The study population included 22 571 COVID-19-hospitalized patients (Table 1, Figure 1A). The median follow-up time varied from 594 to 615 days over the four age groups. The age-, sex-, and region-matched control group, representing the general Austrian population, included 217 295 controls (Table 1).

**Table 1:**
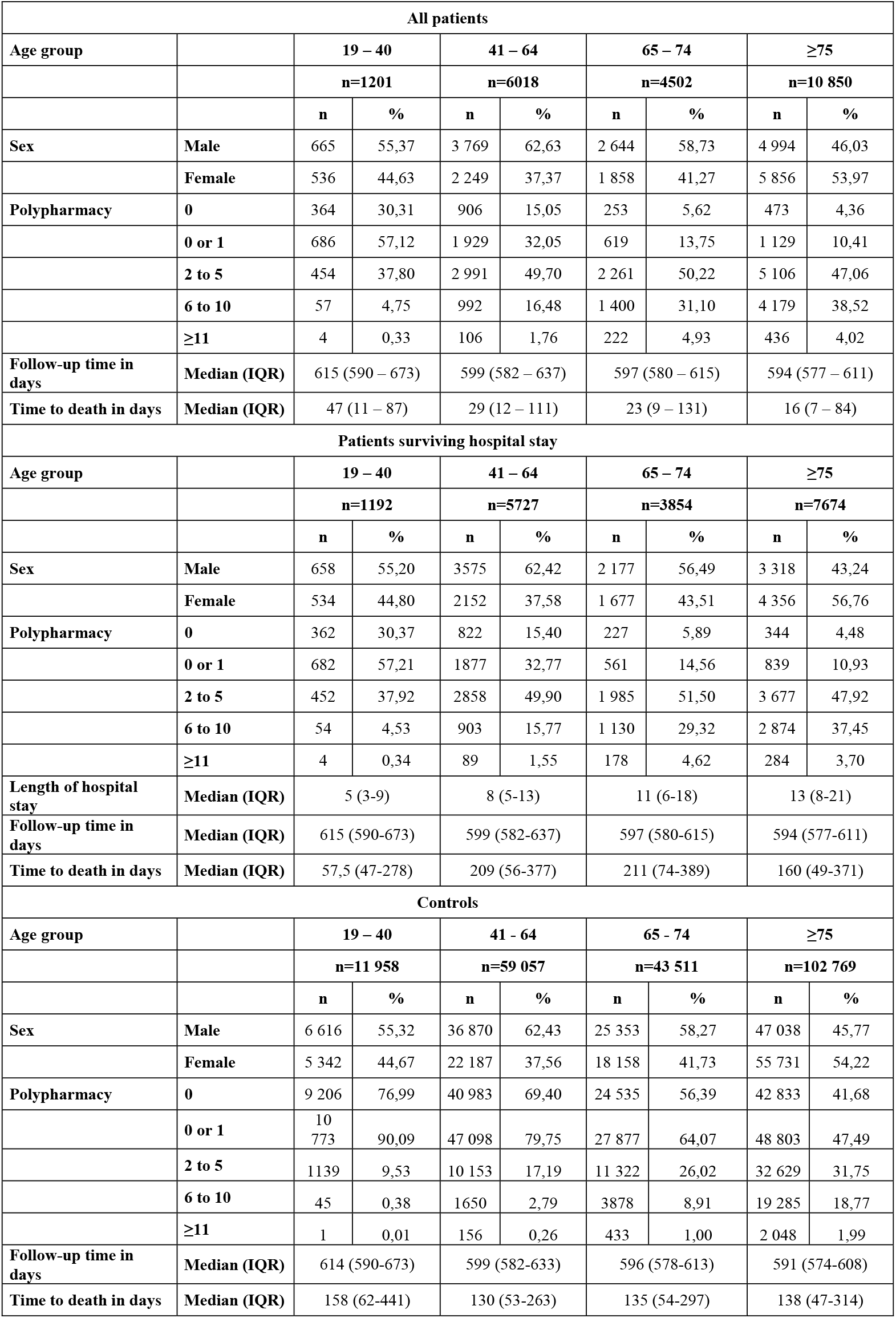
Demographic data for COVID-19-hospitalized patients in 2020 in Austria and corresponding age-, sex-, and region-matched controls.

| All patients |  |  |  |  |  |  |  |  |  |
| --- | --- | --- | --- | --- | --- | --- | --- | --- | --- |
| Age group |  | 19 – 40 |  | 41 – 64 |  | 65 – 74 |  | ≥75 |  |
|  |  | n=1201 |  | n=6018 |  | n=4502 |  | n=10 850 |  |
|  |  | n | % | n | % | n | % | n | % |
| Sex | Male | 665 | 55,37 | 3 769 | 62,63 | 2 644 | 58,73 | 4 994 | 46,03 |
|  | Female | 536 | 44,63 | 2 249 | 37,37 | 1 858 | 41,27 | 5 856 | 53,97 |
| Polypharmacy | 0 | 364 | 30,31 | 906 | 15,05 | 253 | 5,62 | 473 | 4,36 |
|  | 0 or 1 | 686 | 57,12 | 1 929 | 32,05 | 619 | 13,75 | 1 129 | 10,41 |
|  | 2 to 5 | 454 | 37,80 | 2 991 | 49,70 | 2 261 | 50,22 | 5 106 | 47,06 |
|  | 6 to 10 | 57 | 4,75 | 992 | 16,48 | 1 400 | 31,10 | 4 179 | 38,52 |
|  | ≥11 | 4 | 0,33 | 106 | 1,76 | 222 | 4,93 | 436 | 4,02 |
| Follow-up time in days | Median (IQR) | 615 (590 – 673) |  | 599 (582 – 637) |  | 597 (580 – 615) |  | 594 (577 – 611) |  |
| Time to death in days | Median (IQR) | 47 (11 – 87) |  | 29 (12 – 111) |  | 23 (9 – 131) |  | 16 (7 – 84) |  |
| Patients surviving hospital stay |  |  |  |  |  |  |  |  |  |
| Age group |  | 19 – 40 |  | 41 – 64 |  | 65 – 74 |  | ≥75 |  |
|  |  | n=1192 |  | n=5727 |  | n=3854 |  | n=7674 |  |
|  |  | n | % | n | % | n | % | n | % |
| Sex | Male | 658 | 55,20 | 3575 | 62,42 | 2 177 | 56,49 | 3 318 | 43,24 |
|  | Female | 534 | 44,80 | 2152 | 37,58 | 1 677 | 43,51 | 4 356 | 56,76 |
| Polypharmacy | 0 | 362 | 30,37 | 822 | 15,40 | 227 | 5,89 | 344 | 4,48 |
|  | 0 or 1 | 682 | 57,21 | 1877 | 32,77 | 561 | 14,56 | 839 | 10,93 |
|  | 2 to 5 | 452 | 37,92 | 2858 | 49,90 | 1 985 | 51,50 | 3 677 | 47,92 |
|  | 6 to 10 | 54 | 4,53 | 903 | 15,77 | 1 130 | 29,32 | 2 874 | 37,45 |
|  | ≥11 | 4 | 0,34 | 89 | 1,55 | 178 | 4,62 | 284 | 3,70 |
| Length of hospital stay | Median (IQR) | 5 (3-9) |  | 8 (5-13) |  | 11 (6-18) |  | 13 (8-21) |  |
| Follow-up time in days | Median (IQR) | 615 (590-673) |  | 599 (582-637) |  | 597 (580-615) |  | 594 (577-611) |  |
| Time to death in days | Median (IQR) | 57,5 (47-278) |  | 209 (56-377) |  | 211 (74-389) |  | 160 (49-371) |  |
| Controls |  |  |  |  |  |  |  |  |  |
| Age group |  | 19 – 40 |  | 41 – 64 |  | 65 – 74 |  | ≥75 |  |
|  |  | n=11 958 |  | n=59 057 |  | n=43 511 |  | n=102 769 |  |
|  |  | n | % | n | % | n | % | n | % |
| Sex | Male | 6 616 | 55,32 | 36 870 | 62,43 | 25 353 | 58,27 | 47 038 | 45,77 |
|  | Female | 5 342 | 44,67 | 22 187 | 37,56 | 18 158 | 41,73 | 55 731 | 54,22 |
| Polypharmacy | 0 | 9 206 | 76,99 | 40 983 | 69,40 | 24 535 | 56,39 | 42 833 | 41,68 |
|  | 0 or 1 | 10 773 | 90,09 | 47 098 | 79,75 | 27 877 | 64,07 | 48 803 | 47,49 |
|  | 2 to 5 | 1139 | 9,53 | 10 153 | 17,19 | 11 322 | 26,02 | 32 629 | 31,75 |
|  | 6 to 10 | 45 | 0,38 | 1650 | 2,79 | 3878 | 8,91 | 19 285 | 18,77 |
|  | ≥11 | 1 | 0,01 | 156 | 0,26 | 433 | 1,00 | 2 048 | 1,99 |
| Follow-up time in days | Median (IQR) | 614 (590-673) |  | 599 (582-633) |  | 596 (578-613) |  | 591 (574-608) |  |
| Time to death in days | Median (IQR) | 158 (62-441) |  | 130 (53-263) |  | 135 (54-297) |  | 138 (47-314) |  |

**Figure 1:**
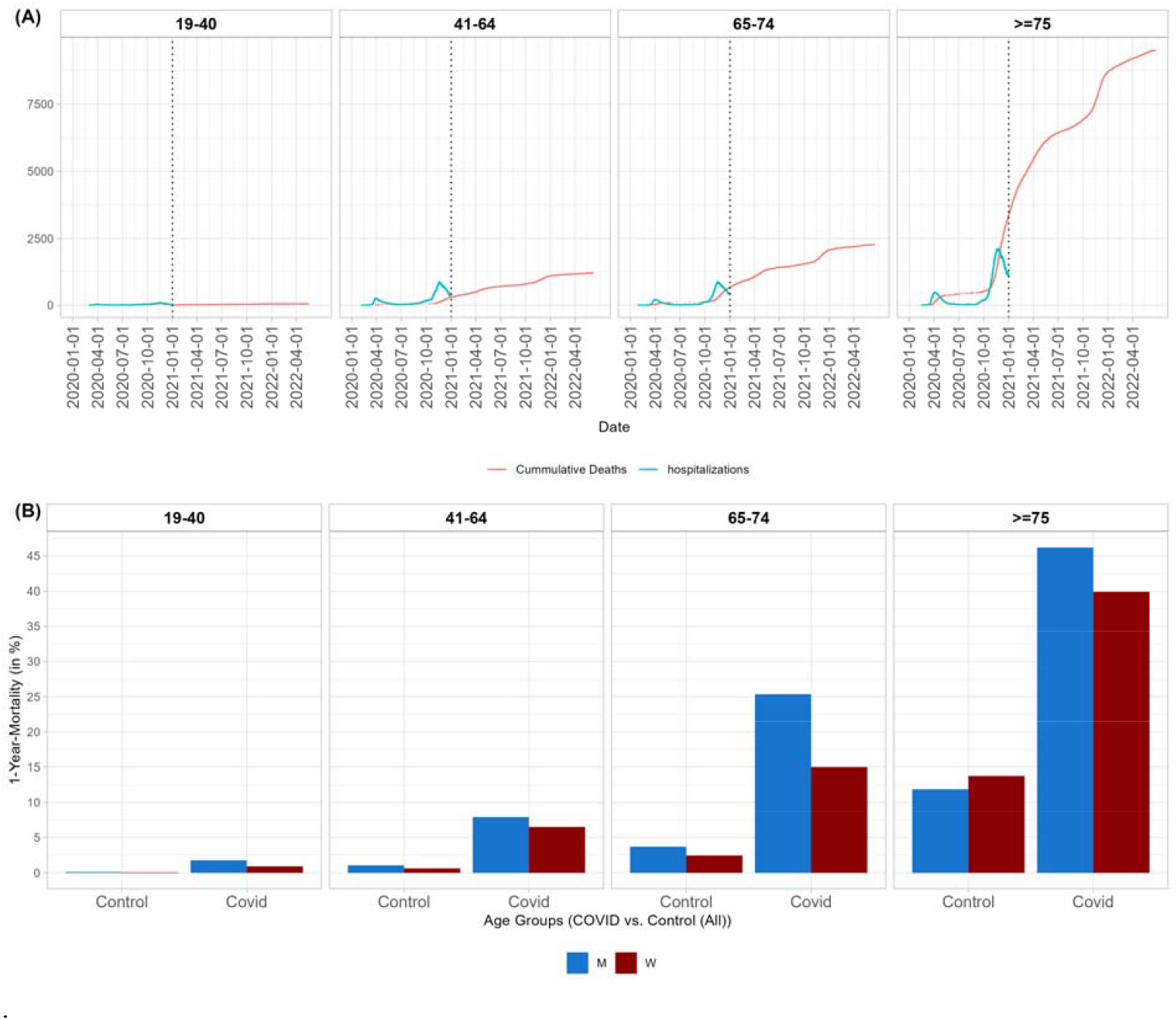
Mortality by age group. (A) Timeline of hospitalization due to COVID-19 in 2020 (hospitalizations per day in blue) and timeline of death (cumulative number of deaths in red) of COVID-19 hospitalized patients. (B) 1-year mortality in age groups and by sex for COVID-19-hospitalized patients and controls. For details on mortality rates for patients and controls, as well as patients surviving the hospital stay with and without propensity score matching, see Tables S8 and S9.

Polypharmacy of more than six medication groups was observed more often in the older age groups (65-74 years and ≥75 years). In the older age groups, 5,62% (65-75 years) and 4,36% (≥75 years) of patients and 56,39% (65-75 years) and 41,68% (≥75 years) of controls did not receive any drug out of the investigated medication groups (Table 1).

Patients hospitalized due to COVID-19 are expected to have more comorbidities and, therefore, more medications than the general population. This was also observed in our Austrian population. Patients received more drugs in all investigated medication groups (Table 2, Tables S4 to S7) across all age groups compared to controls.

**Table 2:**
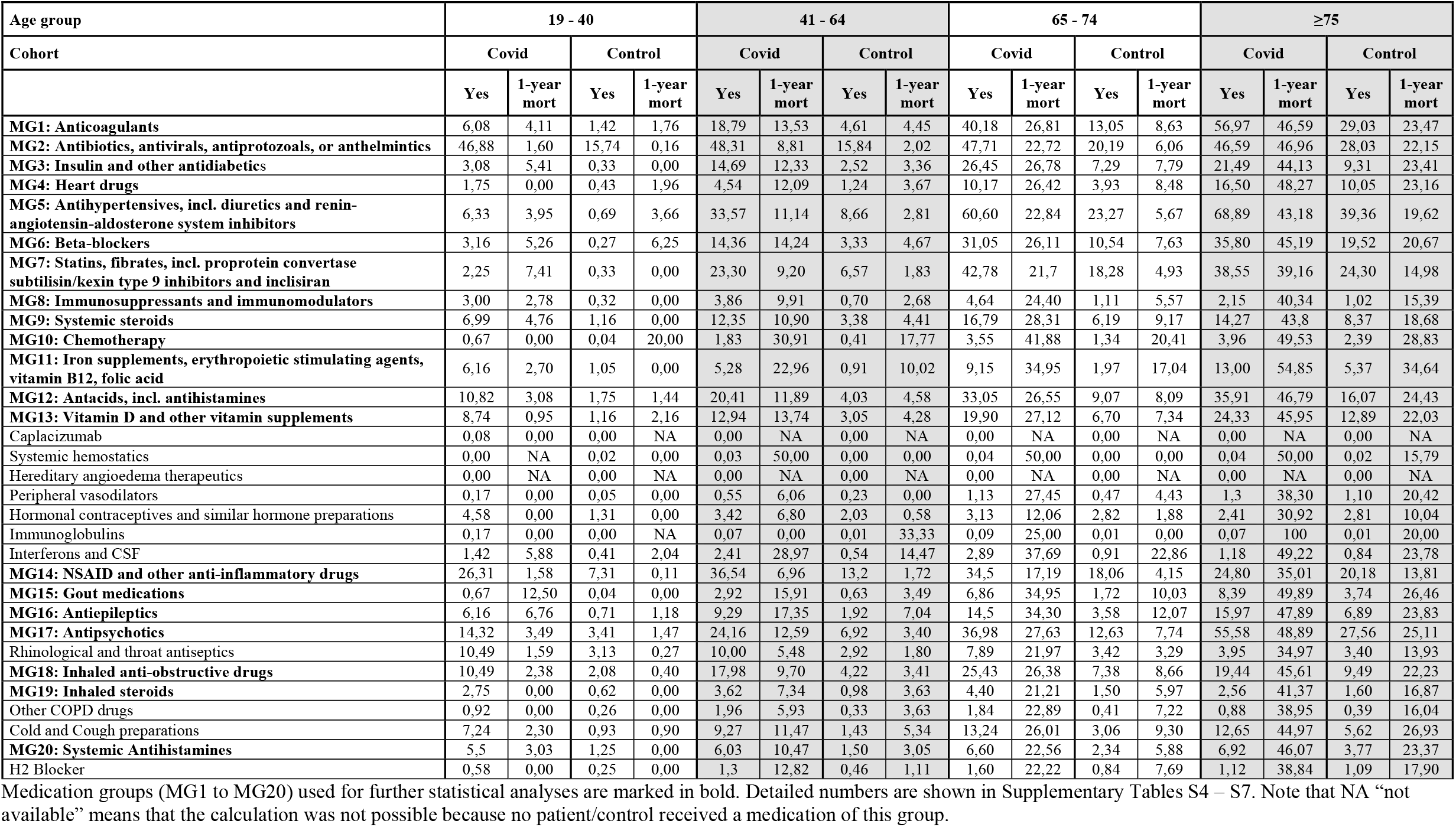
Percentages of COVID-hospitalized patients and controls receiving at least one medication in the investigated medication group and corresponding 1-year mortality by age group.

Interestingly, we observed that 24,2% of patients aged 41-64 years, 37% of patients aged 65-74 years, and 55,6% of patients aged ≥75 years received antipsychotics before COVID hospitalization. In the control group, the rate of prescribed antipsychotics was 6,9% for 41-64 years, 12,6% for 65-74 years, and 27,6% for ≥75 years.

### All-cause mortality for COVID-patients

In the 19-40 years age group, 1,05% of males and 0,37% of females dies in hospital. One-year mortality rates in this age group were 1,8% in males and 0,93% in females. For older age groups, higher mortality rates were observed. In the 41-64 years age group, 5,15% of males and 4,31% of females died during the hospital stay. One-year mortality was 7,91% for males and 6.49% for females. In the 65-74 years age group, 17,66% of males and 9,97% of females died during the hospital stay. Within 1 year after hospital admission, 25,38% of males and 14,96% of females died. Furthermore, 33,56% of males and 25,61% of females aged ≥75 years died during the hospital stay. One-year mortality rates in this age group were 46,2% for males and 39,94% for females (Table S8).

In all age groups, we found a trend of a larger risk of death for men compared to women, which was significant in the older age groups (age >65 years) after multiplicity correction (Table S10). The number of hospital admissions per day and the cumulative number of deaths are shown in Figure 1A. One-year mortality rates are presented in Figure 1B.

Simple Cox-regression models evaluated a significant higher risk of all-cause death for patients with a larger number of prescribed medication groups compared to patients receiving drugs from none of the medication groups or only one medication group. No significant relationship was observed for patients aged 19-41 years (Figure 2A-D and Figure S1).

**Figure 2:**
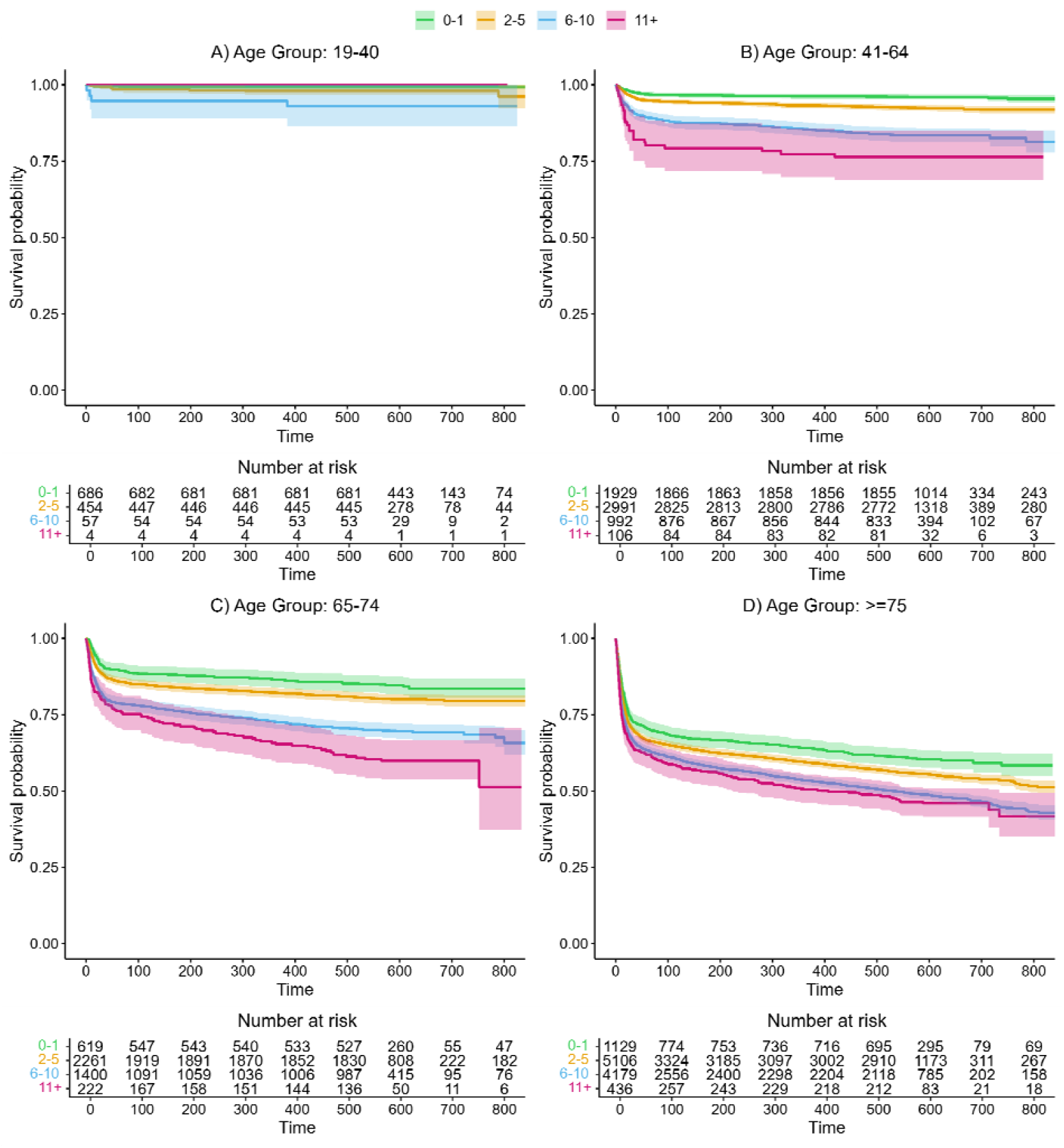
Kaplan-Meier curves and corresponding 95% confidence intervals for polypharmacy groups within COVID-19-hospitalized patients by age group. For details on hazard ratios, confidence intervals, and p-values, see Table S10. Curves are shown separately for patients with drugs in 0-1 medication groups (green), 2-5 medication groups (orange), 6-10 medication groups (blue), and >10 medication groups (magenta).

Due to the large number of medication groups and their potential interactions, polypharmacy is just one factor in a complex system and may not completely explain the associations of prescribed medications with all-cause death after COVID-19. Therefore, we included all 20 medication groups in the statistical Cox regression models to evaluate the effects of individual medication groups. Due to the small number of events in the youngest age group (19-40 years), not all 20 medication groups could be included in the model for this age group.

Figure 3 summarizes the results of the Cox regression model of all-cause mortality when including the twenty medication groups (Table S11. In the youngest group (19-40 years), only prescribed vitamin D and other vitamin supplements (p<0,001) and systemic antihistamines (p=0,002) were significantly associated with survival. For patients receiving vitamin supplements before the COVID-19 hospital stay, we observed a significantly lower risk of death. In addition, systemic antihistamines were significantly associated with poor survival. The other medication groups did not show significant results. However, due to the small number of events in this age group, the results should be interpreted with caution.

**Figure 3:**
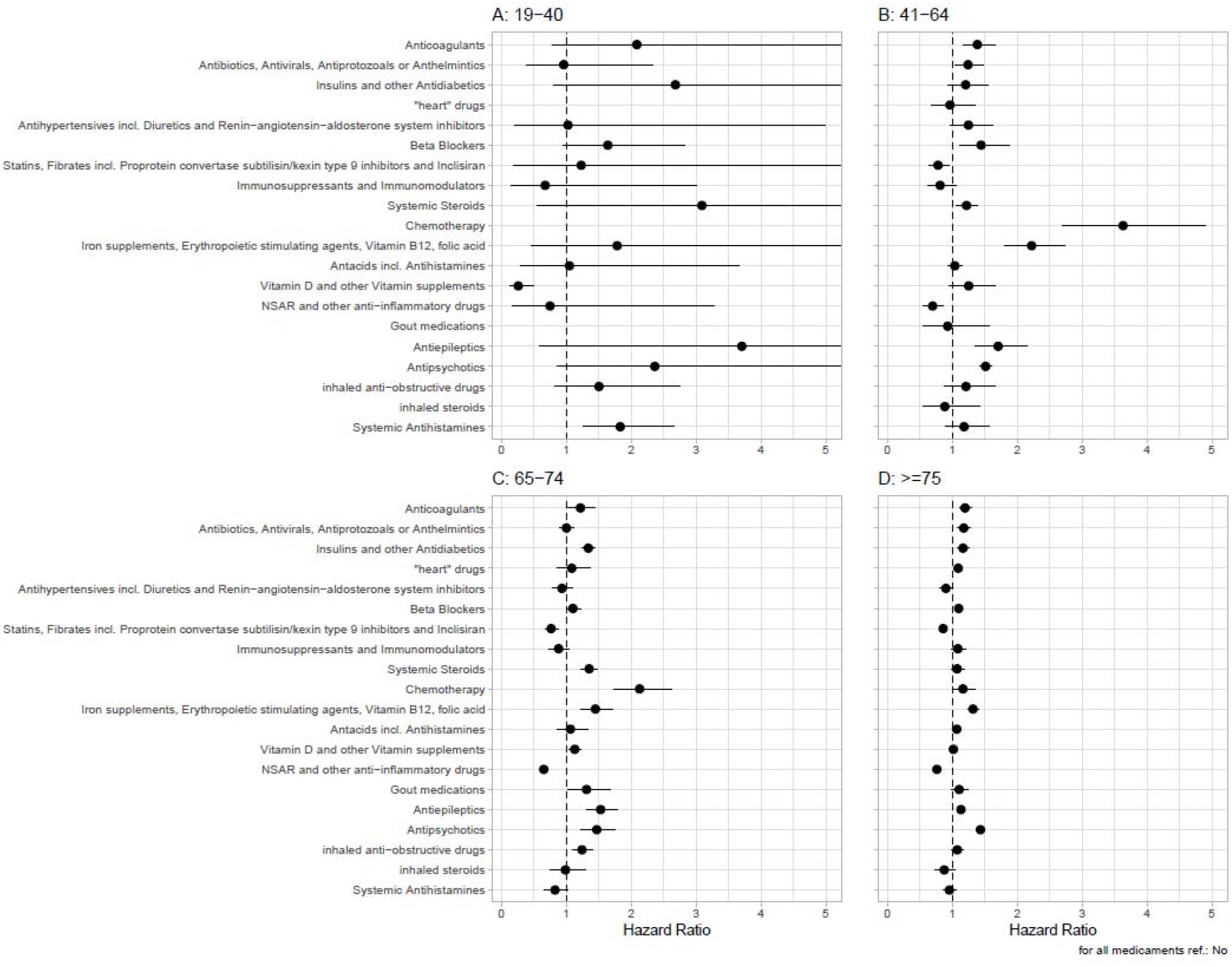
Hazard ratios and 95% confidence intervals for medication groups for the outcome all-cause mortality in COVID-19-hospitalized patients. For details, see Table S11. A hazard ratio >1 indicates a larger risk for all-cause death for patients receiving a drug in the corresponding medication group.

For several medication groups, the results were different between the age groups. Anticoagulants (p<0,001 for 41-64 years and ≥75 years); Antibiotics, antivirals, antiprotozoals, or anthelmintics (p<0,001 for ≥75 years); Insulin and other antidiabetics (p<0,001 for age groups ≥65 years); Heart drugs (p=0,001 for ≥75 years); Beta-blockers (p<0,001 for ≥75 years); Systemic steroids (p<0,001 for 65-74 years); Chemotherapy (p<0,001 for 41-74 years); Antacids (p<0,001 for ≥75 years); Vitamin D or other vitamin supplements (p=0,001 for 64-74 years); and Inhaled anti-obstructive drugs (p<0,001 for 64-74 years) were significantly associated with poor survival. Statins and fibrates were significantly associated with a lower risk of death in patients >64 years old.

In the three age groups >40 years a significant association of antiepileptics (all p<0,001), antipsychotics (all p<0,001), and the group Iron supplements, erythropoietic stimulating agents (ESA), vitamin B12 (B12), and folic acid (FA) (all p<0,001) with a higher risk of all-cause death was observed. Furthermore, for NSAID and other anti-inflammatory drugs, a significant larger probability for survival was observed (all age groups>40: p<0,001). Figure 4 shows the Kaplan-Meier curves for important prescribed medication groups associated with poor survival while Figure S6 shows the Kaplan-Meier curves of medication groups associated with poor survival in the control cohort.

**Figure 4:**
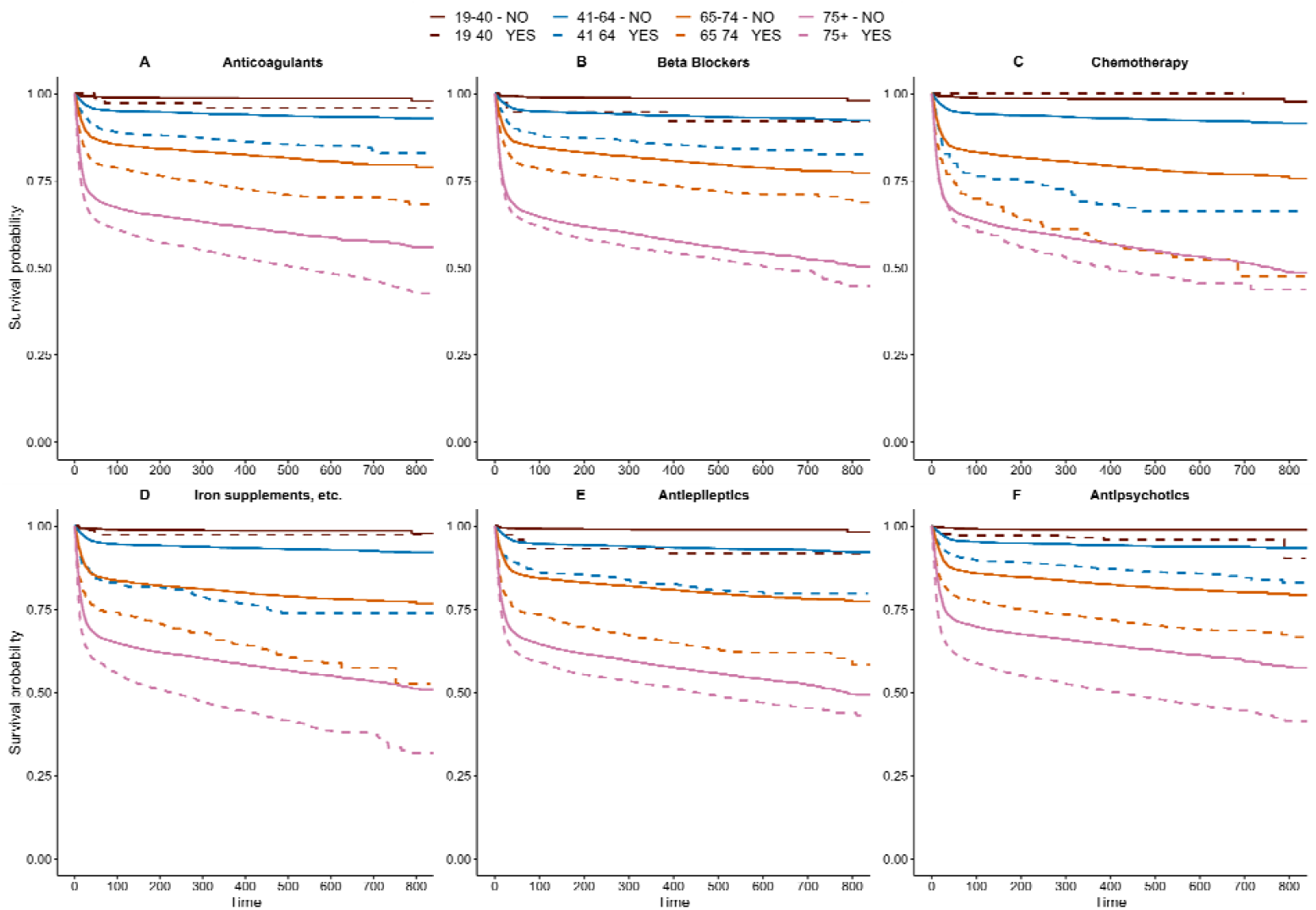
Kaplan-Meier curves of medication groups associated with poor survival of COVID-19 hospitalized patients. For details on hazard ratios, confidence interval, and p-values, see Table S11.

In the models including all investigated medication groups, the factor polypharmacy did not remain significant, indicating that polypharmacy alone may not be a predictor of all-cause death, and specific medication groups may be more important factors for outcomes in COVID-19-hospitalized patients.

### All-cause death after COVID-hospital survival

We also evaluated all-cause death for the subgroup of patients who survived the COVID-19 hospital stay. Among these patients, 0.76% of males and 0.56% of females aged 19-40 years died within the first year after hospital discharge. The 1-year mortality rates were 2.91% and 2.28% for males and females aged 41-64 years, 9.37% and 5.78% for males and females aged 65-74 years, and 19.02% and 19.26% for males and females aged ≥75 years, respectively (Table S9).

In the age groups >40 years, we found significant associations with a higher risk of death after surviving the COVID hospitalization for chemotherapy (all p<0,001) and the medicament group Iron supplements, erythropoietic stimulating agents, B12, and FA (all p<0,001). For antiepileptics, a significantly higher risk of all-cause death was found in the 41-64 and 65-74 years age groups, with an observed trend in the oldest age group (p=0,016). Again, antipsychotics were significantly associated with poor survival in all age groups >40 years (all p<0,001).

NSAID and other anti-inflammatory drugs had a significant association with a lower risk of all-cause death after hospital survival in the age groups >40 years (p<0,001) and statins and fibrates in the two age groups >65 years (both p<0,001). Detailed results are shown in Table S12 and Figure S2.

### Re-hospitalization due to any reason after COVID-hospital survival

For the subgroup of patients who survived the COVID hospital stay, we evaluated the secondary outcome “hospitalization due to any cause.” In the age groups >40 years, a significantly higher risk of readmission with anticoagulants (all p<0,001), antiepileptics (all p<0,001), systemic steroids (all p<0,001), and chemotherapy (all p<0,008) was observed.

We found a trend of a higher risk for re-hospitalization for patients receiving antipsychotics (p<0,003 for 19-64 years and ≥75 years, p=0,006 for 65-74 years). Detailed results are shown in Table S13 and Figure S3.

### Comparison of COVID-Patients to Controls

The age-, sex-, and region-matched control population received less medication than the patient population prior to the hospital stay. Interestingly, in controls without medication, remarkably good survival was observed even in the older age groups (Figure S4, S5), whereas a steep decrease in survival after hospital admission was observed in the COVID patients (Figure S1).

As a sensitivity analysis, we evaluated the difference between COVID-19-hospitalized patients and the Austrian control population concerning all-cause death and hospitalization due to any reasons. As patients had potentially more severe co-morbidities and received more medication, we attempted to account for this imbalance using propensity score matching (PSM) with the 20 medication groups in addition to the age, sex, and region. Due to too small a number of events, this analysis could not be performed for all-cause death in the 19-40 years age group.

For all other subgroups, a significant difference in the risk of all-cause death was found between COVID-19-hospitalized patients and PSM-controls (all p<0,001). For the subgroup of patients surviving the COVID-hospital stay, reduced hazard ratios were observed for the comparison to the PSM-controls. The difference between patients and controls remained significant in the age groups 41-64 years (p<0,001) and 65-74 years (p<0,001), but not in the oldest age group (p=0,078). Kaplan-Meier curves for propensity score-matched COVID patients and controls are shown in Figure S7.

Concerning hospitalization due to any reason, in all four age groups, we observed a significantly greater probability of re-hospitalization among the COVID-19-hospitalized patients compared to controls (all p<0,001). These results may indicate that polypharmacy may not completely explain the worse effect in patients with severe COVID-19 (Table S14, Figure S3).

## Discussion

In this retrospective study, we evaluated whether baseline medication profiles may be associated with survival or hospitalization due to any reason after COVID-19-related hospitalization in an Austrian population. Hospitalized COVID-19 patients had a higher drug prescription load prior to COVID-hospitalization and increased long-term mortality, especially in patients >75 years old. Pre-COVID prescription of antipsychotic drugs, antiepileptic drugs, chemotherapy, iron/FA/ B12, beta-blockers, and anticoagulants was significantly associated with increased mortality, whereas patients who were prescribed NSAIDs and other anti-inflammatory drugs prior to COVID-19 hospitalization had a significantly lower risk of all-cause death. Due to our study design, we were able to present the “real life” prescription and mortality rate of patients hospitalized with the diagnosis of COVID-19 in Austria, as well as in a matched control population that was followed from 2020 for up to a maximum of 880 days.

Polypharmacy frequently occurs in patients with COVID-19^15^ and may be associated with increased morbidity and mortality.^15,16^ Analyzing the prescription data in the control population, we detected no drug prescription in the investigated medication groups in 77% (19-40 years), 69% (41-64 years), 56% (65-74 years), and 41% (≥75 years) of controls. In contrast, 30% (19-40 years), 15% (41-64 years), 6% (65-74 years), and 4% (≥75 years) of COVID-19-hospitalized patients had no drug prescription.

We found that the intake of antipsychotic drugs was associated with a significant increased risk of death. The estimated mortality for patients with prescribed antipsychotic drugs was 56.3% (CI: 54.7 – 57.9) within a two-years follow up in the age group ≥75 years as compared to 40.83% (CI: 42.5 – 39.1) for patients without antipsychotic drugs. The mortality rate for controls with prescribed antipsychotic drugs was 32.9% (CI: 32.1 – 33.7) as compared to patients not receiving these drugs (11.4%, CI: 11.7 – 11.1) within two years follow-up for patients ≥75 years.

The association of anti-inflammatory medication, such as NSAIDs, with lower mortality could be attributed to the pathophysiology of COVID-19, with a high pro-inflammatory state in the second phase of the disease, also referred to as the cytokine storm phase,^19,20^ which could be attenuated by anti-inflammatory medication. In patients with high pro-inflammatory state in need of oxygen therapy and SARS-CoV-2-associated lung infiltration, anti-inflammatory therapy (e.g., dexamethasone) was the only pharmacological intervention to reduce mortality.^21^ However, we cannot exclude the possibility of bias by indication with the selection of possibly healthier patients using NSAIDS on a regular basis because more comorbid patients (e.g., with chronic kidney disease, diabetes, cardiovascular disease, or heart failure) are regularly advised to avoid NSAIDs.

The pronounced association of antipsychotic medication with higher mortality is assumed to be related to the comorbidities in a population that has high prescription rates of such medication, especially at higher ages. Antipsychotic drugs are associated with severe COVID-19 morbidity and mortality.^16^ Antipsychotic medication is mainly prescribed to patients in resident or nursing homes with dementia or behavioral disorders.^22,23^ The number of antipsychotic prescriptions is higher in nursing homes (57,1%) compared to residential homes (29,5%),^24^ emphasizing more frequent prescriptions in a more morbid population. Therefore, chronic psychotic medication use at more advanced ages is most probably indicative of patients with severe impairments.^25-27^

Among nursing home residents, a significant positive association was found between the total number of medications and 30-day COVID-related adjusted mortality.^17^ After additional correction for dementia and use of Proton pump inhibitors (PPI), vitamin D, antipsychotics, and antithrombotics, this effect was no longer significant, suggesting that polypharmacy itself may not be the problem, but the type of medication. In our analyses, polypharmacy did not remain significant after correcting for several medication groups.

In patients who were discharged alive from a COVID-19-related hospitalization, the risk of post discharge death of patients >64 of age within 180 days was nearly twice that observed in historical controls admitted to the hospital with influenza. ^12^ Although readmission after COVID-19-related hospitalization was common, the frequency by 180 days was similar to the frequency of patients discharged alive from influenza-related hospitalization. Furthermore, crude differences in drug use between COVID-19 patients and the general population were found in antithrombotic agents, antiepileptics, anti-gout preparations, and cardiac therapy. ^14^

Scant data are available on clinical outcomes in patients discharged alive from COVID-19 hospitalization. Data from a large study with patients discharged after COVID-19 reported an increased risk of readmission and mortality during a follow-up of 140 days.^10^ In a German cohort of hospitalized COVID-19 patients, 30-day all-cause mortality was 23,9% and 180-day all-cause mortality 29,6%. Another study after COVID-19 hospitalization among patients in Italy reported an 8% age-related overall relative increase in all-cause death after 6 months of follow-up.^13^ However, age was the only independent predictor of mortality after multivariate analysis. Another report from the US investigated the 12-month mortality after recovery from the initial episode of COVID-19 and reported a significantly higher 12-month adjusted all-cause mortality risk for patients with severe COVID-19 compared to both COVID-19-negative patients and mild COVID-19 patients. A large retrospective long-term outcome cohort study indicated an overall 2-year mortality risk that was worse by day 180 among those infected with COVID-19 compared to matched uninfected comparators, but there was no excess mortality during the subsequent 1,5 years.^28^ In our study, we observed increased long-term mortality and increased risk of hospitalization due to any reason after surviving COVID-19 hospitalization. Mortality was more pronounced within the first 50 days after index-hospitalization.

The main strength of this study is the use of a large, representative, real-world national database. The retrospective design, however, is a limitation, which we sought to mitigate by including several potential confounding factors in the statistical models and performing propensity score matching to support meaningful comparisons. Yet, as in any observational research, even with the large sample size and long-term follow-up, unmeasured confounding leading to bias is still possible. The study population was drawn exclusively from the Austrian Health Insurance Funds, raising potential concerns about the generalizability and external validity of the findings to a broader patient population. Furthermore, no information was available from the Austrian Insurance Fund on vaccination or intensive care in hospitals. However, vaccination was first available in the very end of 2020 and therefore, it may not be an important factor for patients hospitalized with COVID-19 in 2020.

In conclusion, this large Austrian cohort of COVID-19-hospitalized patients and matched controls an increased short– and long-term risk of mortality was observed. Patients hospitalized with COVID-19 had a higher drug prescription load (polypharmacy). Antipsychotics were significantly associated with poor survival in patients >40 years old. Our findings may help identify the most vulnerable patients at higher risk of mortality after COVID-19 discharge regardless of age by screening prescribed medication groups, with implications for preventive measures. Antipsychotics are assumed to be an underrecognized medication group linked to worse patient outcomes after COVID-19 hospitalization.

## Acknowledgements

We thank the anonymous referees and editors for their support. This work was financed by ARGE Ankersmit of the surgical research laboratory.

## Author Contributions Statement

H.J.A., B.R., R.W., and A.C.G. were responsible for conceptualization. P.K., B.R., A.C.G., C.W., J.M., and H.J.A. conceived the study and curated the data. C.W., P.K., and A.C.G. cleaned, analyzed, and verified the underlying data. H.J.A., A.C.G., R.W., C.W., and P.K. wrote the paper and visualized the data. A.C.G., B.R., C.W., P.K., D.T-W., M.M., J.M., C.A., J.A., R.W., and H.J.A. commented on the paper, oversaw the analysis, and edited the final manuscript. All authors contributed to drafting the paper and revised the manuscript for important intellectual content.

## Data availability Statement

Data that support the findings of this study are available upon request from the corresponding author.

## References

1) Williamson EJ., Walker AJ., Bhaskaran K., Bacon S., Bate X., Morton CE., Curtis HJ., Mehrkar A., Evans D., Inglesby P., Cockburn J., McDonald H., MacKenna B., Tomlinson L., Douglas IJ., Rentsch CT., Mathur R., Wong AYS., Grieve R., Harrisonh D., Forbes H., Schultze A., Croker R., Parry J., Hester F., Harper S., Perera R., Evans SJW., Liam Smeeth, Goldacre B. (2020). Factors associated with COVID-19-related death using OpenSAFELY, Nature: 584: 430–436

2) Posch M., Bauer P., Posch A., Koenig F. (2020). Analysis of Austrian COVID-19 deaths by age and sex. Wiener Klinische Wochenschrift 132: 685–689.

3) Bauer P., Brugger J., Koenig F., Posch M. (2021). An international comparison of age and sex dependency of COVID-19 deaths in 2020: a descriptive analysis. Scientific reports, 11: 19143.

4) Kautzky-Willer A., Kaleta M., Lindner SD., Leutner M., Thurner S., Klimek P. (2022) Sex Differences in Clinical Characteristics and Outcomes of Patients with SARS-CoV-2 Infection Admitted to Intensive Care Units in Austria. JPM, (12): 4.

5) Zajic P., Hiesmayr M., Bauer P., Baron DM., Gruber A., Joannidis M., Posch M., Metnitz PHG. (2023) Nationwide analysis of hospital admissions and outcomes of patients with SARS-CoV-2 infection in Austria in 2020 and 2021. Scientific Reports: 13: 8548

6) Sze S., Pan D., Nevill CR., Gray LJ., Martin CA., Nazareth J., Minhas JS., Divall P., Khunti K., Abrams KR., Nellums LB., Pareek M. (2020). Ethnicity and clinical outcomes in COVID-19: A systematic review and meta-analysis. Eclinical Medicine, 29-30: 100630.

7) Zheng Z., Peng F., Xu B., Zhao J., Liu H., Peng J., Li, Q., Jiang C., Zhou Y., Liu S., Ye C., Zhang P., Xing Y., Guo H., Tang W. (2020) Risk factors of critical & mortal COVID-19 cases: A systematic literature review and meta-analysis. Journal of Infection, 81: e16–e25

8) Ssentongo P., Ssentongo AE., Heilbrunn ES., Ba DM., Chinchilli VM. (2020). Association of cardiovascular disease and 10 other pre-existing comorbidities with COVID-19 mortality: A systematic review and meta-analysis. Plos One, doi: 10.1371/journal.pone.0238215

9) Liu S., Cao Y., Du T., Zhi Y. (2020). Prevalence of Comorbid Asthma and Related Outcomes in COVID-19: A Systematic Review and Meta-Analysis. J Allerg Clin Immunol Pract: doi: 10.1016/j.jaip.2020.11.054

10) Ayoubkhani D., Khunti K., Nafilyan V., Maddox T, Humberstone B., Diamond I, Banerjee A (2021). Post-covid syndrome in individuals admitted to hospital with covid-19: a retrospective cohort study. British Journal of Medicine, 372: n693

11) Mainous III AG., Rooks BJ., Wu V., Orlando FA. (2021). COVID-19 Post-acute Sequelae among Adults: 12 Month Mortality Risk. Frontiers in Medicine. doi: 10.3389/fmed.2021.778434

12) Oseran AS., Song Y., Xu J., Dahareh IJ., Wadhera RK., Lemos JA., Das SR., Sun T., Yen RW., Kazi DS. (2023) Long term risk of death and readmission after hospital admission with covid-19 among older adults: retrospective cohort study. BMJ: 382: e076222

13) Renda G, Ricci F, Spinoni EG, et al. (2022) Predictors of Mortality and Cardiovascular Outcome at 6 Months after Hospitalization for COVID-19. J Clin Med., 11 (3): 729.

14) Orlando V., Coscioni E., Guarino I., Mucherino S., Perrella A., Trama U., Limongelli G., Menditto E (2021) Drug-utilisation profiles and COVID-19, Scientific Reports, 11: 8913

15) Ghasemi H, Darvishi N, Salari N, Hosseinian-Far A, Akbari H, Mohammadi M. (2022). Global prevalence of polypharmacy among the COVID-19 patients: a comprehensive systematic review and meta-analysis of observational studies. Trop Med Heal., 50 (1): 60.

16) Iloanusi S, Mgbere O, Essien EJ. (2021) Polypharmacy among COVID-19 patients: A systematic review. J Am Pharm Assoc., 61 (5): e14–e25.

17) Visser AGR, Winkens B, Schols JMGA, Janknegt R, Spaetgens B. (2023) The impact of polypharmacy on 30-day COVID-related mortality in nursing home residents: a multicenter retrospective cohort study. Eur Geriatr Med., 14 (1): 51–57.

18) R Core Team (2021). R: A language and environment for statistical computing. R Foundation for Statistical Computing, Vienna, Austria. https://www.R-project.org

19) Fajgenbaum DC, June CH. (2020) Cytokine Storm. N Engl J Med, 383 (23): 2255–2273.

20) Wendt R, Lingitz MT, Laggner M, et al. (2021) Clinical Relevance of Elevated Soluble ST2, HSP27 and 20S Proteasome at Hospital Admission in Patients with COVID-19. Biology. 10 (11): 1186.

21) Group RC, Horby P, Lim WS, et al. (2020). Dexamethasone in Hospitalized Patients with Covid-19. N Engl J Med., 384 (8): 693–704.

22) Ivers NM, Taljaard M, Giannakeas V, Reis C, Williams E, Bronskill S. (2019) Public reporting of antipsychotic prescribing in nursing homes: population-based interrupted time series analyses. BMJ Qual Saf., 28 (2): 121.

23) Rochon PA, Stukel TA, Bronskill SE, et al. (2007) Variation in Nursing Home Antipsychotic Prescribing Rates. Arch Intern Med., 167 (7): 676–683.

24) Chakraborty A, Linton CR. (2012). Antipsychotic prescribing in dementia patients in care homes: proactive in-reach service improved quality of care. Int J Geriatr Psychiatry, 27 (10): 1097–1098. doi:10.1002/gps.2827

25) Lövheim H, Sandman PO, Kallin K, Karlsson S, Gustafson Y. (2006) Relationship between antipsychotic drug use and behavioral and psychological symptoms of dementia in old people with cognitive impairment living in geriatric care. Int Psychogeriatr., 18 (4): 713–726.

26) Gauthier S, Cummings J, Ballard C, et al. (2010). Management of behavioral problems in Alzheimer’s disease. Int Psychogeriatr, 22 (3): 346–372.

27) Coon JT, Abbott R, Rogers M, et al. (2014). Interventions to Reduce Inappropriate Prescribing of Antipsychotic Medications in People With Dementia Resident in Care Homes: A Systematic Review. J Am Méd Dir Assoc, 15(10): 706–718.

28) Iwashyna TJ, Seelye S, Berkowitz TS, et al. (2023) Late Mortality After COVID-19 Infection Among US Veterans vs Risk-Matched Comparators. JAMA Intern Med., 183 (10): 1111–1119.

