## Supplemental Tables and Figures for "Baseline drug treatments and long-term outcomes in COVID-19-hospitalized patients: results of the 2020 AUTCOV study"

+ contributed equally

### Definitions of primary and secondary outcome

This study includes all patients aged >18 years who were hospitalized in Austria due to the main diagnosis COVID-19 (ICD-10 Codes U071, U072, U049) from 1 January 2020 to 31 December 2020. The patient data were available from the Austrian Health Insurance Funds. The data set includes 22 571 patients from the first and second pandemic wave. For each patient, medication prescriptions (based on ATC codes) were available from 1 year before the index COVID hospital stay to study cut-off.

A control group (age-, sex- and region-matched, approximately 1:10) not hospitalized due to the main diagnosis COVID-19 in the year 2020 was randomly chosen from the population registered in the Austrian Health Insurance Funds to represent the general Austrian population. It should be noted, that the control population can (but does not necessarily have to) be hospitalized in the follow-up time due to any reason with the exception of COVID-19. ATC codes for the control group were available from 1 year before the first COVID patient was hospitalized in 2020 up to the study cut-off. The control group included 220 068 cases. Death dates were available at the time of study cut-off.

To evaluate the outcome hospitalization due to any reason for each patient, data were scanned from the index COVID-hospital stay to the study cut-off date. Readmissions due to COVID-19 within 2 weeks after the index hospital stay were considered to belong to the index hospital stay. Notably, for age-, sex-, and region-matched controls, time to death or hospitalization was calculated based on the date of the index hospital stay of the matched patient.

All outcome definitions are summarized in **Table S1**.

**Table S1. Definitions of primary and secondary outcomes**

| Outcome | Definition |
|---|---|
| **All-cause mortality (primary outcome)** | all-cause mortality based on death date<br>for patients: time from COVID-19 hospital admission to death or last follow-up<br>for controls: time from COVID-19 hospital admission of the age-, sex and region-matched COVID-19 patient to death or last follow-up |
| **All-cause mortality after hospital stay (secondary outcome)** | all-cause mortality based on death date for the subgroup of patients surviving hospital stay<br>for patients: time from (alive) COVID-19 hospital discharge to death or last follow-up<br>for controls: time from (alive) COVID-19 hospital discharge of the age-, sex and region-matched COVID-19 patient to death or last follow-up |
| **Hospitalization due to any reason (secondary outcome)** | based on billing information (MEL-Code):<br>for patients: time from (alive) COVID-19 hospital discharge to the first hospital admission (due to any reason) after index hospital stay<br>for controls: time to the first hospital admission after the index COVID –19 hospital discharge of the age-, sex- and region-matched patient |

### Definitions of confounders

The statistical models were calculated separately for four age groups: 19-40 years, 41-64 years, 65-74 years, ≥75 years. Within age groups, the variable age was categorized into the categories described in **Table S2**.

**Table S2: Categorization of age groups**

| Age groups | Categorization within age groups |
|---|---|
| **19-40 years** | AG1: 19-25, AG2: 26-35, AG3: 31-35, AG-R: 36-40 (reference) |
| **41-64 years** | AG1: 41-45, AG2: 46-50, AG3: 51-55, AG4: 56-60, AG-R: 61-64 (reference) |
| **65-74 years** | AG1: 65-70, AG-R: 71-74 (reference) |
| **≥75 years** | AG1: 75-80, AG2: 81-85, AG3: 86-90, AG-R: >90 (reference) |

The confounder "half-year" is a binary confounder being 0 if a patient was hospitalized between 1 January 2020 and 30 June 2020 and 1 if a patients was hospitalized between the 1 July 2020 and 31 December 2020, approximately representing the two pandemic waves.

ATC codes describing prescribed medication were available from the Austrian Health Insurance Funds 1 year before the hospitalization due to COVID-19. ATC codes for medications before hospitalization were summarized into medication groups (Table S3) using binary variables, which were set to 1 if the patient received at least one medicament from the corresponding medication group at least once in the year before COVID-19 hospitalization. Not all medication groups were considered in the statistical models. Considered medication groups are marked as MG1 to MG20. Note that patients receiving systemic hemostatics, peripheral vasodilators, immunoglobulins, interferons, CSF, other COPD drugs, or H2-blockers were underrepresented in all four age groups and not investigated further. Hormonal contraceptives were excluded due to the underrepresentation in older age groups, and cold and cough preparations and rhinological and throat antiseptics were not further investigated because no associations with outcomes were expected.

**Table S3: Definitions of medication groups**

| Medication group | ATC codes |
|---|---|
| **Anticoagulants (MG1)** | B01AA01, B01AA02, B01AA03, B01AA04, B01AA07, B01AA08, B01AA09, B01AA10, B01AA11, B01AA12, B01AB01, B01AB02, B01AB04, B01AB05, B01AB06, B01AB07, B01AB08, B01AB09, B01AB10, B01AB11, B01AB12, B01AB51, B01AC01, B01AC02, B01AC03, B01AC04, B01AC05, B01AC06, B01AC07, B01AC08, B01AC09, B01AC10, B01AC11, B01AC13, B01AC14, B01AC15, B01AC16, B01AC17, B01AC18, B01AC19, B01AC21, B01AC22, B01AC23, B01AC24, B01AC25, B01AC27, B01AC30, B01AC56, B01AD01, B01AD02, B01AD03, B01AD04, B01AD05, B01AD06, B01AD07, B01AD08, B01AD09, B01AD10, B01AD11, B01AD12, B01AE01, B01AE02, B01AE03, B01AE04, B01AE05, B01AE06, B01AE07, B01AF01, B01AF02, B01AF03, B01AF04, B01AX01, B01AX04, B01AX05, B01AX06 |
| **Antibiotics, antivirals, antiprotozoals, or anthelmintics (MG2)** | A07AA01, A07AA02, A07AA03, A07AA04, A07AA05, A07AA06, A07AA07, A07AA08, A07AA09, A07AA10, A07AA11, A07AA12, A07AA13, A07AA51, A07AA54, A07AB02, A07AB03, A07AB04, A07AC01, J01AA01, J01AA02, J01AA03, J01AA04, J01AA05, J01AA06, J01AA07, J01AA08, J01AA09, J01AA10, J01AA11, J01AA12, J01AA13, J01AA14, J01AA15, J01AA20, J01AA56, J01BA01, J01BA02, J01BA52, J01CA01, J01CA02, J01CA03, J01CA04, J01CA05, J01CA06, J01CA07, J01CA08, J01CA09, J01CA10, J01CA11, J01CA12, J01CA13, J01CA14, J01CA15, J01CA16, J01CA17, J01CA18, J01CA19, J01CA20, J01CA51, J01CE01, J01CE02, J01CE03, J01CE04, J01CE05, J01CE06, J01CE07, J01CE08, J01CE09, J01CE10, J01CE30, J01CF01, J01CF02, J01CF03, J01CF04, J01CF05, J01CG01, J01CG02, J01CR01, J01CR02, J01CR03, J01CR04, J01CR05, J01CR50, J01DA01, J01DA02, J01DA03, J01DA04, J01DA05, J01DA06, J01DA07, J01DA08, J01DA09, J01DA10, J01DA11, J01DA12, J01DA13, J01DA14, J01DA15, J01DA16, J01DA17, J01DA18, J01DA19, J01DA21, J01DA22, J01DA23, J01DA24, J01DA25, J01DA26, J01DA27, J01DA30, J01DA31, J01DA32, J01DA33, J01DA34, J01DA35, J01DA36, J01DA37, J01DA38, J01DA39, J01DA40, J01DA41, J01DA42, J01DA63, J01DB01, J01DB02, J01DB03, J01DB04, J01DB05, J01DB06, J01DB07, J01DB08, J01DB09, J01DB10, J01DB11, J01DB12, J01DC01, J01DC02, J01DC03, J01DC04, J01DC05, J01DC06, J01DC07, J01DC08, J01DC09, J01DC10, J01DC11, J01DC12, J01DC13, J01DC14, J01DD01, J01DD02, J01DD03, J01DD04, |

| | |
|---|---|
| | J01DD05, J01DD06, J01DD07, J01DD08, J01DD09, J01DD10, J01DD11, J01DD12, J01DD13, J01DD14, J01DD15, J01DD16, J01DD18, J01DD52, J01DD54, J01DD62, J01DD63, J01DD64, J01DE01, J01DE02, J01DF01, J01DF02, J01DH02, J01DH03, J01DH04, J01DH06, J01DH51, J01DH52, J01DH55, J01DH56, J01DI01, J01DI02, J01DI04, J01DI54, J01EA01, J01EA02, J01EB01, J01EB02, J01EB03, J01EB04, J01EB05, J01EB06, J01EB07, J01EB08, J01EB20, J01EC01, J01EC02, J01EC03, J01EC20, J01ED01, J01ED02, J01ED03, J01ED04, J01ED05, J01ED06, J01ED07, J01ED08, J01ED09, J01ED20, J01EE01, J01EE02, J01EE03, J01EE04, J01EE05, J01EE06, J01EE07, J01FA01, J01FA02, J01FA03, J01FA05, J01FA06, J01FA07, J01FA08, J01FA09, J01FA10, J01FA11, J01FA12, J01FA13, J01FA14, J01FA15, J01FF01, J01FF02, J01FG01, J01FG02, J01GA01, J01GA02, J01GB01, J01GB03, J01GB04, J01GB05, J01GB06, J01GB07, J01GB08, J01GB09, J01GB10, J01GB11, J01GB12, J01GB13, J01GB14, J01MA01, J01MA02, J01MA03, J01MA04, J01MA05, J01MA06, J01MA07, J01MA08, J01MA09, J01MA10, J01MA11, J01MA12, J01MA13, J01MA14, J01MA15, J01MA16, J01MA17, J01MA18, J01MA19, J01MA21, J01MA22, J01MA23, J01MA24, J01MB01, J01MB02, J01MB03, J01MB04, J01MB05, J01MB06, J01MB07, J01RA01, J01RA02, J01RA03, J01RA04, J01XA01, J01XA02, J01XA03, J01XA04, J01XA05, J01XB01, J01XB02, J01XC01, J01XD01, J01XD02, J01XD03, J01XE01, J01XE02, J01XX01, J01XX02, J01XX03, J01XX04, J01XX05, J01XX06, J01XX07, J01XX08, J01XX09, J01XX11, J01XX12, J02AA01, J02AA02, J02AB01, J02AB02, J02AC01, J02AC02, J02AC03, J02AC04, J02AC05, J02AX01, J02AX04, J02AX05, J02AX06, J04AA01, J04AA02, J04AA03, J04AB01, J04AB02, J04AB03, J04AB04, J04AB05, J04AB06, J04AB30, J04AC01, J04AC51, J04AD01, J04AD02, J04AD03, J04AK01, J04AK02, J04AK03, J04AK04, J04AK05, J04AK06, J04AK07, J04AM01, J04AM02, J04AM03, J04AM04, J04AM05, J04AM06, J04AM07, J04AM08, J04BA01, J04BA02, J04BA03, P01AA01, P01AA02, P01AA04, P01AA05, P01AA52, P01AB01, P01AB02, P01AB03, P01AB04, P01AB05, P01AB06, P01AB07, P01AC01, P01AC02, P01AC03, P01AC04, P01AR01, P01AR02, P01AR03, P01AR53, P01AX01, P01AX02, P01AX04, P01AX05, P01AX06, P01AX07, P01AX08, P01AX09, P01AX10, P01AX11, P01AX52, P01BA01, P01BA02, P01BA03, P01BA06, P01BA07, P01BB01, P01BB02, P01BB51, P01BC01, P01BC02, P01BD01, P01BD51, P01BE01, P01BE02, P01BE03, P01BE04, P01BE05, P01BE52, P01BF01, P01BF02, P01BF03, P01BF04, P01BF05, P01BF06, P01BX01, P01CA02, P01CA03, P01CB01, P01CB02, P01CC01, P01CC02, P01CD01, P01CD02, P01CX01, P01CX02, P01CX03, P01CX04, P02BA01, P02BA02, P02BB01, P02BX01, P02BX02, P02BX03, P02BX04, P02CA01, P02CA02, P02CA03, P02CA04, P02CA05, P02CA06, P02CA51, P02CB01, P02CB02, P02CC01, P02CC02, P02CE01, P02CF01, P02CX01, P02CX02, P02CX03, P02DA01, P02DX01, P02DX02, P03AA01, P03AA02, P03AA03, P03AA04, P03AA05, P03AA54, P03AB01, P03AB02, P03AB51, P03AC01, P03AC02, P03AC03, P03AC04, P03AC51, P03AC52, P03AC53, P03AC54, P03AX01, P03AX02, P03AX03, P03AX04, P03AX05, P03BA01, P03BA02, P03BA03, P03BA04, P03BX01, P03BX02, P03BX03, P03BX04, P03BX05, P03BX06 |
| **Insulin and other antidiabetics (MG3)** | A10AB01, A10AB02, A10AB03, A10AB04, A10AB05, A10AB06, A10AB30, A10AC01, A10AC02, A10AC03, A10AC04, A10AC30, A10AD01, A10AD02, A10AD03, A10AD04, A10AD05, A10AD06, A10AD30, A10AE01, A10AE02, A10AE03, A10AE04, A10AE05, A10AE06, A10AE30, A10AE54, A10AE56, A10AF01, A10BA01, A10BA02, A10BA03, A10BB01, A10BB02, A10BB03, A10BB04, A10BB05, A10BB06, A10BB07, A10BB08, A10BB09, A10BB10, A10BB11, A10BB12, A10BB31, A10BC01, A10BD01, A10BD02, A10BD03, A10BD04, A10BD05, A10BD06, A10BD07, A10BD08, A10BD09, A10BD10, A10BD11, A10BD12, A10BD13, A10BD14, A10BD15, A10BD16, A10BD17, A10BD18,A10BD19, A10BD20, A10BD21, A10BD22, A10BD23, A10BD24, A10BD25, A10BD26, A10BF01, A10BF02, A10BF03, A10BG01, A10BG02, A10BG03, A10BG04, A10BH01, A10BH02, A10BH03, A10BH04, A10BH05, A10BH06, A10BH07, A10BH08, A10BH52, A10BJ01, A10BJ02, A10BJ03, A10BJ04, A10BJ05, A10BJ06, A10BK01, A10BK02, A10BK03, A10BK04, A10BK05, A10BK06, A10BK07, A10BX01, A10BX02, A10BX03, A10BX04, A10BX05, A10BX06, A10BX07, A10BX08, A10BX09, A10BX10, A10BX11, A10BX12, A10BX13, A10BX14, A10XA01 |

| | |
|---|---|
| **Heart drugs (MG4)** | C01AA01, C01AA02, C01AA03, C01AA04, C01AA05, C01AA06, C01AA07, C01AA08,C01AA09, C01AA52, C01AB01, C01AB51, C01AC01, C01AC03, C01AX02, C01BA01, C01BA02, C01BA03, C01BA04, C01BA05, C01BA08, C01BA12, C01BA51, C01BA71, C01BB01, C01BB02, C01BB03, C01BB04, C01BC03, C01BC04, C01BC07, C01BC08, C01BD01, C01BD02, C01BD03, C01BD04, C01BD05, C01BD06, C01BD07, C01BG01, C01BG07, C01BG11, C01CA01, C01CA02, C01CA03, C01CA04, C01CA05, C01CA06, C01CA07, C01CA08, C01CA09, C01CA10, C01CA11, C01CA12, C01CA13, C01CA14, C01CA15, C01CA16, C01CA17, C01CA18, C01CA19, C01CA21, C01CA22, C01CA23, C01CA24, C01CA25, C01CA26, C01CA30, C01CA51, C01CE01, C01CE02, C01CE03, C01CE04, C01CX06, C01CX07, C01CX08, C01CX09, C01DA02, C01DA04, C01DA05, C01DA07, C01DA08, C01DA09, C01DA13, C01DA14, C01DA20, C01DA38, C01DA52, C01DA54, C01DA55, C01DA57, C01DA58, C01DA59, C01DA63, C01DA70, C01DB01, C01DX01, C01DX02, C01DX03, C01DX04, C01DX05, C01DX06, C01DX07, C01DX08, C01DX09, C01DX10, C01DX11, C01DX12, C01DX13, C01DX14, C01DX15, C01DX16, C01DX18, C01DX19, C01DX51, C01DX52, C01DX53, C01DX54, C01EA01, C01EB02, C01EB03, C01EB04, C01EB05, C01EB06, C01EB07, C01EB09, C01EB10, C01EB11, C01EB12, C01EB13, C01EB15, C01EB16, C01EB17, C01EB18, C01EB19, C01EB21 |
| **Antihypertensives, including diuretics and renin-angiotensin-aldosterone system inhibitors (MG5)** | C02AA01, C02AA02, C02AA03, C02AA04, C02AA05, C02AA06, C02AA07, C02AA52, C02AA53, C02AA57, C02AB01, C02AB02, C02AC01, C02AC02, C02AC04, C02AC05, C02AC06, C02BA01, C02BB01, C02CA01, C02CA02, C02CA03, C02CA04, C02CA06, C02CA08, C02CC01, C02CC02, C02CC03, C02CC04, C02CC05, C02CC06, C02CC07, C02DA01, C02DB01, C02DB02, C02DB03, C02DB04, C02DC01, C02DD01, C02DG01, C02KA01, C02KB01, C02KC01, C02KD01, C02KX01, C02KX02, C02KX03, C02KX04, C02KX05, C02KX52, C02LA01, C02LA02, C02LA03, C02LA04, C02LA07, C02LA08, C02LA09, C02LA50, C02LA51, C02LA52, C02LA71, C02LB01, C02LC01, C02LC05, C02LC51, C02LE01, C02LF01, C02LG01, C02LG02, C02LG03, C02LG51, C02LG73, C02LK01, C02LL01, C02LX01, C03AA01, C03AA02, C03AA03, C03AA04, C03AA05, C03AA06, C03AA07, C03AA08, C03AA09, C03AA13, C03AB01, C03AB02, C03AB03, C03AB04, C03AB05, C03AB06, C03AB07, C03AB08, C03AB09, C03AH01, C03AH02, C03AX01, C03BA02, C03BA03, C03BA04, C03BA05, C03BA07, C03BA08, C03BA09, C03BA10, C03BA11, C03BA12, C03BA13, C03BA82, C03BB02, C03BB03, C03BB04, C03BB05, C03BB07, C03BC01, C03BD01, C03BX03, C03CA01, C03CA02, C03CA03, C03CA04, C03CB01, C03CB02, C03CC01, C03CC02, C03CD01, C03CX01, C03DA01, C03DA02, C03DA03, C03DA04, C03DB01, C03DB02, C03EA01, C03EA02, C03EA03, C03EA04, C03EA05, C03EA06, C03EA07, C03EA12, C03EA13, C03EA14, C03EB01, C03EB02, C08CA01, C08CA02, C08CA03, C08CA04, C08CA05, C08CA06, C08CA07, C08CA08, C08CA09, C08CA10, C08CA11, C08CA12, C08CA13, C08CA14, C08CA15, C08CA16, C08CA51, C08CA55, C08CX01, C08DA01, C08DA02, C08DA51, C08DB01, C08EA01, C08EA02, C08EX01, C08EX02, C08GA01, C08GA02, C09AA01, C09AA02, C09AA03, C09AA04, C09AA05, C09AA06, C09AA07, C09AA08, C09AA09, C09AA10, C09AA11, C09AA12, C09AA13, C09AA14, C09AA15, C09AA16, C09BA01, C09BA02, C09BA03, C09BA04, C09BA05, C09BA06, C09BA07, C09BA08, C09BA09, C09BA12, C09BA13, C09BA15, C09BB02, C09BB03, C09BB04, C09BB05, C09BB06, C09BB07, C09BB10, C09BB12, C09BX01, C09BX03, C09BX04, C09BX05, C09CA01, C09CA02, C09CA03, C09CA04, C09CA05, C09CA06, C09CA07, C09CA08, C09CA09, C09DA01, C09DA02, C09DA03, C09DA04, C09DA06, C09DA07, C09DA08, C09DA10, C09DB01, C09DB02, C09DB04, C09DB05, C09DB06, C09DB07, C09DB09, C09DX01, C09DX02, C09DX03, C09DX04, C09DX05, C09DX06, C09DX07, C09XA01, C09XA02, C09XA52, C09XA53, C09XA54 |
| **Beta-blockers (MG6)** | C07AA01, C07AA02, C07AA03, C07AA05, C07AA06, C07AA07, C07AA12, C07AA14, C07AA15, C07AA16, C07AA17, C07AA19, C07AA23, C07AA27, C07AA57, C07AB01, C07AB02, C07AB03, C07AB04, C07AB05, C07AB06, C07AB07, C07AB08, C07AB09, C07AB10, C07AB11, C07AB12, C07AB13, C07AB14, C07AB52, C07AG01, C07AG02, C07BA02, C07BA05, C07BA06, C07BA07, C07BA12, C07BA68, C07BB02, C07BB03, C07BB04, C07BB06, C07BB07, C07BB12, C07BB52, C07BG01, C07CA02, C07CA03, C07CA17, C07CA23, C07CB02, C07CB03, C07CB53, C07CG01, C07DA06, C07DB01, C07FA05, C07FB02, C07FB03, C07FB07, C07FB13, C07FX01, C07FX02, C07FX03, C07FX04, C07FX05, C07FX06 |
| **Statins, fibrates, including proprotein convertase subtilisin/kexin type 9 inhibitors and inclisiran (MG7)** | C10AA01, C10AA02, C10AA03, C10AA04, C10AA05, C10AA06, C10AA07, C10AA08, C10AA51, C10AA52, C10AA53, C10AA55, C10AB01, C10AB02, C10AB03, C10AB04, C10AB05, C10AB06, C10AB07, C10AB08, C10AB09, C10AB10, C10AB11, C10AX06, C10AX07, C10AX08, C10AX09, C10AX10, C10AX11, C10AX12, C10AX13, C10AX14, C10AX15, C10AX16, C10BA01, C10BA02, C10BA03, C10BA04, C10BA05, C10BA06, C10BA07, C10BA08, C10BA09, C10BA10, C10BX01, C10BX02, C10BX03, C10BX04, C10BX05, C10BX06, C10BX07, C10BX08, C10BX09, C10BX10, C10BX11, C10BX12, C10BX13, C10BX14, C10BX15, C10BX16, C10BX17, C10BX18 |
| **Immunosuppressants and immunomodulators (MG8)** | D11AH05, L04AA01, L04AA02, L04AA03, L04AA04, L04AA05, L04AA06, L04AA08, L04AA09, L04AA10, L04AA11, L04AA12, L04AA13, L04AA14, L04AA15, L04AA16, L04AA17, L04AA18, L04AA19, L04AA21, L04AA22, L04AA23, L04AA24, L04AA25, L04AA26, L04AA27, L04AA28, L04AA29, L04AA31, L04AA32, L04AA33, L04AA34, |

| | |
|---|---|
| | L04AA36, L04AA37, L04AA38, L04AA39, L04AA40, L04AA41, L04AA42, L04AA43, L04AA44, L04AA45, L04AA46, L04AB01, L04AB02, L04AB03, L04AB04, L04AB05, L04AB06, L04AB07, L04AC01, L04AC02, L04AC03, L04AC04, L04AC05, L04AC06, L04AC07, L04AC08, L04AC09, L04AC10, L04AC11, L04AC12, L04AC13, L04AC14, L04AC15, L04AC16, L04AC17, L04AC18, L04AC19, L04AD01, L04AD02, L04AD03, L04AX01, L04AX02, L04AX03, L04AX04, L04AX05, L04AX06, L04AX07, L04AX08 |
| **Systemic steroids (MG9)** | H02AA01, H02AA02, H02AA03, H02AB01, H02AB02, H02AB03, H02AB04, H02AB05, H02AB06, H02AB07, H02AB08, H02AB09, H02AB10, H02AB11, H02AB12, H02AB13, H02AB14, H02AB15, H02AB17, H02BX01, H02CA01, H02CA02, H02CA03 |
| **Chemotherapy (MG10)** | L01AA01, L01AA02, L01AA03, L01AA05, L01AA06, L01AA07, L01AA08, L01AA09, L01AB01, L01AB02, L01AB03, L01AC01, L01AC02, L01AC03, L01AD01, L01AD02, L01AD03, L01AD04, L01AD05, L01AD06, L01AD07, L01AD08, L01AG01, L01AX01, L01AX02, L01AX03, L01AX04, L01BA01, L01BA03, L01BA04, L01BB02, L01BB03, L01BB04, L01BB05, L01BB06, L01BB07, L01BC01, L01BC02, L01BC03, L01BC04, L01BC05, L01BC06, L01BC07, L01BC08, L01BC09, L01BC52, L01BC53, L01BC59, L01CA01, L01CA02, L01CA03, L01CA04, L01CA05, L01CB01, L01CB02, L01CC01, L01CD01, L01CD02, L01CD03, L01CD04, L01CE01, L01CE02, L01CE03, L01CE04, L01CX01, L01DA01, L01DB01, L01DB02, L01DB03, L01DB04, L01DB05, L01DB06, L01DB07, L01DB08, L01DB09, L01DB10, L01DB11, L01DC01, L01DC02, L01DC03, L01DC04, L01EA01, L01EA02, L01EA03, L01EA04, L01EA05, L01EB01, L01EB02, L01EB03, L01EB04, L01EB05, L01EB07, L01EB08, L01EC01, L01EC02, L01EC03, L01ED01, L01ED02, L01ED03, L01ED04, L01ED05, L01EE01, L01EE02, L01EE03, L01EF01, L01EF02, L01EF03, L01EG01, L01EG02, L01EH01, L01EH02, L01EJ01, L01EJ02, L01EK01, L01EK03, L01EL01, L01EL02, L01EM01, L01EM02, L01EM03, L01EX01, L01EX02, L01EX03, L01EX04, L01EX05, L01EX07, L01EX08, L01EX09, L01EX10, L01EX11, L01EX12, L01EX13, L01EX14, L01XA01, L01XA02, L01XA03, L01XA04, L01XA05, L01XB01, L01XC01, L01XC02, L01XC03, L01XC04, L01XC05, L01XC06, L01XC07, L01XC08, L01XC09, L01XC10, L01XC11, L01XC12, L01XC13, L01XC14, L01XC15, L01XC16, L01XC17, L01XC18, L01XC19, L01XC21, L01XC22, L01XC23, L01XC24, L01XC25, L01XC26, L01XC27, L01XC28, L01XC29, L01XC31, L01XC32, L01XC33, L01XC34, L01XC35, L01XC36, L01XC37, L01XC38, L01XC39, L01XC40, L01XC41, L01XD01, L01XD02, L01XD03, L01XD04, L01XD05, L01XD06, L01XD07, L01XE01, L01XE02, L01XE03, L01XE04, L01XE05, L01XE06, L01XE07, L01XE08, L01XE09, L01XE10, L01XE11, L01XE12, L01XE13, L01XE14, L01XE15, L01XE16, L01XE17, L01XE18, L01XE21, L01XE23, L01XE24, L01XE25, L01XE26, L01XE27, L01XE28, L01XE29, L01XE31, L01XE33, L01XE34, L01XE35, L01XE36, L01XE37, L01XE38, L01XE39, L01XE41, L01XE42, L01XE43, L01XE44, L01XE45, L01XE46, L01XE47, L01XE48, L01XE50, L01XE51, L01XE52, L01XE53, L01XE54, L01XE56, L01XE57, L01XF01, L01XF02, L01XF03, L01XG01, L01XG02, L01XG03, L01XH01, L01XH02, L01XH03, L01XH05, L01XJ01, L01XJ02, L01XJ03, L01XK01, L01XK02, L01XK03, L01XK04, L01XX01, L01XX02, L01XX03, L01XX05, L01XX07, L01XX08, L01XX09, L01XX10, L01XX11, L01XX14, L01XX16, L01XX17, L01XX18, L01XX19, L01XX22, L01XX23, L01XX24, L01XX25, L01XX27, L01XX28, L01XX29, L01XX31, L01XX32, L01XX33, L01XX34, L01XX35, L01XX36, L01XX37, L01XX38, L01XX39, L01XX40, L01XX41, L01XX42, L01XX43, L01XX44, L01XX45, L01XX46, L01XX47, L01XX48, L01XX50, L01XX51, L01XX52, L01XX53, L01XX54, L01XX55, L01XX56, L01XX57, L01XX58, L01XX59, L01XX60, L01XX61, L01XX62, L01XX63, L01XX64, L01XX65, L01XX66, L01XX67, L01XX68, L01XX70, L01XX71, L01XY01, L01XY02, L02AA01, L02AA02, L02AA03, L02AA04, L02AB01, L02AB02, L02AB03, L02AE01, L02AE02, L02AE03, L02AE04, L02AE05, L02AE51, L02BA01, L02BA02, L02BA03, L02BB01, L02BB02, L02BB03, L02BB04, L02BB05, L02BB06, L02BG01, L02BG02, L02BG03, L02BG04, L02BG05, L02BG06, L02BX01, L02BX02, L02BX03 |
| **Iron supplements, erythropoietic stimulating agents, vitamin B12, folic acid (MG11)** | B03AA01, B03AA02, B03AA03, B03AA04, B03AA05, B03AA06, B03AA07, B03AA08, B03AA09, B03AA10, B03AA11, B03AA12, B03AB01, B03AB02, B03AB03, B03AB04, B03AB05, B03AB06, B03AB07, B03AB08, B03AB09, B03AB10, B03AC01, B03AC02, B03AC03, B03AC05, B03AC06, B03AC07, B03AD01, B03AD02, B03AD03, B03AD04, B03AD05, B03AE01, B03AE02, B03AE03, B03AE04, B03AE10, B03BA01, B03BA02, B03BA03, B03BA04, B03BA05, B03BA51, B03BA53, B03BB01, B03BB51, B03XA01, B03XA02, B03XA03, B03XA05, B03XA06 |
| **Antacids, including antihistamines (MG12)** | A02BB01, A02BB02, A02BC01, A02BC02, A02BC03, A02BC04, A02BC05, A02BC06, A02BC08 |
| **Vitamin D and other vitamin supplements (MG13)** | A11CA01, A11CA02, A11CC01, A11CC02, A11CC03, A11CC04, A11CC05, A11CC06, A11CC07, A11CC20, A11CC55, A11DA01, A11DA02, A11DA03, A11GA01, A11GB01, A11HA01, A11HA02, A11HA03, A11HA04, A11HA05, A11HA06, A11HA07, A11HA08, A11HA30, A11HA31, A11HA32 |
| **Caplacizumab** | B01AX07 |
| **Systemic hemostatics** | B02BX01, B02BX02, B02BX03, B02BX04, B02BX05, B02BX06, B02BX07, B02BX08, B02BX09 |
| **Hereditary angioedema therapeutics** | B06AC01, B06AC02, B06AC03, B06AC04, B06AC05 |

| | |
|---|---|
| **Peripheral vasodilators** | C04AA01, C04AA02, C04AA31, C04AB01, C04AB02, C04AC01, C04AC02, C04AC03, C04AC07, C04AD01, C04AD02, C04AD03, C04AD04, C04AE01, C04AE02, C04AE04, C04AE51, C04AE54, C04AF01, C04AX01, C04AX02, C04AX07, C04AX10, C04AX11, C04AX13, C04AX17, C04AX19, C04AX20, C04AX21, C04AX23, C04AX24, C04AX26, C04AX27, C04AX28, C04AX30, C04AX32 |
| **Hormonal contraceptives and similar hormone preparations** | G03AA01, G03AA02, G03AA03, G03AA04, G03AA05, G03AA06, G03AA07, G03AA08, G03AA09, G03AA10, G03AA11, G03AA12, G03AA13, G03AA14, G03AA15, G03AA16, G03AA17, G03AB01, G03AB02, G03AB03, G03AB04, G03AB05, G03AB06, G03AB07, G03AB08, G03AB09, G03AC01, G03AC02, G03AC03, G03AC04, G03AC05, G03AC06, G03AC07, G03AC08, G03AC09, G03AC10, G03AD01, G03AD02, G03BA01, G03BA02, G03BA03, G03BB01, G03BB02, G03CA01, G03CA03, G03CA04, G03CA06, G03CA07, G03CA09, G03CA53, G03CA57, G03CB01, G03CB02, G03CB03, G03CB04, G03CC02, G03CC03, G03CC04, G03CC05, G03CC06, G03CC07, G03CX01, G03DA01, G03DA02, G03DA03, G03DA04, G03DB01, G03DB02, G03DB03, G03DB04, G03DB05, G03DB06, G03DB07, G03DB08, G03DC01, G03DC02, G03DC03, G03DC04, G03DC05, G03DC06, G03DC31, G03EA01, G03EA02, G03EA03, G03EK01, G03FA01, G03FA02, G03FA03, G03FA04, G03FA05, G03FA06, G03FA07, G03FA08, G03FA09, G03FA10, G03FA11, G03FA12, G03FA13, G03FA14, G03FA15, G03FA16, G03FA17, G03FB01, G03FB02, G03FB03, G03FB04, G03FB05, G03FB06, G03FB07, G03FB08, G03FB09, G03FB10, G03FB11, G03GA01, G03GA02, G03GA03, G03GA04, G03GA05, G03GA06, G03GA07, G03GA08, G03GA09, G03GA10, G03GA30, G03GB01, G03GB02, G03GB03, G03HA01, G03HB01, G03XA01, G03XA02, G03XB01, G03XB02, G03XC01, G03XC02, G03XC03, G03XX01 |
| **Immunoglobulins** | J06BA01, J06BA02 |
| **Interferons and CSF** | L03AA02, L03AA03, L03AA09, L03AA10, L03AA12, L03AA13, L03AA14, L03AA16, L03AA17, L03AB01, L03AB02, L03AB03, L03AB04, L03AB05, L03AB06, L03AB07, L03AB08, L03AB09, L03AB10, L03AB11, L03AB12, L03AB13, L03AB15, L03AB60, L03AB61, L03AC01, L03AC02, L03AX01, L03AX02, L03AX03, L03AX04, L03AX05, L03AX07, L03AX08, L03AX09, L03AX10, L03AX11, L03AX12, L03AX13, L03AX14, L03AX15, L03AX16, L03AX17, L03AX21 |
| **NSAID and other anti-inflammatory drugs (MG14)** | M01AA01, M01AA02, M01AA03, M01AA05, M01AA06, M01AB01, M01AB02, M01AB03, M01AB04, M01AB05, M01AB06, M01AB07, M01AB08, M01AB09, M01AB10, M01AB11, M01AB12, M01AB13, M01AB14, M01AB15, M01AB16, M01AB17, M01AB51, M01AB55, M01AC01, M01AC02, M01AC04, M01AC05, M01AC06, M01AC56, M01AE01, M01AE02, M01AE03, M01AE04, M01AE05, M01AE06, M01AE07, M01AE08, M01AE09, M01AE10, M01AE11, M01AE12, M01AE13, M01AE14, M01AE15, M01AE16, M01AE17, M01AE18, M01AE51, M01AE52, M01AE53, M01AE56, M01AG01, M01AG02, M01AG03, M01AG04, M01AH01, M01AH02, M01AH03, M01AH04, M01AH05, M01AH06, M01AH07, M01AX01, M01AX02, M01AX04, M01AX05, M01AX07, M01AX12, M01AX13, M01AX14, M01AX17, M01AX18, M01AX21, M01AX22, M01AX23, M01AX24, M01AX25, M01AX26, M01AX68, M01BA01, M01BA02, M01BA03, M01CA03, M01CB01, M01CB02, M01CB03, M01CB04, M01CB05, M01CC01, M01CC02 |
| **Gout medications (MG15)** | M04AA01, M04AA02, M04AA03, M04AA51, M04AB01, M04AB02, M04AB03, M04AB04, M04AB05, M04AC01, M04AC02, M04AX01 |
| **Antiepileptics (MG16)** | N03AA01, N03AA02, N03AA03, N03AA04, N03AA30, N03AB01, N03AB02, N03AB03, N03AB04, N03AB05, N03AB52, N03AB54, N03AC01, N03AC02, N03AC03, N03AD01, N03AD02, N03AD03, N03AD51, N03AE01, N03AF01, N03AF02, N03AF03, N03AF04, N03AG01, N03AG02, N03AG03, N03AG04, N03AG05, N03AG06, N03AX03, N03AX07, N03AX09, N03AX10, N03AX11, N03AX12, N03AX13, N03AX14, N03AX15, N03AX16, N03AX17, N03AX18, N03AX21, N03AX22, N03AX23, N03AX24, N03AX30 |
| **Antipsychotics (MG17)** | N05AA01, N05AA02, N05AA03, N05AA04, N05AA05, N05AA06, N05AA07, N05AB01, N05AB02, N05AB03, N05AB04, N05AB05, N05AB06, N05AB07, N05AB08, N05AB09, N05AB10, N05AC01, N05AC02, N05AC03, N05AC04, N05AD01, N05AD02, N05AD03, N05AD04, N05AD05, N05AD06, N05AD07, N05AD08, N05AD09, N05AE01, N05AE02, N05AE03, N05AE04, N05AF01, N05AF02, N05AF03, N05AF04, N05AF05, N05AG01, N05AG02, N05AG03, N05AH01, N05AH02, N05AH03, N05AH04, N05AH05, N05AH06, N05AK01, N05AL01, N05AL02, N05AL03, N05AL04, N05AL05, N05AL06, N05AL07, N05AN01, N05AX07, N05AX08, N05AX09, N05AX10, N05AX11, N05AX12, N05AX13, N05AX14, N05AX15, N05BA01, N05BA02, N05BA03, N05BA04, N05BA05, N05BA06, N05BA07, N05BA08, N05BA09, N05BA10, N05BA11, N05BA12, N05BA13, N05BA14, N05BA15, N05BA16, N05BA17, N05BA18, N05BA19, N05BA21, N05BA22, N05BA23, N05BA56, N05BB01, N05BB02, N05BB51, N05BC01, N05BC03, N05BC04, N05BC51, N05BD01, N05BE01, N05BX01, N05BX02, N05BX03, N05BX05, N05CA01, N05CA02, N05CA03, N05CA04, N05CA05, N05CA06, N05CA07, N05CA08, N05CA09, N05CA10, N05CA11, N05CA12, N05CA15, N05CA16, N05CA19, N05CA20, N05CA21, N05CA22, N05CB01, N05CB02, N05CC01, N05CC02, N05CC03, N05CC04, N05CC05, N05CD01, N05CD02, N05CD03, N05CD04, N05CD05, N05CD06, N05CD07, N05CD08, N05CD09, N05CD10, N05CD11, N05CD12, N05CD13, N05CD14, N05CD15, N05CE01, N05CE02, N05CE03, N05CF01, N05CF02, N05CF03, N05CF04, N05CH01, N05CH02, N05CM01, N05CM02, N05CM03, N05CM04, N05CM05, N05CM06, N05CM07, N05CM08, |

| | |
|---|---|
| | N05CM09, N05CM10, N05CM11, N05CM12, N05CM13, N05CM15, N05CM16, N05CM17, N05CM18, N05CM19, N05CX01, N05CX02, N05CX03, N05CX04, N05CX05, N05CX06, N06AA01, N06AA02, N06AA03, N06AA04, N06AA05, N06AA06, N06AA07, N06AA08, N06AA09, N06AA10, N06AA11, N06AA12, N06AA13, N06AA14, N06AA15, N06AA16, N06AA17, N06AA18, N06AA19, N06AA21, N06AA23, N06AB02, N06AB03, N06AB04, N06AB05, N06AB06, N06AB07, N06AB08, N06AB09, N06AB10, N06AF01, N06AF02, N06AF03, N06AF04, N06AF05, N06AF06, N06AG02, N06AG03, N06AX01, N06AX02, N06AX03, N06AX04, N06AX05, N06AX06, N06AX07, N06AX08, N06AX09, N06AX10, N06AX11, N06AX12, N06AX13, N06AX14, N06AX15, N06AX16, N06AX17, N06AX18, N06AX19, N06AX21, N06AX22, N06AX23, N06AX24, N06AX25, N06AX26, N06AX27, N06BA01, N06BA02, N06BA03, N06BA04, N06BA05, N06BA06, N06BA07, N06BA08, N06BA09, N06BA10, N06BA11, N06BA12, N06BA14, N06BC01, N06BC02, N06BX01, N06BX02, N06BX03, N06BX04, N06BX05, N06BX06, N06BX07, N06BX08, N06BX09, N06BX10, N06BX11, N06BX12, N06BX13, N06BX14, N06BX15, N06BX16, N06BX17, N06BX18, N06BX21, N06CA01, N06CA02, N06DA01, N06DA02, N06DA03, N06DA04, N06DA52, N06DX01, N06DX02 |
| **Rhinological and throat antiseptics** | R01AA02, R01AA03, R01AA04, R01AA05, R01AA06, R01AA07, R01AA08, R01AA09, R01AA10, R01AA11, R01AA12, R01AA13, R01AA14, R01AB01, R01AB02, R01AB03, R01AB05, R01AB06, R01AB07, R01AB08, R01AC01, R01AC02, R01AC03, R01AC04, R01AC05, R01AC06, R01AC07, R01AC08, R01AC51, R01AD01, R01AD02, R01AD03, R01AD04, R01AD05, R01AD06, R01AD07, R01AD08, R01AD09, R01AD11, R01AD12, R01AD13, R01AD52, R01AD53, R01AD57, R01AD58, R01AD60, R01AX01, R01AX02, R01AX03, R01AX05, R01AX06, R01AX07, R01AX08, R01AX09, R01AX10, R01AX30, R01BA01, R01BA02, R01BA03, R01BA51, R01BA52, R01BA53, R02AA01, R02AA02, R02AA03, R02AA05, R02AA06, R02AA09, R02AA10, R02AA11, R02AA12, R02AA13, R02AA14, R02AA15, R02AA16, R02AA17, R02AA18, R02AA19, R02AA20, R02AB01, R02AB02, R02AB03, R02AB04, R02AB30, R02AD01, R02AD02, R02AD03, R02AD04, R02AX01, R02AX03 |
| **Inhaled anti-obstructive drugs (MG18)** | R03AA01, R03AB02, R03AB03, R03AC02, R03AC03, R03AC04, R03AC05, R03AC06, R03AC07, R03AC08, R03AC09, R03AC10, R03AC11, R03AC12, R03AC13, R03AC14, R03AC15, R03AC16, R03AC17, R03AC18, R03AC19, R03AK01, R03AK02, R03AK03, R03AK04, R03AK05, R03AK06, R03AK07, R03AK08, R03AK09, R03AK10, R03AK11, R03AK13, R03AK14, R03AL01, R03AL02, R03AL03, R03AL04, R03AL05, R03AL06, R03AL07, R03AL08, R03AL09, R03AL10, R03AL11, R03AL12, R03BB01, R03BB02, R03BB03, R03BB04, R03BB05, R03BB06, R03BB07, R03BB08, R03BC01, R03BC03, R03BX01, R03CA02, R03CB01, R03CB02, R03CB03, R03CB51, R03CB53, R03CC02, R03CC03, R03CC04, R03CC05, R03CC06, R03CC07, R03CC08, R03CC09, R03CC10, R03CC11, R03CC12, R03CC13, R03CC14, R03CC53, R03CC63, R03DA01, R03DA02, R03DA03, R03DA04, R03DA05, R03DA06, R03DA07, R03DA08, R03DA09, R03DA10, R03DA11, R03DA12, R03DA20, R03DA51, R03DA54, R03DA55, R03DA57, R03DA74, R03DB01, R03DB02, R03DB03, R03DB04, R03DB05, R03DB06 |
| **Inhaled steroids (MG19)** | R03BA01, R03BA02, R03BA03, R03BA04, R03BA05, R03BA06, R03BA07, R03BA08, R03BA09 |
| **Other COPD drugs** | R03DC01, R03DC02, R03DC03, R03DC04, R03DX01, R03DX02, R03DX03, R03DX05, R03DX06, R03DX07, R03DX08, R03DX09, R03DX10 |
| **Cold and cough preparations** | R05CA01, R05CA02, R05CA03, R05CA04, R05CA05, R05CA06, R05CA07, R05CA08, R05CA09, R05CA10, R05CA11, R05CA12, R05CA13, R05CB01, R05CB02, R05CB03, R05CB04, R05CB05, R05CB06, R05CB07, R05CB08, R05CB09, R05CB10, R05CB11, R05CB12, R05CB13, R05CB14, R05CB15, R05CB16, R05DA01, R05DA03, R05DA04, R05DA05, R05DA06, R05DA07, R05DA08, R05DA09, R05DA10, R05DA11, R05DA12, R05DA20, R05DB01, R05DB02, R05DB03, R05DB04, R05DB05, R05DB07, R05DB09, R05DB10, R05DB11, R05DB12, R05DB13, R05DB14, R05DB15, R05DB16, R05DB17, R05DB18, R05DB19, R05DB20, R05DB21, R05DB22, R05DB23, R05DB24, R05DB25, R05DB26, R05DB27, R05DB28, R05FA01, R05FA02, R05FB01, R05FB02 |
| **Systemic antihistamines (MG20)** | R06AA01, R06AA02, R06AA04, R06AA06, R06AA07, R06AA08, R06AA09, R06AA52, R06AA54, R06AA56, R06AA57, R06AA59, R06AB01, R06AB02, R06AB03, R06AB04, R06AB05, R06AB06, R06AB07, R06AB51, R06AB52, R06AB54, R06AB56, R06AC01, R06AC02, R06AC03, R06AC04, R06AC05, R06AC06, R06AC52, R06AC53, R06AD01, R06AD02, R06AD03, R06AD04, R06AD05, R06AD06, R06AD07, R06AD08, R06AD09, R06AD52, R06AD55, R06AE01, R06AE03, R06AE04, R06AE05, R06AE06, R06AE07, R06AE09, R06AE51, R06AE53, R06AE55, R06AX01, R06AX02, R06AX03, R06AX04, R06AX05, R06AX07, R06AX08, R06AX09, R06AX11, R06AX12, R06AX13, R06AX15, R06AX16, R06AX17, R06AX18, R06AX19, R06AX21, R06AX22, R06AX23, R06AX24, R06AX25, R06AX26, R06AX27, R06AX28, R06AX29, R06AX53, R06AX58 |
| **H2-blocker** | A02BA01, A02BA02, A02BA03, A02BA04, A02BA05, A02BA06, A02BA07, A02BA08, A02BA51, A02BA53 |

### Detailed Statistical Analysis Plan

For each patient, age, sex, region (Vienna, Lower Austria, Upper Austria, Burgenland, Carinthia, Salzburg, Tyrol, Styria, or Vorarlberg), and ATC codes describing prescribed medications were available from the Austrian Health Insurance Funds from 1 year before the hospitalization due to COVID-19. Due to the known effect of age on COVID-19 outcomes, all analyses were performed separately for four age subgroups: 19-40 years, 41-64 years, 65-74 years, ≥75 years (Table S2). ATC codes for medications before hospitalization were summarized in medication groups (Table S3).
A binary variable was defined for each medication group, which was set to 1 if a drug of the corresponding ICD10-codes was prescribed at least once 1 year before the index COVID-19 hospitalization and 0 if no drug was prescribed. Twenty medication groups were used for statistical modeling (MG1: Anticoagulants, MG2: Antibiotics, antivirals, antiprotozoals, or anthelmintics, MG3: Insulin and other antidiabetics, MG4: Heart drugs, MG5: Antihypertensives, including diuretics and renin-angiotensin-aldosterone system inhibitors, MG6: Beta-blockers, MG7: Statins, fibrates, including proprotein convertase subtilisin/kexin type 9 inhibitors and inclisiran, MG8: Immunosuppressants and immunomodulators, MG9: Systemic steroids, MG10: Chemotherapy, MG11: Iron supplements, erythropoietic stimulating agents, vitamin B12, folic acid, MG12: Antacids, including antihistamines, MG13: Vitamin D and other vitamin supplements, MG 14: NSAID and other anti-inflammatory drugs, MG15: Gout medications, MG16: Antiepileptics, MG17: Antipsychotics, MG18: Inhaled anti-obstructive drugs, MG19: Inhaled steroids, MG20: Systemic antihistamines). For controls, a similar medication profile was generated using the drugs prescribed 1 year before the index COVID-hospital stay of the matched patient. These 20 medication groups were also used to define polypharmacy.

**Descriptive statistics**
Numbers and percentages were used to summarize categorical variables, medians and interquartile ranges for continuous variables.

**Statistical analyses within patients**
To evaluate the association between polypharmacy and all-cause death, a simple Cox regression model was calculated accounting for sex, age, half-year (1st and 2nd half of 2020, indicating the two waves), and polypharmacy with clustering variable region. Polypharmacy was defined as the number of medication groups in which a patient received prescribed medication, categorized into four groups (0-1, 2-5, 6-10, >11). A similar analysis was performed for the subgroup of patients surviving the hospital stay. For these subgroups, the length of hospital stay was included as a co-variable.
To evaluate the association between polypharmacy and re-hospitalization due to any reason, competing risk models (with competing risk death using the Fine and Grey method) were calculated accounting for sex, age, half-year, and polypharmacy with clustering variable region.
To evaluate the association between several medication groups and all-cause death, a Cox regression model was calculated accounting for sex, age, half-year, polypharmacy, and the 20 medication groups with clustering variable region. Due to the low number of events in the 19-40 years age group and the underrepresentation of some medication groups, the model in this age group accounted for a smaller number of medication groups. A similar analysis was performed for the subgroup of patients surviving the COVID-hospital stay.
For re-hospitalization, a competing risk model (with competing risk death using the Fine and Grey method) was calculated using the same co-variables as described in the model for all-cause death.
Only 20 of the 32 medication groups were evaluated for the statistical models due to the underrepresentation of some medicament groups, with a prevalence over all age groups of <3% of the study population. For a detailed definition of medication groups based on the corresponding ATC codes, see Table S3.

**Statistical analyses comparing patients and controls**
As patients hospitalized due to COVID-19 had potentially more serious co-morbidities compared to controls ("normal Austrian population"), we attempted to account for this imbalance using propensity score matching. Patients and controls were matched for age, sex, region, and medication groups MG1 to MG20. To evaluate the difference between patients and controls in the risk of all-cause death, a Cox regression model was calculated accounting for group, sex, age, half-year, polypharmacy, and the 20 medication groups with clustering variable region.
A similar analysis was performed for the subgroup of patients surviving the COVID-hospital stay. For hospitalization due to any reason, a competing risk model (with competing risk death using the Fine and Gray method) was calculated using the same co-variables as described in the model for all-cause death.
Schönfeld residuals were used to evaluate the proportional hazard assumption and variance inflation factors to evaluate multicollinearity. Due to the large sample size and large number of investigated co-variables (i.e., 25), p<0,002 (=0,05/25 applying conservative Bonferroni correction) was considered significant. All analyses were performed using R, release 4.2.2.

### Comparison of medication groups between cohorts

**Tables S4 to S7** show the numbers and percentages of patients receiving at least one medication of the corresponding medication group, separately for COVID-hospitalized patients and age-, sex- and region- matched controls, and separately for the four age groups. The corresponding observed 1-year mortality rates ("1-year in %") are also given.

**Table S4: Medication groups for COVID-hospitalized patients and controls aged 19-40 years**

| | COVID | | | | Control | | | |
|---|---|---|---|---|---|---|---|---|
| | No: n (%) | 1-year % | Yes: n (%) | 1-year % | No: n (%) | 1-year % | Yes: n (%) | 1-year % |
| **Anticoagulants** | 1 128 (93,9) | 1,2 | 73 (6,1) | 4,1 | 11 788 (98,6) | 0,1 | 170 (1,4) | 1,8 |
| **Antibiotics, antivirals, antiprotozoals, or anthelmintics** | 638 (53,1) | 1,3 | 563 (46,9) | 1,6 | 10 076 (84,3) | 0,1 | 1 882 (15,7) | 0,2 |
| **Insulin and other antidiabetics** | 1 164 (96,9) | 1,3 | 37 (3,1) | 5,4 | 11 918 (99,7) | 0,1 | 40 (0,3) | 0,0 |
| **Heart drugs** | 1 180 (98,2) | 1,4 | 21 (1,8) | 0,0 | 11 907 (99,6) | 0,1 | 51 (0,4) | 2,0 |
| **Antihypertensives, incl. diuretics and renin-angiotensin-aldosterone system inhibitors** | 1 125 (93,7) | 1,2 | 76 (6,3) | 4,0 | 11 876 (99,1) | 0,1 | 82 (0,7) | 3,7 |
| **Beta-blockers** | 1 163 (96,8) | 1,3 | 38 (3,2) | 5,3 | 11 926 (99,3) | 0,1 | 32 (0,3) | 6,3 |
| **Statins, fibrates, incl. proprotein convertase subtilisin/kexin type 9 inhibitors and inclisiran** | 1 174 (97,8) | 1,3 | 27 (2,2) | 7,4 | 11 918 (99,7) | 0,1 | 40 (0,3) | 0,0 |
| **Immunosuppressants and immunomodulators** | 1 165 (97,0) | 1,4 | 36 (3,0) | 2,8 | 11 920 (99,7) | 0,1 | 38 (0,3) | 0,0 |
| **Systemic steroids** | 1 117 (93,0) | 1,2 | 84 (7,0) | 4,8 | 11 819 (98,8) | 0,1 | 139 (1,2) | 0,0 |
| **Chemotherapy** | 1 193 (99,3) | 1,4 | 8 (0,7) | 0,0 | 11 953 (99,9) | 0,1 | 5 (0,1) | 20,0 |
| **Iron supplements, erythropoietic stimulating agents, vitamin B12, folic acid** | 1 127 (93,8) | 1,3 | 74 (6,2) | 2,7 | 11 832 (99,0) | 0,1 | 126 (1,0) | 0,0 |
| **Antacids, incl. antihistamines** | 1 071 (89,2) | 1,2 | 130 (10,8) | 3,1 | 11 749 (98,3) | 0,1 | 209 (1,7) | 1,4 |
| **Vitamin D and other vitamin supplements** | 1 096 (91,3) | 1,5 | 105 (8,7) | 1,0 | 11 819 (98,8) | 0,1 | 139 (1,2) | 2,2 |
| **Caplacizumab** | 1 200 (99,9) | 1,4 | 1 (0,1) | 0,0 | 11 958 (100,0) | 0,1 | 0 (0,0) | |
| **Systemic hemostatics** | 1 201 (100,0) | 1,4 | 0 (0,0) | | 11 956 (100,0) | 0,1 | 2 (0,0) | 0,0 |
| **Hereditary angioedema therapeutics** | 1 201 (100,0) | 1,4 | 0 (0,0) | | 11 958 (100,0) | 0,1 | 0 (0,0) | |
| **Peripheral vasodilators** | 1 199 (99,8) | 1,4 | 2 (0,2) | 0,0 | 11 952 (99,9) | 0,1 | 6 (0,1) | 0,0 |
| **Hormonal contraceptives and similar hormone preparations** | 1 146 (95,4) | 1,5 | 55 (4,6) | 0,0 | 11 801 (98,7) | 0,1 | 157 (1,3) | 0,0 |
| **Immunoglobulins** | 1 199 (99,8) | 1,4 | 2 (0,2) | 0,0 | 11 958 (100,0) | 0,1 | 0 (0,0) | |
| **Interferons and CSF** | 1 184 (98,6) | 1,4 | 17 (1,4) | 5,9 | 11 909 (99,6) | 0,1 | 49 (0,4) | 2,4 |
| **NSAID and other anti-inflammatory drugs** | 885 (73,7) | 1,4 | 316 (26,3) | 1,6 | 11 084 (92,7) | 0,1 | 874 (7,3) | 0,1 |
| **Gout medications** | 1 193 (99,3) | 1,3 | 8 (0,7) | 12,5 | 11 953 (99,9) | 0,1 | 5 (0,1) | 0,0 |
| **Antiepileptics** | 1 127 (93,8) | 1,1 | 74 (6,2) | 6,8 | 11 873 (99,3) | 0,1 | 85 (0,7) | 1,2 |
| **Antipsychotics** | 1 029 (85,7) | 1,1 | 172 (14,3) | 3,5 | 11 550 (96,6) | 0,1 | 408 (3,4) | 1,5 |
| **Rhinological and throat antiseptics** | 1 075 (89,5) | 1,4 | 126 (10,5) | 1,6 | 11 584 (96,9) | 0,1 | 374 (3,1) | 0,3 |
| **Inhaled anti-obstructive drugs** | 1 075 (89,5) | 1,3 | 126 (10,5) | 2,4 | 11 709 (97,9) | 0,1 | 249 (2,1) | 0,4 |
| **Inhaled steroids** | 1 168 (97,3) | 1,5 | 33 (2,8) | 0,0 | 11 884 (99,4) | 0,1 | 74 (0,6) | 0,0 |
| **Other COPD drugs** | 1 190 (99,1) | 1,4 | 11 (0,9) | 0,0 | 11 927 (99,7) | 0,1 | 31 (0,3) | 0,0 |
| **Cold and cough preparations** | 1 114 (92,8) | 1,4 | 87 (7,2) | 2,3 | 11 847 (99,1) | 0,1 | 111 (0,9) | 0,9 |
| **Systemic antihistamines** | 1 135 (94,5) | 1,3 | 66 (5,5) | 3,0 | 11 809 (98,8) | 0,1 | 149 (1,2) | 0,0 |
| **H2-blocker** | 1 194 (99,4) | 1,4 | 7 (0,6) | 0,0 | 11 928 (99,8) | 0,1 | 30 (0,2) | 0,0 |

### Table S5: Medication groups for COVID-hospitalized patients and controls aged 41-64 years

| | COVID | | | | Control | | | |
|---|---|---|---|---|---|---|---|---|
| | **No: n (%)** | **1-year %** | **Yes: n (%)** | **1-year %** | **No: n (%)** | **1-year %** | **Yes: n (%)** | **1-year %** |
| **Anticoagulants** | 4 887 (81,2) | 6,0 | 1 131 (18,8) | 13,5 | 56 335 (95,4) | 0,7 | 2 722 (4,6) | 4,5 |
| **Antibiotics, antivirals, antiprotozoals, or anthelmintics** | 3 111 (51,7) | 6,0 | 2 907 (48,3) | 8,8 | 49 704 (84,2) | 0,7 | 9 353 (15,8) | 2,0 |
| **Insulin and other antidiabetics** | 5 134 (85,3) | 6,5 | 884 (14,7) | 12,3 | 57 568 (97,5) | 0,9 | 1 489 (2,5) | 3,4 |
| **Heart drugs** | 5 745 (95,5) | 7,2 | 273 (4,5) | 12,1 | 58 322 (98,8) | 0,9 | 735 (1,2) | 3,7 |
| **Antihypertensives, incl. diuretics and renin-angiotensin-aldosterone system inhibitors** | 3 998 (66,4) | 5,5 | 2 020 (33,6) | 11,1 | 53 940 (91,3) | 0,7 | 5 117 (8,7) | 2,8 |
| **Beta-blockers** | 5 154 (85,6) | 6,2 | 864 (14,4) | 14,2 | 57 089 (96,7) | 0,8 | 1 968 (3,3) | 4,7 |
| **Statins, fibrates, incl. proprotein convertase subtilisin/kexin type 9 inhibitors and inclisiran** | 4 616 (76,7) | 6,8 | 1 402 (23,3) | 9,2 | 55 178 (93,3) | 0,9 | 3 879 (6,6) | 1,8 |
| **Immunosuppressants and immunomodulators** | 5 786 (96,1) | 7,3 | 232 (3,9) | 9,9 | 58 646 (99,3) | 0,9 | 411 (0,7) | 2,7 |
| **Systemic steroids** | 5 275 (87,7) | 6,9 | 743 (12,4) | 10,9 | 57 063 (96,6) | 0,8 | 1 994 (3,4) | 4,4 |
| **Chemotherapy** | 5 908 (98,2) | 6,9 | 110 (1,8) | 30,9 | 58 815 (99,6) | 0,8 | 242 (0,4) | 17,8 |
| **Iron supplements, erythropoietic stimulating agents, vitamin B12, folic acid** | 5 700 (94,7) | 6,5 | 318 (5,3) | 23,0 | 58 518 (99,1) | 0,8 | 539 (0,9) | 10,0 |
| **Antacids, incl. antihistamines** | 4 790 (79,6) | 6,2 | 1 228 (20,4) | 11,9 | 56 677 (96,0) | 0,8 | 2 380 (4,0) | 4,6 |
| **Vitamin D and other vitamin supplements** | 5 239 (87,1) | 6,4 | 779 (12,9) | 13,7 | 57 257 (97,0) | 0,8 | 1 800 (3,0) | 4,3 |
| **Caplacizumab** | 6 018 (100,0) | 7,4 | 0 (0,0) | | 59 057 (100,0) | 0,9 | 0 (0,0) | |
| **Systemic hemostatics** | 6 016 (100,0) | 7,4 | 2 (0,0) | 50,0 | 59 055 (100,0) | 0,9 | 2 (0,0) | 0,0 |
| **Hereditary angioedema therapeutics** | 6 018 (100,0) | 7,4 | 0 (0,0) | | 59 057 (100,0) | 0,9 | 0 (0,0) | |
| **Peripheral vasodilators** | 5 985 (99,5) | 7,4 | 33 (0,6) | 6,1 | 58 923 (99,8) | 0,9 | 134 (0,2) | 0,0 |
| **Hormonal contraceptives and similar hormone preparations** | 5 812 (96,6) | 7,4 | 206 (3,4) | 6,8 | 57 857 (98,0) | 0,9 | 1 200 (2,0) | 0,6 |
| **Immunoglobulins** | 6 014 (99,9) | 7,4 | 4 (0,1) | 0,0 | 59 054 (100,0) | 0,9 | 3 (0,0) | 33,3 |
| **Interferons and CSF** | 5 873 (97,6) | 6,8 | 145 (2,4) | 29,0 | 58 739 (99,5) | 0,8 | 318 (0,5) | 14,5 |
| **NSAID and other anti-inflammatory drugs** | 3 819 (63,5) | 7,6 | 2 199 (36,5) | 7,0 | 51 259 (86,8) | 0,8 | 7 798 (13,2) | 1,7 |
| **Gout medications** | 5 842 (97,1) | 7,1 | 176 (2,9) | 15,9 | 58 684 (99,4) | 0,9 | 373 (0,6) | 3,2 |
| **Antiepileptics** | 5 459 (90,7) | 6,4 | 559 (9,3) | 17,4 | 57 921 (98,1) | 0,8 | 1 136 (1,9) | 7,0 |
| **Antipsychotics** | 4 564 (75,8) | 5,7 | 1 454 (24,2) | 12,6 | 54 970 (93,1) | 0,7 | 4 087 (6,9) | 3,4 |
| **Rhinological and throat antiseptics** | 5 416 (90,0) | 7,6 | 602 (10,0) | 5,5 | 57 332 (97,1) | 0,9 | 1 725 (2,9) | 1,8 |
| **Inhaled anti-obstructive drugs** | 4 936 (82,0) | 6,9 | 1 082 (18,0) | 9,7 | 56 563 (95,8) | 0,8 | 2 494 (4,2) | 3,4 |
| **Inhaled steroids** | 5 800 (96,4) | 7,4 | 218 (3,6) | 7,3 | 58 478 (99,0) | 0,9 | 579 (1,0) | 3,6 |
| **Other COPD drugs** | 5 900 (98,0) | 7,4 | 118 (2,0) | 5,9 | 58 864 (99,7) | 0,9 | 193 (0,3) | 3,6 |
| **Cold and cough preparations** | 5 460 (90,7) | 6,7 | 558 (9,3) | 11,5 | 58 215 (98,6) | 0,9 | 842 (1,4) | 5,3 |
| **Systemic antihistamines** | 5 655 (94,0) | 7,2 | 363 (6,0) | 10,5 | 58 171 (98,5) | 0,9 | 886 (1,5) | 3,1 |
| **H2-blocker** | 5 940 (98,7) | 7,3 | 78 (1,3) | 12,8 | 58 787 (99,5) | 0,9 | 270 (0,5) | 1,1 |

### Table S6: Medication groups for COVID-hospitalized patients and controls aged 65-74 years

| | Covid | | | | Control | | | |
|---|---|---|---|---|---|---|---|---|
| | No: n (%) | 1-year % | Yes: n (%) | 1-year % | No: n (%) | 1-year % | Yes: n (%) | 1-year % |
| **Anticoagulants** | 2 693 (59,8) | 17,2 | 1 809 (40,2) | 26,8 | 37 832 (87,0) | 2,3 | 5 679 (13,0) | 8,3 |
| **Antibiotics, antivirals, antiprotozoals, or anthelmintics** | 2 354 (52,3) | 19,6 | 2 148 (47,7) | 22,7 | 34 725 (79,8) | 2,4 | 8 786 (20,2) | 6,1 |
| **Insulin and other antidiabetics** | 3 311 (73,6) | 19,0 | 1 191 (26,5) | 26,8 | 40 339 (92,7) | 2,8 | 3 172 (7,3) | 7,8 |
| **Heart drugs** | 4 044 (89,8) | 20,5 | 458 (10,2) | 26,4 | 41 802 (96,1) | 2,9 | 1 709 (3,9) | 8,5 |
| **Antihypertensives, incl. diuretics and renin-angiotensin-aldosterone system inhibitors** | 1 774 (39,4) | 18,4 | 2 728 (60,6) | 22,8 | 33 384 (76,7) | 2,4 | 10 127 (23,3) | 5,7 |
| **Beta-blockers** | 3 104 (69,0) | 18,8 | 1 398 (31,0) | 26,1 | 38 925 (89,5) | 2,6 | 4 586 (10,5) | 7,6 |
| **Statins, fibrates, incl. proprotein convertase subtilisin/kexin type 9 inhibitors and inclisiran** | 2 576 (57,2) | 20,6 | 1 926 (42,8) | 21,7 | 35 557 (81,7) | 2,7 | 7 954 (18,3) | 4,9 |
| **Immunosuppressants and immunomodulators** | 4 293 (95,4) | 20,9 | 209 (4,6) | 24,4 | 43 026 (98,9) | 3,1 | 485 (1,1) | 5,6 |
| **Systemic steroids** | 3 746 (83,2) | 19,6 | 756 (16,8) | 28,3 | 40 818 (93,8) | 2,7 | 2 693 (6,2) | 9,2 |
| **Chemotherapy** | 4 342 (96,5) | 20,3 | 160 (3,5) | 41,9 | 42 928 (98,7) | 2,9 | 583 (1,3) | 20,4 |
| **Iron supplements, erythropoietic stimulating agents, vitamin B12, folic acid** | 4 090 (90,9) | 19,7 | 412 (9,1) | 35,0 | 42 654 (98,0) | 2,9 | 857 (2,0) | 17,0 |
| **Antacids, incl. antihistamines** | 3 014 (67,0) | 18,4 | 1 488 (33,0) | 26,6 | 39 566 (90,9) | 2,6 | 3 945 (9,1) | 8,1 |
| **Vitamin D and other vitamin supplements** | 3 606 (80,1) | 19,6 | 896 (19,9) | 27,1 | 40 597 (93,3) | 2,8 | 2 914 (6,7) | 7,3 |
| **Caplacizumab** | 4 502 (100,0) | 21,1 | 0 (0,0) | | 43 511 (100,0) | 3,1 | 0 (0,0) | |
| **Systemic hemostatics** | 4 500 (100,0) | 21,1 | 2 (0,0) | 50,0 | 43 509 (100,0) | 3,1 | 2 (0,0) | 0,0 |
| **Hereditary angioedema therapeutics** | 4 502 (100,0) | 21,1 | 0 (0,0) | | 43 511 (100,0) | 3,1 | 0 (0,0) | |
| **Peripheral vasodilators** | 4 451 (98,9) | 21,0 | 51 (1,1) | 27,5 | 43 308 (99,5) | 3,1 | 203 (0,5) | 4,4 |
| **Hormonal contraceptives and similar hormone preparations** | 4 361 (96,9) | 21,4 | 141 (3,1) | 12,1 | 42 285 (97,2) | 3,2 | 1 226 (2,8) | 1,9 |
| **Immunoglobulins** | 4 498 (99,9) | 21,1 | 4 (0,1) | 25,0 | 43 507 (100,0) | 3,1 | 4 (0,0) | 0,0 |
| **Interferons and CSF** | 4 372 (97,1) | 20,6 | 130 (2,9) | 37,7 | 43 113 (99,1) | 3,0 | 398 (0,9) | 22,9 |
| **NSAID and other anti-inflammatory drugs** | 2 949 (65,5) | 23,1 | 1 553 (34,5) | 17,2 | 35 651 (81,9) | 2,9 | 7 860 (18,1) | 4,2 |
| **Gout medications** | 4 193 (93,1) | 20,1 | 309 (6,9) | 35,0 | 42 763 (98,3) | 3,0 | 748 (1,7) | 10,0 |
| **Antiepileptics** | 3 849 (85,5) | 18,8 | 653 (14,5) | 34,3 | 41 953 (96,4) | 2,8 | 1 558 (3,6) | 12,1 |
| **Antipsychotics** | 2 837 (63,0) | 17,2 | 1 665 (37,0) | 27,6 | 38 017 (87,4) | 2,5 | 5 494 (12,6) | 7,7 |
| **Rhinological and throat antiseptics** | 4 147 (92,1) | 21,0 | 355 (7,9) | 22,0 | 42 021 (96,6) | 3,1 | 1 490 (3,4) | 3,3 |
| **Inhaled anti-obstructive drugs** | 3 357 (74,6) | 19,3 | 1 145 (25,4) | 26,4 | 40 299 (92,6) | 2,7 | 3 212 (7,4) | 8,7 |
| **Inhaled steroids** | 4 304 (95,6) | 21,1 | 198 (4,4) | 21,2 | 42 858 (98,5) | 3,1 | 653 (1,5) | 6,0 |
| **Other COPD drugs** | 4 419 (98,2) | 21,1 | 83 (1,8) | 22,9 | 43 331 (99,6) | 3,1 | 180 (0,4) | 7,2 |
| **Cold and cough preparations** | 3 906 (86,8) | 20,3 | 596 (13,2) | 26,0 | 42 178 (96,9) | 2,9 | 1 333 (3,1) | 9,3 |
| **Systemic antihistamines** | 4 205 (93,4) | 21,0 | 297 (6,6) | 22,6 | 42 491 (97,7) | 3,1 | 1 020 (2,3) | 5,9 |
| **H2-blocker** | 4 430 (98,4) | 21,1 | 72 (1,6) | 22,2 | 43 147 (99,2) | 3,1 | 364 (0,8) | 7,7 |

**Table S7: Medication groups for COVID-hospitalized patients and controls aged ≥75 years**

| | Covid | | | | Control | | | |
|---|---|---|---|---|---|---|---|---|
| | No: n (%) | 1-year % | Yes: n (%) | 1-year % | No: n (%) | 1-year % | Yes: n (%) | 1-year % |
| **Anticoagulants** | 4 669 (43,0) | 37,8 | 6 181 (57,0) | 46,6 | 72 935 (71,0) | 8,58 | 29 834 (29,0) | 23,5 |
| **Antibiotics, antivirals, antiprotozoals, or anthelmintics** | 5 795 (53,4) | 39,2 | 5 055 (46,6) | 47,0 | 73 958 (72,0) | 9,3 | 28 811 (28,0) | 22,2 |
| **Insulin and other antidiabetics** | 8 518 (78,5) | 42,5 | 2 332 (21,5) | 44,1 | 93 203 (90,7) | 11,82 | 9 566 (9,3) | 23,4 |
| **Heart drugs** | 9 060 (83,5) | 41,7 | 1 790 (16,5) | 48,3 | 92 443 (90,0) | 11,8 | 10 326 (10,0) | 23,2 |
| **Antihypertensives, incl. diuretics and renin-angiotensin-aldosterone system inhibitors** | 3 375 (31,1) | 42,0 | 7 475 (68,9) | 43,2 | 62 314 (60,6) | 8,5 | 40 455 (39,4) | 19,2 |
| **Beta-blockers** | 6 966 (64,2) | 41,5 | 3 884 (35,8) | 45,2 | 82 712 (80,5) | 11,0 | 20 057 (19,5) | 20,7 |
| **Statins, fibrates, incl. proprotein convertase subtilisin/kexin type 9 inhibitors and inclisiran** | 6 667 (61,5) | 45,1 | 4 183 (38,5) | 39,2 | 77 798 (75,7) | 12,2 | 24 971 (24,3) | 15,0 |
| **Immunosuppressants and immunomodulators** | 10 617 (97,9) | 42,9 | 233 (2,1) | 40,3 | 101 723 (99,0) | 12,9 | 1 046 (1,0) | 15,4 |
| **Systemic steroids** | 9 302 (85,7) | 42,7 | 1 548 (14,3) | 43,8 | 94 167 (91,6) | 12,4 | 8 602 (8,4) | 18,7 |
| **Chemotherapy** | 10 420 (96,0) | 42,5 | 430 (4,0) | 49,5 | 100 310 (97,6) | 12,5 | 2 459 (2,4) | 28,8 |
| **Iron supplements, erythropoietic stimulating agents, vitamin B12, folic acid** | 9 439 (87,0) | 41,0 | 1 411 (13,0) | 54,9 | 97 255 (94,6) | 11,7 | 5 514 (5,4) | 34,6 |
| **Antacids, incl. antihistamines** | 6 954 (64,1) | 40,6 | 3 896 (35,9) | 46,8 | 86 259 (83,9) | 10,7 | 16 510 (16,1) | 24,4 |
| **Vitamin D and other vitamin supplements** | 8 210 (75,7) | 41,8 | 2 640 (24,3) | 46,0 | 89 520 (87,1) | 11,6 | 13 249 (12,9) | 22,0 |
| **Caplacizumab** | 10 850 (100,0) | 42,8 | 0 (0,0) | | 102 769 (100,0) | 12,9 | 0 (0,0) | |
| **Systemic hemostatics** | 10 846 (100,0) | 42,8 | 4 (0,0) | 50,0 | 102 750 (100,0) | 12,9 | 19 (0,0) | 15,8 |
| **Hereditary angioedema therapeutics** | 10 850 (100,0) | 42,8 | 0 (0,0) | | 102 769 (100,0) | 12,9 | 0 (0,0) | |
| **Peripheral vasodilators** | 10 709 (98,7) | 42,9 | 141 (1,3) | 38,3 | 101 638 (98,9) | 12,8 | 1 131 (1,1) | 20,4 |
| **Hormonal contraceptives and similar hormone preparations** | 10 588 (97,6) | 43,1 | 262 (2,4) | 30,9 | 99 881 (97,2) | 13,0 | 2 888 (2,8) | 10,0 |
| **Immunoglobulins** | 10 842 (99,9) | 42,8 | 8 (0,1) | 100,0 | 102 754 (100,0) | 12,9 | 15 (0,0) | 20,0 |
| **Interferons and CSF** | 10 722 (98,8) | 42,7 | 128 (1,2) | 49,2 | 101 907 (99,2) | 12,8 | 862 (0,8) | 23,8 |
| **NSAID and other anti-inflammatory drugs** | 8 159 (75,2) | 45,4 | 2 691 (24,8) | 35,0 | 82 029 (79,8) | 12,7 | 20 740 (20,2) | 13,8 |
| **Gout medications** | 9 940 (91,6) | 42,2 | 910 (8,4) | 49,9 | 98 922 (96,3) | 12,4 | 3 847 (3,7) | 26,5 |
| **Antiepileptics** | 9 117 (84,0) | 41,9 | 1 733 (16,0) | 47,9 | 95 693 (93,1) | 12,1 | 7 076 (6,9) | 23,8 |
| **Antipsychotics** | 4 820 (44,4) | 35,2 | 6 030 (55,6) | 48,9 | 74 444 (72,4) | 8,3 | 28 325 (27,6) | 25,1 |
| **Rhinological and throat antiseptics** | 10 421 (96,1) | 43,1 | 429 (3,9) | 35,0 | 99 279 (96,6) | 12,9 | 3 490 (3,4) | 13,9 |
| **Inhaled anti-obstructive drugs** | 8 741 (80,6) | 42,2 | 2 109 (19,4) | 45,6 | 93 013 (90,5) | 11,9 | 9 756 (9,5) | 22,2 |
| **Inhaled steroids** | 10 572 (97,4) | 42,9 | 278 (2,6) | 41,4 | 101 127 (98,4) | 12,8 | 1 642´(1,6) | 16,9 |
| **Other COPD drugs** | 10 755 (99,1) | 42,9 | 95 (0,9) | 39,0 | 102 370 (99,6) | 12,9 | 399 (0,4) | 16,0 |
| **Cold and cough preparations** | 9 478 (87,4) | 42,5 | 1 372 (12,6) | 45,0 | 96 995 (94,4) | 12,0 | 5 774 (5,6) | 26,9 |
| **Systemic antihistamines** | 10 099 (93,1) | 42,6 | 751 (6,9) | 46,1 | 98 893 (96,2) | 12,5 | 3 876 (3,8) | 23,4 |
| **H2-blocker** | 10 729 (98,9) | 42,9 | 121 (1,1) | 38,8 | 101 646 (98,9) | 12,9 | 1 123 (1,1) | 17,9 |

### Survival probabilities

**Table S8** shows the hospital mortality and 30-day, 180-day, 1-year, and 1,5-year mortality rates for COVID-hospitalized patients and age-, sex- and region-matched controls separately for the four age groups and according to sex. Numbers are given for the overall patient and control cohorts, as well as for the propensity score-matched patient and control cohorts.

**Table S9** shows hospital mortality and 30-day, 180-day, 1-year, and 1,5-year mortality rates for COVID-hospitalized patients surviving the hospital stay and age-, sex- and region-matched controls separately for the four age groups and according to sex. Numbers are given for the overall patient and control cohorts, as well as for the propensity score-matched patient and control cohorts.
Note that, for controls, "surviving hospital stay" means that the control did not die within the duration of the hospital stay of the corresponding age-, sex- and region-matched patient.

**Table S8: Mortality for all patients and controls before and after propensity score matching (PSM)**

| | | | All patients and controls | | | | Patients and controls after PSM | | | |
|---|---|---|---|---|---|---|---|---|---|---|
| | | | COVID (all) | | Controll (all) | | COVID (PSM) | | Control (PSM) | |
| | | | Alive | Dead | Alive | Dead | Alive | Dead | Alive | Dead |
| Sex | Age | Time | n (%) | n (%) | n (%) | n (%) | n (%) | n (%) | n (%) | n (%) |
| Male | 19-40 | Hospital | 658 (99,0) | 7 (1,0) | 6 615 (100,0) | 1 (0,0) | 505 (99,2) | 4 (0,8) | 949 (99,9) | 1 (0,1) |
| | | 30-day | 658 (99,0) | 7 (1,0) | 6 614 (100,0) | 2 (0,0) | 506 (99,4) | 3 (0,6) | 949 (99,9) | 1 (0,1) |
| | | 180-day | 653 (98,2) | 12 (1,8) | 6 609 (99,9) | 7 (0,1) | 504 (99,0) | 5 (1,0) | 948 (99,8) | 2 (0,2) |
| | | 1-year | 653 (98,2) | 12 (1,8) | 6 608 (99,9) | 8 (0,1) | 504 (99,0) | 5 (1,0) | 948 (99,8) | 2 (0,2) |
| | | 1.5-year | 652 (98,1) | 13 (1,9) | 6 606 (99,9) | 10 (0,2) | 504 (99,0) | 5 (1,0) | 947 (99,7) | 3 (0,3) |
| | 41-64 | Hospital | 3 575 (94,9) | 194 (5,2) | 36 836 (99,9) | 34 (0,1) | 3 204 (95,3) | 157 (4,7) | 6 167 (99,9) | 9 (0,1) |
| | | 30-day | 3 599 (95,5) | 170 (4,5) | 36 790 (99,8) | 80 (0,2) | 3 225 (96,0) | 136 (4,0) | 6 152 (99,6) | 24 (0,4) |
| | | 180-day | 3 508 (93,1) | 261 (6,9) | 36 566 (99,2) | 304 (0,8) | 3 154 (93,8) | 207 (6,2) | 6 086 (98,5) | 90 (1,5) |
| | | 1-year | 3 471 (92,1) | 298 (7,9) | 36 473 (98,9) | 397 (1,1) | 3 122 (92,9) | 239 (7,1) | 6 058 (98,1) | 118 (1,9) |
| | | 1.5-year | 3 442 (91,3) | 327 (8,7) | 36 414 (98,8) | 456 (1,2) | 3 100 (92,2) | 261 (7,8) | 6 039 (97,8) | 137 (2,2) |
| | 65-74 | Hospital | 2 177 (82,3) | 467 (17,7) | 25 240 (99,6) | 113 (0,4) | 1 971 (83,4) | 393 (16,6) | 4 178 (99,4) | 27 (0,6) |
| | | 30-day | 2 199 (83,2) | 445 (16,8) | 25 168 (99,3) | 185 (0,7) | 1 992 (84,3) | 372 (15,7) | 4 161 (99,0) | 44 (1,0) |
| | | 180-day | 2 055 (77,7) | 589 (22,3) | 24 662 (97,3) | 691 (2,7) | 1 873 (79,2) | 491 (20,8) | 4 023 (95,7) | 182 (4,3) |
| | | 1-year | 1 973 (74,6) | 671 (25,4) | 24 426 (96,3) | 927 (3,7) | 1 806 (76,4) | 558 (23,6%) | 3 943 (93,8) | 262 (6,2) |
| | | 1.5-year | 1 914 (72,4) | 730 (27,6) | 24 253 (95,7) | 1 100 (4,3) | 1 762 (74,5) | 602 (25,5) | 3 884 (92,4) | 321 (7,6) |
| | ≥75 | Hospital | 3 318 (66,4) | 1 676 (33,6) | 46 324 (98,5) | 714 (1,5) | 3 256 (66,6) | 1 634 (33,4) | 8 546 (97,3) | 240 (2,7) |
| | | 30-day | 3 264 (65,4) | 1 730 (34,6) | 45 736 (97,2) | 1 302 (2,8) | 3 202 (65,5) | 1 688 (34,5) | 8 345 (95,0) | 441 (5,0) |
| | | 180-day | 2 859 (57,3) | 2 135 (42,8) | 42 971 (91,4) | 4 067 (8,6) | 2 810 (57,5) | 2 080 (42,5) | 7 464 (85,0) | 1322 (15,0) |
| | | 1-year | 2 687 (53,8) | 2 307 (46,2) | 41 454 (88,1) | 5 584 (11,9) | 2 641 (54,0) | 2 249 (46,0) | 6 953 (79,1) | 1833 (20,9) |
| | | 1.5-year | 2 516 (50,4) | 2478 (49,6) | 40 306 (85,7) | 6 732 (14,3) | 2 476 (50,6) | 2 414 (49,4) | 6 598 (75,1) | 2188 (24,9) |
| Female | 19-40 | Hospital | 534 (99,6) | 2 (0,4) | 5 342 (100,0) | 0 (0,0) | 409 (99,7) | 1 (0,3) | 735 (100,0) | 0 (0,0) |
| | | 30-day | 534 (99,6) | 2 (0,4) | 5 342 (100,0) | 0 (0,0) | 409 (99,8) | 1 (0,2) | 735 (100,0) | 0 (0,0) |
| | | 180-day | 533 (99,4) | 3 (0,6) | 5 339 (99,9) | 3 (0,1) | 409 (99,8) | 1 (0,2) | 735 (100,0) | 0 (0,0) |
| | | 1-year | 531 (99,1) | 5 (0,9) | 5 338 (99,9) | 4 (0,1) | 408 (99,5) | 2 (0,5) | 735 (100,0) | 0 (0,0) |
| | | 1.5-year | 531 (99,1) | 5 (0,9) | 5 336 (99,9) | 6 (0,1) | 408 (99,5) | 2 (0,5) | 735 (100,0) | 0 (0,0) |
| | 41-64 | Hospital | 2 152 (95,7) | 97 (4,3) | 22 171 (99,9) | 16 (0,1) | 1 919 (96,4) | 71 (3,6) | 3 710 (99,9) | 3 (0,1) |
| | | 30-day | 2 162 (96,1) | 87 (3,9) | 22 157 (99,9) | 30 (0,1) | 1 927 (96,8) | 63 (3,2) | 3 705 (99,8) | 8 (0,2) |
| | | 180-day | 2 120 (94,3) | 129 (5,7) | 22 079 (99,5) | 108 (0,5) | 1 894 (95,2) | 96 (4,8) | 3 674 (99,0) | 39 (1,0) |
| | | 1-year | 2 103 (93,5) | 146 (6,5) | 22 046 (99,4) | 141 (0,6) | 1 881 (94,5) | 109 (5,5) | 3 660 (98,6) | 53 (1,4) |
| | | 1.5-year | 2 090 (92,9) | 159 (7,1) | 22 022 (99,3) | 165 (0,7) | 1 871 (94,0) | 119 (6,0) | 3 650 (98,3) | 63 (1,7) |
| | 65-74 | Hospital | 1 677 (90,3) | 181 (9,7) | 18 109 (99,7) | 49 (0,3) | 1 542 (91,0) | 153 (9,0) | 3 235 (99,5) | 17 (0,5) |
| | | 30-day | 1 687 (90,8) | 171 (9,2) | 18 067 (99,5) | 91 (0,5) | 1 546 (91,2) | 149 (8,8) | 3 227 (99,2) | 25 (0,8) |
| | | 180-day | 1 608 (86,5) | 250 (13,5) | 17 834 (98,2) | 324 (1,8) | 1 481 (87,4) | 214 (12,6) | 3 148 (96,8) | 104 (3,2) |
| | | 1-year | 1 580 (85,0) | 278 (15,0) | 17 722 (97,6) | 436 (2,4) | 1 460 (86,1) | 235 (13,9) | 3 117 (95,9) | 135 (4,1) |
| | | 1.5-year | 1 542 (83,0) | 316 (17,0) | 17646 (97,2) | 512 (2,8) | 1 427 (84,2) | 268 (15,8) | 3 095 (95,2) | 157 (4,8) |
| | ≥75 | Hospital | 4 356 (74,4) | 1500 (25,6) | 54708 (98,2) | 1 023 (1,8) | 4 312 (74,5) | 1 478 (25,5) | 11 692 (97,1) | 353 (2,9) |
| | | 30-day | 4 260 (72,8) | 1596 (27,3) | 53916 (96,7) | 1 815 (3,3) | 4 217 (72,8) | 1 573 (27,2) | 11 405 (94,7) | 640 (5,3) |
| | | 180-day | 3 764 (64,3) | 2092 (35,7) | 50045 (89,8) | 5 686 (10,2) | 3 729 (64,4) | 2 061 (35,6) | 10 165 (84,4) | 1 880 (15,6) |
| | | 1-year | 3 517 (60,1) | 2339 (39,9) | 48056 (86,2) | 7 675 (13,8) | 3 485 (60,2) | 2 305 (39,8) | 9 515 (79,0) | 2 530 (21,0) |
| | | 1.5-year | 3 308 (56,5) | 2548 (43,5) | 46450 (83,3) | 9 281 (16,7) | 3 278 (56,6) | 2 512 (43,4) | 9 020 (74,9) | 3 025 (25,1) |

**Table S9: Mortality for all patients and controls surviving the hospital stay before and after propensity score matching (PSM)**

| | | | Patients surviving hospital | | | | Patients surviving hospital, after PSM | | | |
|---|---|---|---|---|---|---|---|---|---|---|
| | | | COVID (all) | | Controll (all) | | COVID (PSM) | | Control (PSM) | |
| | | | Alive | Dead | Alive | Dead | Alive | Dead | Alive | Dead |
| Sex | Age | Time | n (%) | n (%) | n (%) | n (%) | n (%) | n (%) | n (%) | n (%) |
| Male | 19-40 | 30-day | 656 (99,7) | 2 (0,3) | 6 614 (100,0) | 1 (0,0) | 506 (100,0) | 0 (0,0) | 915 (100,0) | 0 (0,0) |
| | | 180-day | 653 (99,2) | 5 (0,8) | 6 609 (99,9) | 6 (0,1) | 504 (99,6) | 2 (0,4) | 914 (99,9) | 1 (0,1) |
| | | 1-year | 653 (99,2) | 5 (0,8) | 6 608 (99,9) | 7 (0,1) | 504 (99,6) | 2 (0,4) | 914 (99,9) | 1 (0,1) |
| | | 1.5-year | 652 (99,1) | 6 (0,9) | 6 606 (99,9) | 9 (0,1) | 504 (99,6) | 2 (0,4) | 913 (99,8) | 2 (0,2) |
| | 41-64 | 30-day | 3 554 (99,4) | 21 (0,6) | 36 786 (99,9) | 50 (0,1) | 3 216 (99,5) | 16 (0,5) | 5 842 (99,7) | 16 (0,3) |
| | | 180-day | 3 508 (98,1) | 67 (1,9) | 36 566 (99,3) | 270 (0,7) | 3 178 (98,3) | 54 (1,7) | 5 788 (98,8) | 70 (1,2) |
| | | 1-year | 3 471 (97,1) | 104 (2,9) | 36 473 (99,0) | 363 (1,0) | 3 149 (97,4) | 83 (2,6) | 5 764 (98,4) | 94 (1,6) |
| | | 1.5-year | 3 442 (96,3) | 133 (3,7) | 36 414 (98,9) | 422 (1,1) | 3 128 (96,8) | 104 (3,2) | 5 749 (98,1) | 109 (1,9) |
| | 65-74 | 30-day | 2 136 (98,1) | 41 (1,9) | 25 155 (99,7) | 85 (0,3) | 1 966 (98,3) | 34 (1,7) | 3 644 (99,5) | 17 (0,5) |
| | | 180-day | 2 055 (94,4) | 122 (5,6) | 24 662 (97,7) | 578 (2,3) | 1 898 (94,9) | 102 (5,1) | 3 529 (96,4) | 132 (3,6) |
| | | 1-year | 1 973 (90,6) | 204 (9,4) | 24 426 (96,8) | 814 (3,2) | 1 830 (91,5) | 170 (8,5) | 3 464 (94,6) | 197 (5,4) |
| | | 1.5-year | 1 914 (87,9) | 263 (12,1) | 24 253 (96,1) | 987 (3,9) | 1 782 (89,1) | 218 (10,9) | 3 422 (93,5) | 239 (6,5) |
| | ≥75 | 30-day | 3 177 (95,8) | 141 (4,3) | 45 694 (98,6) | 630 (1,4) | 3 112 (95,8) | 136 (4,2) | 5 930 (97,7) | 143 (2,3) |
| | | 180-day | 2 859 (86,2) | 459 (13,8) | 42 971 (92,8) | 3 353 (7,2) | 2 803 (86,3) | 445 (13,7) | 5 329 (87,8) | 744 (12,2) |
| | | 1-year | 2 687 (81,0) | 631 (19,0) | 41 454 (89,5) | 4 870 (10,5) | 2 635 (81,1) | 613 (18,9) | 4 986 (82,1) | 1 087 (17,9) |
| | | 1.5-year | 2 516 (75,8) | 802 (24,2) | 40 306 (87,0) | 6 018 (13,0) | 2 468 (76,0) | 780 (24,0) | 4 758 (78,4) | 1 315 (21,6) |
| Female | 19-40 | 30-day | 534 (100,0) | 0 (0,0) | 5 342 (100,0) | 0 (0,0) | 387 (100,0) | 0 (0,0) | 735 (100,0) | 0 (0,0) |
| | | 180-day | 533 (99,8) | 1 (0,2) | 5 339 (99,9) | 3 (0,1) | 387 (100,0) | 0 (0,0) | 735 (100,0) | 0 (0,0) |
| | | 1-year | 531 (99,4) | 3 (0,6) | 5 338 (99,9) | 4 (0,1) | 387 (100,0) | 0 (0,0) | 735 (100,0) | 0 (0,0) |
| | | 1.5-year | 531 (99,4) | 3 (0,6) | 5 336 (99,9) | 6 (0,1) | 387 (100,0) | 0 (0,0) | 735 (100,0) | 0 (0,0) |
| | 41-64 | 30-day | 2 145 (99,7) | 7 (0,3) | 22 156 (99,9) | 15 (0,1) | 1 902 (99,7) | 5 (0,3 | 3 649 (99,9) | 4 (0,1) |
| | | 180-day | 2 120 (98,5) | 32 (1,5) | 22 079 (99,6) | 92 (0,4) | 1 884 (98,8) | 23 (1,2) | 3 627 (99,3) | 26 (0,7) |
| | | 1-year | 2 103 (97,7) | 49 (2,3) | 22 046 (99,4) | 125 (0,6) | 1 870 (98,1) | 37 (1,9) | 3 615 (99,0) | 38 (1,0) |
| | | 1.5-year | 2 090 (97,1) | 62 (2,9) | 22 022 (99,3) | 149 (0,7) | 1 862 (97,6) | 45 (2,4) | 3 610 (98,8) | 43 (1,2) |
| | 65-74 | 30-day | 1 664 (99,2) | 13 (0,8) | 18 062 (99,7) | 47 (0,3) | 1 509 (99,2) | 13 (0,8) | 2 920 (99,7) | 8 (0,3) |
| | | 180-day | 1 608 (95,9) | 69 (4,1) | 17 834 (98,5) | 275 (1,5) | 1 461 (96,0) | 61 (4,0) | 2 851 (97,4) | 77 (2,6) |
| | | 1-year | 1 580 (94,2) | 97 (5,8) | 17 722 (97,9) | 387 (2,1) | 1 440 (94,6) | 82 (5,4) | 2 825 (96,5) | 103 (3,5) |
| | | 1.5-year | 1 542 (91,9) | 135 (8,1) | 17 646 (97,4) | 463 (2,6) | 1 406 (92,4) | 116 (7,6) | 2 802 (95,7) | 126 (4,3) |
| | ≥75 | 30-day | 4 184 (96,0) | 172 (4,0) | 53 871 (98,5) | 837 (1,5) | 4 124 (96,0) | 172 (4,0) | 8 601 (97,6) | 210 (2,4) |
| | | 180-day | 3 764 (86,4) | 592 (13,6) | 50 045 (91,5) | 4 663 (8,5) | 3 710 (86,4) | 586 (13,6) | 7 736 (87,8) | 1 075 (12,2) |
| | | 1-year | 3 517 (80,7) | 839 (19,3) | 48 056 (87,8) | 6 652 (12,2) | 3 469 (80,8) | 827 (19,2) | 7 261 (82,4) | 1 550 (17,6) |
| | | 1.5-year | 3 308 (75,9) | 1 048 (24,1) | 46 450 (84,9) | 8 258 (15,1) | 3 262 (75,9) | 1 034 (24,1) | 6 880 (78,1) | 1 931 (21,9) |

### Analysis of the COVID patient cohort

**Table S10** shows the results of the simple Cox regression models including sex, age, half-year, and polypharmacy with clustering variable region (federal state). Note that the models for the subgroup of patients surviving the hospital stay additionally included the length of hospital stay. For the outcome hospitalization due to any reason after index COVID hospital stay, a competing risk model with competing risk death was calculated.

**Figure S1** shows Kaplan-Meier curves for COVID-hospitalized patients by age group separately for patients with 0 medication groups and at least one medication group. The corresponding Kaplan-Meier curves for all polypharmacy groups are shown in the manuscript in Figure 2.

**Table S11** shows the results of the multivariable Cox regression model including sex, age, half-year, polypharmacy, and several medicament groups with clustering variable region (federal state) for the outcome all-cause mortality. A graphical representation of the hazard rates and confidence intervals is shown in the manuscript in Figure 3.

**Table S12** shows the results of the multivariable Cox regression model including sex, age, half-year, polypharmacy, and several medicament groups with clustering variable region (federal state) for the outcome all-cause mortality for the subgroup of patients surviving the COVID-hospital stay. A graphical representation of the hazard rates and confidence intervals is shown in the manuscript in **Figure S2**.

**Table S13** shows the results of the competing risk model including sex, age, half-year, polypharmacy, and several medicament groups with clustering variable region (federal state) for the outcome hospitalization due to any reason after index COVID-hospital stay. A graphical representation of the hazard rates and confidence intervals is shown in the manuscript in **Figure S3**.

**Table S10: Results of simple models for all-cause mortality, all-cause mortality and re-hospitalization after COVID-hospital survival**

| All patients: all-cause mortality | | | | | | | | |
|---|---|---|---|---|---|---|---|---|
| | Age 19-40 years | | Age 41-64 years | | Age 65-74 years | | Age ≥75 years | |
| | HR (CI) | p-value | HR (CI) | p-value | HR (CI) | p-value | HR (CI) | p-value |
| **Sex: W (ref:M)** | 0,42 (0,20 - 0,88) | 0,022 | 0,76 (0,61 - 0,96) | 0,020 | 0,54 (0,48 - 0,62) | <0,001 | 0,72 (0,68 - 0,77) | <0,001 |
| **Age groups: AG1 (ref: AG-R)** | 0,56 (0,15 - 2,13) | 0,397 | 0,65 (0,51 - 0,82) | <0,001 | 0,72 (0,64 - 0,81) | <0,001 | 0,64 (0,58 - 0,70) | <0,001 |
| **AG2 (ref: AG-R)** | 0,77 (0,22 - 2,67) | 0,676 | 0,41 (0,34 - 0,49) | <0,001 | | | 0,42 (0,38 - 0,47) | <0,001 |
| **AG3 (ref: AG-R)** | 0,48 (0,13 - 1,76) | 0,265 | 0,34 (0,28 - 0,43) | <0,001 | | | 0,31 (0,27 - 0,34) | <0,001 |
| **AG4 (ref: AG-R)** | | | 0,19 (0,13 - 0,27) | <0,001 | | | | |
| **Half-year: (ref: 1. Half year)** | 3,20 (0,34 - 30,10) | 0,309 | 1,12 (0,74 - 1,68) | 0,603 | 0,73 (0,56 - 0,96) | 0,022 | 0,95 (0,83 - 1,10) | 0,520 |
| **Polypharmacy: 2-5 (ref: 0-1)** | 2,95 (0,87 - 10,00) | 0,083 | 1,69 (1,32 - 2,18) | <0,001 | 1,28 (1,06 - 1,55) | 0,010 | 1,16 (1,04 - 1,29) | 0,010 |
| **Polypharmacy: 6-10 (ref: 0-1)** | 8,79 (1,71 - 45,25) | 0,009 | 3,56 (2,92 - 4,33) | <0,001 | 2,18 (1,72 - 2,76) | <0,001 | 1,49 (1,31 - 1,69) | <0,001 |
| **Polypharmacy: ≥11 (ref: 0-1)** | NA | NA | 4,93 (3,36 - 7,23) | <0,001 | 3,06 (2,35 - 3,97) | <0,001 | 1,81 (1,51 - 2,18) | <0,001 |
| **Patients surviving index COVID hospital stay: all-cause mortality** | | | | | | | | |
| | Age 19-40 years | | Age 41-64 years | | Age 65-74 years | | Age ≥75 years | |
| | HR (CI) | p-value | HR (CI) | p-value | HR (CI) | p-value | HR (CI) | p-value |
| **Sex: W (ref:M)** | 0,62 (0,19 - 2,08) | 0,437 | 0,75 (0,63 - 0,90) | 0,002 | 0,61 (0,51 - 0,73) | <0,001 | 0,86 (0,80 - 0,92) | <0,001 |
| **Age groups: AG1 (ref: AG-R)** | 0,24 (0,05 - 1,06) | 0,059 | 0,64 (0,45 - 0,91) | 0,014 | 0,78 (0,69 - 0,89) | <0,001 | 0,59 (0,52 - 0,68) | <0,001 |
| **AG2 (ref: AG-R)** | 0,72 (0,28 - 1,83) | 0,487 | 0,42 (0,34 - 0,53) | <0,001 | | | 0,36 (0,30 - 0,43) | <0,001 |
| **AG3 (ref: AG-R)** | NA | NA | 0,41 (0,24 - 0,72) | 0,002 | | | 0,25 (0,21 - 0,30) | <0,001 |
| **AG4 (ref: AG-R)** | | | 0,29 (0,16 - 0,55) | <0,001 | | | | |
| **Half-year: (ref: 1. Half year)** | 1,47 (0,18 - 12,32) | 0,719 | 1,20 (0,67 - 2,14) | 0,549 | 0,78 (0,55 - 1,11) | 0,170 | 1,06 (0,87 - 1,29) | 0,543 |
| **Polypharmacy: 2-5 (ref: 0-1)** | 9,00 (1,72 - 47,02) | 0,009 | 2,19 (1,30 - 3,69) | 0,003 | 1,27 (0,83 - 1,94) | 0,263 | 1,27 (1,10 - 1,47) | <0,001 |
| **Polypharmacy: 6-10 (ref: 0-1)** | 8,52 (1,16 - 62,38) | 0,035 | 5,02 (3,21 - 7,83) | <0,001 | 2,08 (1,47 - 2,95) | <0,001 | 1,75 (1,45 - 2,10) | <0,001 |
| **Polypharmacy: ≥11 (ref: 0-1)** | NA | NA | 4,86 (2,07 - 11,42) | <0,001 | 4,09 (3,14 - 5,34) | <0,001 | 1,92 (1,60 - 2,30) | <0,001 |
| **Length of hospital stay** | 1,03 (1,01 - 1,04) | <0,001 | 1,02 (1,01 - 1,02) | <0,001 | 1,01 (1,01 - 1,01) | <0,001 | 1,01 (1,01 - 1,01) | <0,001 |
| **Patients surviving index COVID hospital stay: Hospitalization due to any reason (with competing risk death)** | | | | | | | | |
| | Age 19-40 years | | Age 41-64 years | | Age 65-74 years | | Age ≥75 years | |
| | HR (CI) | p-value | HR (CI) | p-value | HR (CI) | p-value | HR (CI) | p-value |
| **Sex: W (ref:M)** | 1,75 (1,40 - 2,20) | <0,001 | 0,86 (0,79 – 0,95) | 0,002 | 0,80 (0,73 – 0,88) | <0,001 | 0,88 (0,82 – 0,93) | <0,001 |
| **Age groups: AG1 (ref: AG-R)** | 1,01 (0,75 – 1,35) | 0,97 | 0,90 (0,80 – 1,02) | 0,092 | 0,91 (0,83 – 1,00) | 0,047 | 1,18 (1,04 – 1,35) | 0,012 |
| **AG2 (ref: AG-R)** | 1,14 (0,84 – 1,55) | 0,39 | 0,83 (0,73 – 0,94) | 0,004 | | | 1,25 (1,10 – 1,41) | <0,001 |
| **AG3 (ref: AG-R)** | 1,06 (0,77 – 1,47) | 0,72 | 0,83 (0,72 – 0,96) | 0,014 | | | 1,32 (1,17 – 1,49) | <0,001 |
| **AG4 (ref: AG-R)** | | | 0,68 (0,56 – 0,83) | <0,001 | | | | |
| **Half-year: (ref: 1. Half year)** | 1,02 (0,75 – 1,39) | 0,9 | 1,05 (0,92 – 1,20) | 0,49 | 1,02 (0,87 – 1,19) | 0,81 | 1,01 (0,91 -1,11) | 0,92 |
| **Polypharmacy: 2-5 (ref: 0-1)** | 1,32 (1,04 – 1,68) | 0,024 | 1,43 (1,28 – 1,59) | <0,001 | 1,18 (1,01 – 1,37) | 0,032 | 1,29 (1,14 – 1,45) | <0,001 |
| **Polypharmacy: 6-10 (ref: 0-1)** | 2,85 (1,89 – 4,32) | <0,001 | 1,99 (1,75 – 2,28) | <0,001 | 1,56 (1,33 – 1,83) | <0,001 | 1,83 (1,62 – 2,07) | <0,001 |
| **Polypharmacy: ≥11 (ref: 0-1)** | | | 2,90 (2,12 – 3,97) | <0,001 | 2,03 (1,61 – 2,57) | <0,001 | 2,36 (1,98 – 2,81) | <0,001 |
| **Length of hospital stay** | 1,02 (1,01 – 1,03) | <0,001 | 1,02 (1,01 – 1,02) | <0,001 | 1,01 (1,01-1,01) | <0,001 | 1,01 (1,01 – 1,01) | <0,001 |

Note: details of the age group definitions can be found in Table S2. HR, hazard ratio; CI, confidence interval.

**Figure S1: Kaplan-Meier curves for COVID-hospitalized patients by age group.** Corresponding 95% confidence intervals are given for pharmacy (No: 0 vs. Yes: >1). Curves are shown separately for patients with 0 medication groups (green) and at least one medication group (orange).

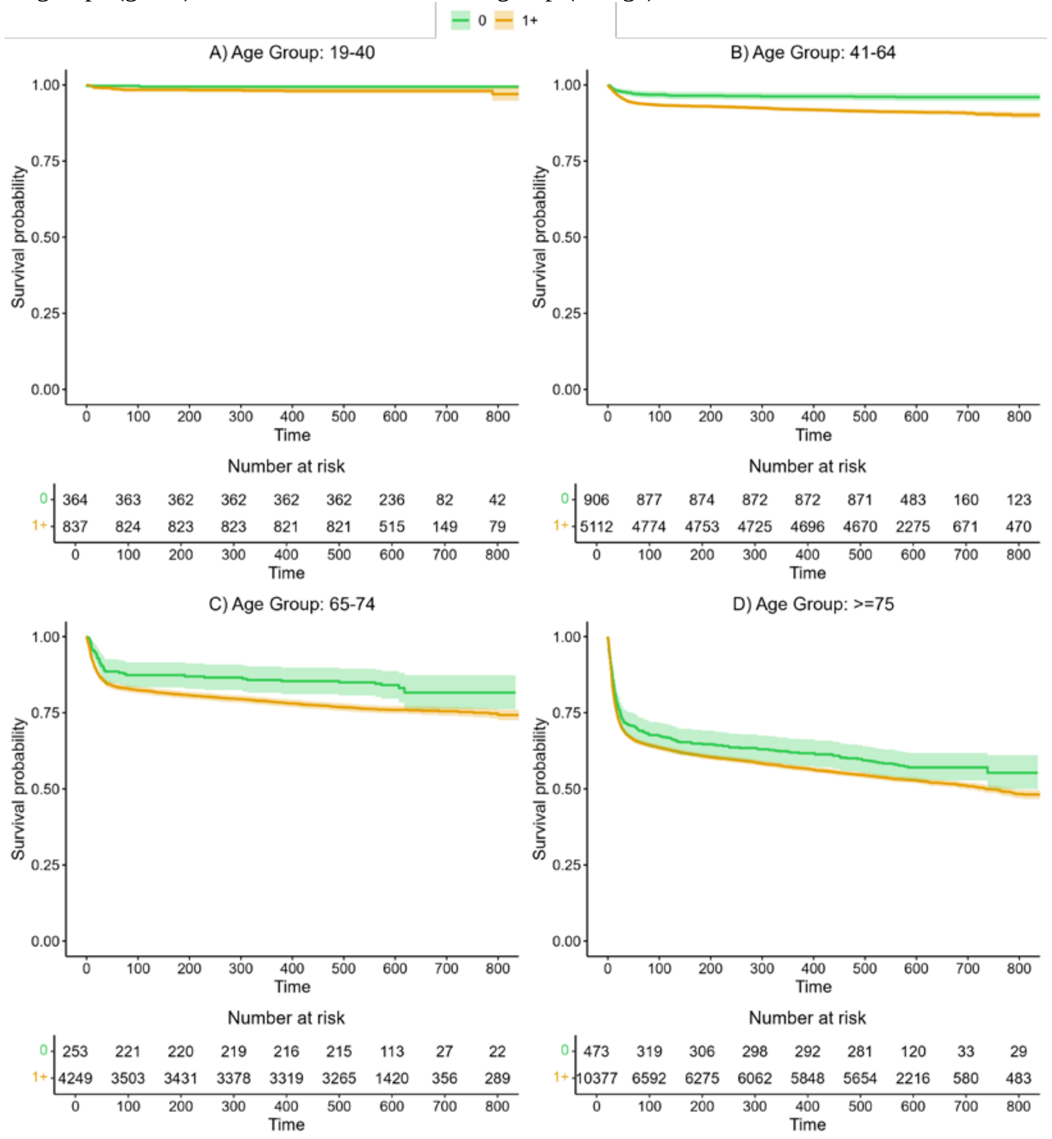

**Table S11: Results of the model for all-cause mortality including medication groups by age group**

| All Patients: all-cause mortality | | | | | | | | |
|---|---|---|---|---|---|---|---|---|
| | Age 19-40 years | | Age 41-64 years | | Age 65-74 years | | Age ≥75 years | |
| | HR (CL) | p-value | HR (CL) | p-value | HR (CL) | p-value | HR (CL) | p-value |
| Sex: W (ref:M) | 0,34 (0,13 - 0,90) | 0,030 | 0,70 (0,55 - 0,89) | 0,004 | 0,53 (0,46 - 0,60) | <0,001 | 0,69 (0,64 - 0,74) | <0,001 |
| Age groups: AG1 (ref: AG-R) | 0,69 (0,13 - 3,79) | 0,672 | 0,69 (0,55 - 0,87) | 0,001 | 0,78 (0,69 - 0,88) | <0,001 | 0,67 (0,62 - 0,73) | <0,001 |
| AG2 (ref: AG-R) | 0,88 (0,24 - 3,18) | 0,841 | 0,43 (0,35 - 0,53) | <0,001 | | | 0,45 (0,41 - 0,50) | <0,001 |
| AG3 (ref: AG-R) | 0,41 (0,08 - 2,23) | 0,302 | 0,35 (0,28 - 0,43) | <0,001 | | | 0,35 (0,31 - 0,39) | <0,001 |
| AG4 (ref: AG-R) | | | 0,20 (0,14 - 0,28) | <0,001 | | | | |
| Half-year: (ref: 1. Half year) | 6,58 (0,30 - 146,02) | 0,234 | 1,07 (0,75 - 1,54) | 0,701 | 0,72 (0,57 - 0,92) | 0,009 | 0,96 (0,84 - 1,09) | 0,521 |
| Polypharmacy: 2-5 (ref: 0-1) | 1,30 (0,31 - 5,49) | 0,719 | 1,13 (0,81 - 1,57) | 0,480 | 1,09 (0,84 - 1,41) | 0,529 | 0,93 (0,80 - 1,09) | 0,354 |
| Polypharmacy: 6-10 (ref: 0-1) | 1,14 (0,06 - 22,69) | 0,934 | 1,07 (0,59 - 1,92) | 0,827 | 1,15 (0,75 - 1,76) | 0,534 | 0,88 (0,73 - 1,06) | 0,180 |
| Polypharmacy: ≥11 (ref: 0-1) | NA | NA | 0,53 (0,22 - 1,27) | 0,153 | 0,91 (0,46 - 1,78) | 0,776 | 0,78 (0,61 - 0,99) | 0,042 |
| Anticoagulants | 2,09 (0,79 - 5,50) | 0,137 | 1,38 (1,16 - 1,65) | <0,001 | 1,22 (1,02 - 1,45) | 0,026 | 1,20 (1,11 - 1,29) | <0,001 |
| Antibiotics, antivirals, antiprotozoals, or anthelmintics | 0,96 (0,39 - 2,34) | 0,929 | 1,24 (1,04 - 1,48) | 0,016 | 1,00 (0,90 - 1,11) | 0,975 | 1,18 (1,07 - 1,29) | <0,001 |
| Insulin and other antidiabetics | 2,68 (0,81 - 8,86) | 0,106 | 1,20 (0,94 - 1,54) | 0,142 | 1,34 (1,24 - 1,45) | <0,001 | 1,16 (1,08 - 1,25) | <0,001 |
| Heart drugs | NA | NA | 0,96 (0,68 - 1,35) | 0,829 | 1,08 (0,86 - 1,36) | 0,483 | 1,09 (1,04 - 1,15) | 0,001 |
| Antihypertensives, incl. diuretics and renin-angiotensin-aldosterone system inhibitors | 1,03 (0,21 - 4,99) | 0,974 | 1,25 (0,96 - 1,62) | 0,097 | 0,93 (0,79 - 1,10) | 0,420 | 0,90 (0,81 - 1,00) | 0,055 |
| Beta-blockers | 1,64 (0,95 - 2,82) | 0,074 | 1,44 (1,11 - 1,88) | 0,006 | 1,10 (1,00 - 1,22) | 0,053 | 1,10 (1,05 - 1,15) | <0,001 |
| Statins, fibrates, incl. proprotein convertase subtilisin/kexin type 9 inhibitors and inclisiran | 1,23 (0,19 - 7,91) | 0,828 | 0,78 (0,63 - 0,96) | 0,019 | 0,77 (0,68 - 0,87) | <0,001 | 0,86 (0,80 - 0,92) | <0,001 |
| Immunosuppressants and | 0,67 (0,15 - 3,01) | 0,606 | 0,81 (0,62 - 1,06) | 0,120 | 0,88 (0,74 - 1,05) | 0,160 | 1,08 (0,98 - 1,20) | 0,108 |
| Systemic steroids | 3,09 (0,55 - 17,47) | 0,202 | 1,22 (1,07 - 1,39) | 0,003 | 1,35 (1,23 - 1,49) | <0,001 | 1,07 (0,98 - 1,18) | 0,132 |
| Chemotherapy | NA | NA | 3,63 (2,69 - 4,90) | <0,001 | 2,13 (1,73 - 2,62) | <0,001 | 1,16 (1,00 - 1,35) | 0,045 |
| Iron supplements, erythropoietic stimulating agents, vitamin B12, folic acid | 1,78 (0,45 - 7,04) | 0,409 | 2,22 (1,81 - 2,73) | <0,001 | 1,45 (1,22 - 1,72) | <0,001 | 1,32 (1,24 - 1,41) | <0,001 |
| Antacids, incl. antihistamines | 1,05 (0,30 - 3,66) | 0,941 | 1,04 (0,93 - 1,16) | 0,520 | 1,06 (0,85 - 1,33) | 0,578 | 1,07 (1,04 - 1,10) | <0,001 |
| Vitamin D and other vitamin supplements | 0,26 (0,14 - 0,50) | <0,001 | 1,25 (0,94 - 1,66) | 0,122 | 1,13 (1,05 - 1,22) | 0,001 | 1,02 (0,97 - 1,06) | 0,501 |
| NSAID and other anti-inflammatory drugs | 0,75 (0,17 - 3,28) | 0,701 | 0,70 (0,56 - 0,87) | 0,001 | 0,65 (0,60 - 0,71) | <0,001 | 0,76 (0,73 - 0,79) | <0,001 |
| Gout medications | NA | NA | 0,93 (0,55 - 1,57) | 0,778 | 1,31 (1,03 - 1,68) | 0,031 | 1,11 (0,98 - 1,25) | 0,103 |
| Antiepileptics | 3,71 (0,58 - 23,57) | 0,165 | 1,71 (1,35 - 2,15) | <0,001 | 1,53 (1,31 - 1,78) | <0,001 | 1,13 (1,08 - 1,19) | <0,001 |
| Antipsychotics | 2,36 (0,85 - 6,57) | 0,099 | 1,51 (1,43 - 1,60) | <0,001 | 1,47 (1,23 - 1,75) | <0,001 | 1,43 (1,39 - 1,48) | <0,001 |
| Inhaled anti-obstructive drugs | 1,50 (0,82 - 2,75) | 0,184 | 1,21 (0,89 - 1,65) | 0,229 | 1,24 (1,10 - 1,40) | <0,001 | 1,08 (0,98 - 1,18) | 0,105 |
| Inhaled steroids | NA | NA | 0,88 (0,55 - 1,41) | 0,608 | 0,99 (0,75 - 1,30) | 0,928 | 0,88 (0,74 - 1,04) | 0,119 |
| Systemic antihistamines | 1,83 (1,26 - 2,67) | 0,002 | 1,18 (0,89 - 1,56) | 0,244 | 0,83 (0,67 - 1,03) | 0,086 | 0,96 (0,86 - 1,06) | 0,386 |

Note: details of the age group definitions can be found in Table S2. A graphical summary of the medication group effects is presented in Figure 3. HR, hazard ratio; CI, confidence interval; NA, “not available” variable could not be included in the model due to low event-rate.

**Table S12: Results of the model for all-cause mortality for the subgroup of patients surviving the hospital stay including medication groups by age group**

| Patients surviving hospital stay: all-cause mortality after hospital stay | | | | | | | | |
|---|---|---|---|---|---|---|---|---|
| | Age 19-40 years | | Age 41-64 years | | Age 65-74 years | | Age ≥75 years | |
| | HR (CI) | p-value | HR (CI) | p-value | HR (CI) | p-value | HR (CI) | p-value |
| Sex: W (ref:M) | 0,62 (0,09 - 4,04) | 0,617 | 0,66 (0,55 - 0,80) | <0,001 | 0,57 (0,47 - 0,68) | <0,001 | 0,81 (0,77 - 0,86) | <0,001 |
| Age groups: AG1 (ref: AG-R) | 1,16 (0,98 - 1,37) | 0,092 | 0,66 (0,48 - 0,91) | 0,010 | 0,86 (0,74 - 1,00) | 0,055 | 0,64 (0,56 - 0,74) | <0,001 |
| AG2 (ref: AG-R) | | | 0,42 (0,33 - 0,54) | <0,001 | | | 0,40 (0,33 - 0,48) | <0,001 |
| AG3 (ref: AG-R) | | | 0,40 (0,23 - 0,71) | 0,002 | | | 0,30 (0,26 - 0,35) | <0,001 |
| AG4 (ref: AG-R) | | | 0,27 (0,15 - 0,49) | <0,001 | | | | |
| Half-year: (ref: 1. Half year) | 1,59 (0,12 - 21,35) | 0,726 | 1,18 (0,67 - 2,08) | 0,567 | 0,75 (0,56 - 1,01) | 0,055 | 1,08 (0,89 - 1,31) | 0,460 |
| Polypharmacy: 2-5 (ref: 0-1) | 3,11 (0,24 - 40,49) | 0,387 | 1,31 (0,65 - 2,62) | 0,448 | 1,14 (0,71 - 1,81) | 0,594 | 0,93 (0,77 - 1,14) | 0,510 |
| Polypharmacy: 6-10 (ref: 0-1) | 0,19 (0,01 - 9,09) | 0,397 | 1,38 (0,48 - 3,98) | 0,546 | 1,33 (0,81 - 2,20) | 0,257 | 0,90 (0,64 - 1,25) | 0,522 |
| Polypharmacy: ≥11 (ref: 0-1) | NA | NA | 0,44 (0,09 - 2,27) | 0,328 | 1,56 (0,84 - 2,90) | 0,160 | 0,65 (0,45 - 0,93) | 0,020 |
| Length of hospital stay | 1,03 (1,00 - 1,06) | 0,022 | 1,01 (1,01 - 1,02) | <0,001 | 1,01 (1,01 - 1,01) | <0,001 | 1,01 (1,01 - 1,01) | <0,001 |
| Anticoagulants | 3,66 (1,36 - 9,89) | 0,010 | 1,78 (1,39 - 2,29) | <0,001 | 1,26 (0,97 - 1,63) | 0,089 | 1,31 (1,17 - 1,46) | <0,001 |
| Antibiotics, antivirals, antiprotozoals, or anthelmintics | 2,21 (0,74 - 6,60) | 0,154 | 1,53 (1,16 - 2,03) | 0,003 | 0,90 (0,72 - 1,13) | 0,360 | 1,28 (1,12 - 1,46) | <0,001 |
| Insulin and other antidiabetics | NA | NA | 0,89 (0,63 - 1,24) | 0,491 | 1,32 (1,08 - 1,61) | 0,006 | 1,14 (1,01 - 1,29) | 0,036 |
| Heart drugs | NA | NA | 0,89 (0,63 - 1,26) | 0,507 | 1,24 (0,86 - 1,78) | 0,245 | 1,19 (1,11 - 1,28) | <0,001 |
| Antihypertensives, incl. diuretics and renin-angiotensin-aldosterone system inhibitors | 2,01 (0,75 - 5,43) | 0,168 | 1,17 (0,94 - 1,45) | 0,164 | 0,92 (0,69 - 1,22) | 0,568 | 0,88 (0,72 - 1,08) | 0,224 |
| Beta-blockers | 7,01 (2,86 - 17,17) | <0,001 | 1,42 (0,96 - 2,10) | 0,082 | 1,03 (0,88 - 1,21) | 0,703 | 1,05 (0,95 - 1,16) | 0,372 |
| Statins, fibrates, incl. proprotein convertase subtilisin/kexin type 9 inhibitors and inclisiran | 1,31 (0,10 - 17,50) | 0,837 | 0,60 (0,40 - 0,92) | 0,020 | 0,66 (0,56 - 0,78) | <0,001 | 0,86 (0,80 - 0,92) | <0,001 |
| Immunosuppressants and immunomodulators | NA | NA | 0,60 (0,36 - 0,98) | 0,043 | 0,79 (0,52 - 1,19) | 0,256 | 0,98 (0,69 - 1,37) | 0,890 |
| Systemic steroids | NA | NA | 1,49 (0,94 - 2,38) | 0,093 | 1,47 (1,10 - 1,98) | 0,010 | 1,02 (0,84 - 1,24) | 0,829 |
| Chemotherapy | NA | NA | 4,15 (2,62 - 6,59) | <0,001 | 3,08 (2,17 - 4,35) | <0,001 | 1,44 (1,19 - 1,72) | <0,001 |
| Iron supplements, erythropoietic stimulating agents, | 2,74 (0,43 - 17,53) | 0,288 | 2,64 (1,91 - 3,65) | <0,001 | 1,86 (1,43 - 2,42) | <0,001 | 1,67 (1,49 - 1,87) | <0,001 |
| Antacids, incl. antihistamines | 0,61 (0,26 - 1,45) | 0,263 | 0,90 (0,67 - 1,20) | 0,476 | 0,87 (0,63 - 1,20) | 0,400 | 0,95 (0,87 - 1,04) | 0,300 |
| Vitamin D and other vitamin supplements | 0,28 (0,04 - 1,71) | 0,168 | 1,28 (1,05 - 1,56) | 0,014 | 1,03 (0,85 - 1,26) | 0,732 | 1,00 (0,91 - 1,11) | 0,992 |
| NSAID and other anti-inflammatory drugs | 0,68 (0,08 - 6,01) | 0,729 | 0,65 (0,53 - 0,80) | <0,001 | 0,72 (0,61 - 0,86) | <0,001 | 0,70 (0,64 - 0,76) | <0,001 |
| Gout medications | NA | NA | 0,51 (0,23 - 1,13) | 0,099 | 0,99 (0,68 - 1,45) | 0,978 | 1,02 (0,87 - 1,18) | 0,841 |
| Antiepileptics | 3,99 (0,37 - 43,09) | 0,255 | 1,92 (1,44 - 2,57) | <0,001 | 1,37 (1,16 - 1,62) | <0,001 | 1,17 (1,03 - 1,33) | 0,016 |
| Antipsychotics | 2,00 (0,69 - 5,81) | 0,203 | 1,80 (1,54 - 2,10) | <0,001 | 1,63 (1,38 - 1,93) | <0,001 | 1,59 (1,47 - 1,71) | <0,001 |
| Inhaled anti-obstructive drugs | 1,95 (0,61 - 6,17) | 0,258 | 1,15 (0,68 - 1,97) | 0,599 | 1,25 (1,00 - 1,57) | 0,054 | 1,21 (1,10 - 1,34) | <0,001 |
| Inhaled steroids | NA | NA | 0,79 (0,43 - 1,46) | 0,456 | 0,96 (0,63 - 1,45) | 0,834 | 0,84 (0,61 - 1,16) | 0,285 |
| Systemic antihistamines | 1,35 (0,15 - 11,72) | 0,787 | 1,53 (0,99 - 2,38) | 0,058 | 0,86 (0,61 - 1,23) | 0,421 | 0,90 (0,82 - 0,99) | 0,037 |

HR, hazard ratio; CI, confidence interval, NA, “not available” variable could not be included in the model due to low event-rate.

**Figure S2: Hazard ratios and 95% confidence intervals for medication groups for the outcome all-cause mortality for the subgroup of patients surviving the COVID-hospital stay.** For details on hazard ratios, confidence limits, and p-values, see Table S12. A hazard ratio >1 indicates a larger risk for all-cause death for patients receiving a drug in the corresponding medication group.

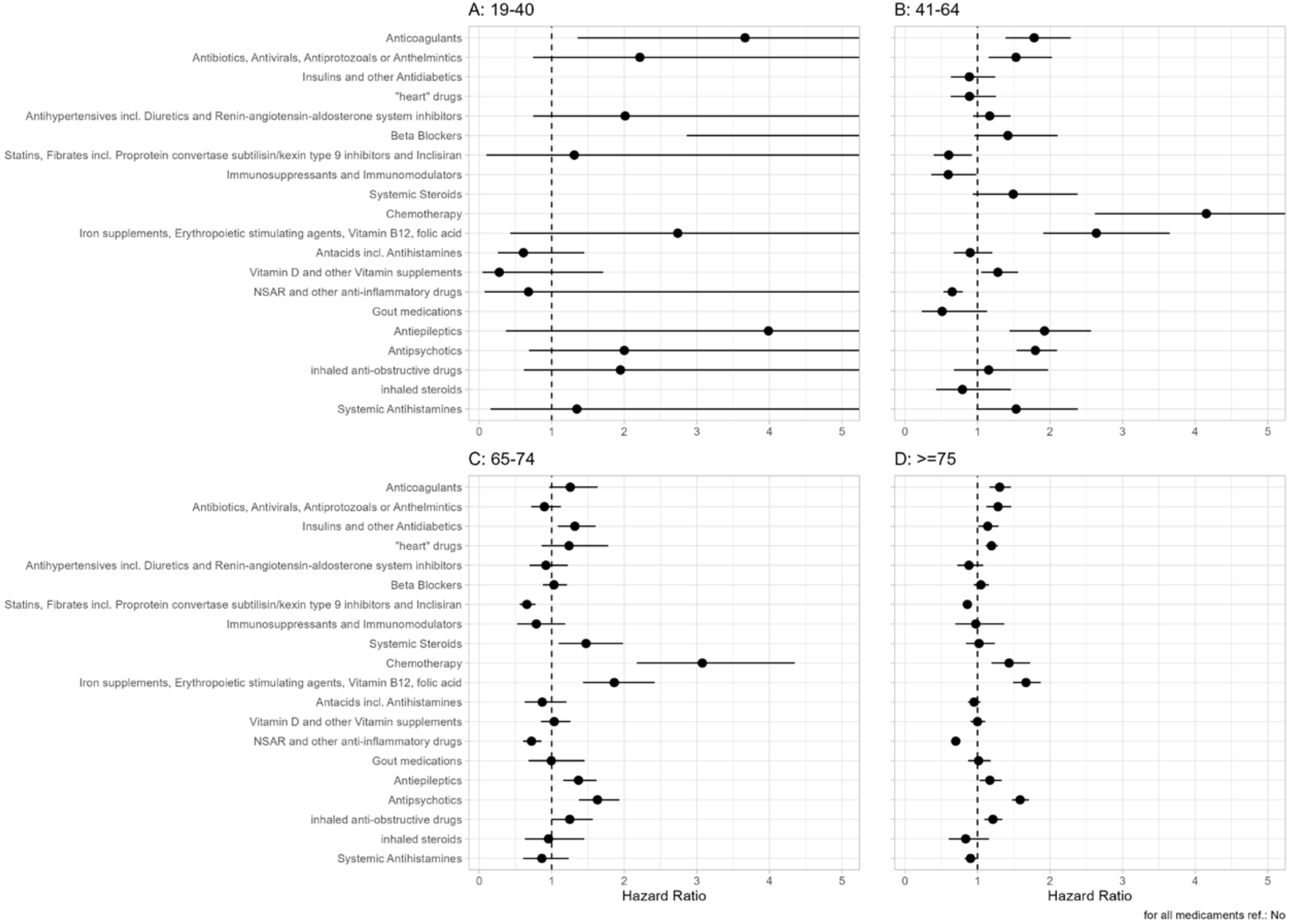

**Table S13: Results of the model for hospitalization due to any reason for the subgroup of patients surviving the index COVID hospital stay including medication groups by age group**

| Patients surviving index COVID-19 hospital stay: Hospitalization due to any reason (with competing risk death) | | | | | | | | |
|---|---|---|---|---|---|---|---|---|
| | **Age 19-40 years** | | **Age 41-64 years** | | **Age 65-74 years** | | **Age ≥75 years** | |
| | **HR (CI)** | **p-value** | **HR (CI)** | **p-value** | **HR (CI)** | **p-value** | **HR (CI)** | **p-value** |
| **Sex: W (ref:M)** | 1,65 (1,30 - 2,09) | <0,001 | 0,83 (0,76 - 0,91) | <0,001 | 0,81 (0,73 - 0,89) | <0,001 | 0,88 (0,82 - 0,94) | <0,001 |
| **Age groups: AG1 (ref: AG-R)** | 1,09 (0,81 - 1,46) | 0,57 | 0,89 (0,79 - 1,01) | 0,071 | 0,94 (0,85 - 1,03) | 0,19 | 1,19 (1,05 - 1,36) | 0,009 |
| **AG2 (ref: AG-R)** | 1,21 (0,88 - 1,66) | 0,24 | 0,83 (0,73 - 0,95) | 0,006 | | | 1,26 (1,11 - 1,43) | <0,001 |
| **AG3 (ref: AG-R)** | 1,12 (0,80 - 1,57) | 0,51 | 0,83 (0,72 - 0,97) | 0,016 | | | 1,36 (1,20 - 1,54) | <0,001 |
| **AG4 (ref: AG-R)** | | | 0,70 (0,57 - 0,85) | <0,001 | | | | |
| **Half-year: (ref: 1. Half year)** | 1,01 (0,74 - 1,39) | 0,95 | 1,00 (0,88 - 1,15) | 0,95 | 1,02 (0,87 - 1,18) | 0,84 | 1,00 (0,90 - 1,11) | 0,95 |
| **Polypharmacy: 2-5 (ref: 0-1)** | 0,95 (0,64 - 1,40) | 0,79 | 1,04 (0,88 - 1,23) | 0,66 | 0,98 (0,81 - 1,20) | 0,87 | 0,96 (0,82 - 1,11) | 0,55 |
| **Polypharmacy: 6-10 (ref: 0-1)** | 1,18 (0,46 - 3,02) | 0,73 | 0,84 (0,61 - 1,18) | 0,32 | 1,04 (0,75 - 1,44) | 0,82 | 0,96 (0,76 - 1,21) | 0,73 |
| **Polypharmacy: ≥11 (ref: 0-1)** | | | 0,58 (0,32 - 1,04) | 0,069 | 0,93 (0,55 - 1,58) | 0,8 | 0,84 (0,59 - 1,19) | 0,33 |
| **Length of hospital stay** | 1,02 (1,01 - 1,03) | <0,001 | 1,01 (1,01 - 1,02) | <0,001 | 1,01 (1,01 - 1,01) | <0,001 | 1,01 (1,01 - 1,01) | <0,001 |
| **Anticoagulants** | 1,49 (0,94 - 2,38) | 0,09 | 1,37 (1,21 - 1,54) | <0,001 | 1,20 (1,07 - 1,34) | 0,001 | 1,23 (1,14 - 1,33) | <0,001 |
| **Antibiotics, antivirals, antiprotozoals, or** | 1,05 (0,81 - 1,36) | 0,71 | 1,15 (1,04 - 1,28) | 0,009 | 1,08 (0,98 - 1,2) | 0,13 | 1,12 (1,05 - 1,20) | 0,001 |
| **Insulin and other antidiabetics** | 2,26 (1,29 - 3,97) | 0,0044 | 1,06 (0,92 - 1,22) | 0,43 | 1,02 (0,90 - 1,15) | 0,75 | 1,09 (1,01 - 1,19) | 0,031 |
| **Heart drugs** | 1,15 (0,53 - 2,49) | 0,72 | 1,21 (1,00 - 1,47) | 0,052 | 1,18 (1,01 - 1,37) | 0,037 | 1,18 (1,08 - 1,29) | <0,001 |
| **Antihypertensives, incl. diuretics and renin-angiotensin-aldosterone system inhibitors** | 1,67 (1,06 - 2,62) | 0,026 | 1,12 (1,00 - 1,26) | 0,048 | 1,04 (0,93 - 1,17) | 0,5 | 1,09 (1,00 - 1,19) | 0,039 |
| **Beta-blockers** | 2,17 (1,24 - 3,8) | 0,007 | 1,27 (1,1 - 1,45) | <0,001 | 1,01 (0,90 - 1,14) | 0,82 | 1,05 (0,97 - 1,13) | 0,2 |
| **Statins, fibrates, incl. proprotein convertase subtilisin/kexin type 9 inhibitors and inclisiran** | 0,76 (0,36 - 1,59) | 0,46 | 0,92 (0,82 - 1,04) | 0,18 | 1,04 (0,93 - 1,16) | 0,52 | 1,00 (0,92 - 1,07) | 0,91 |
| **Immunosuppressants and immunomodulators** | 0,53 (0,22 - 1,28) | 0,16 | 1,00 (0,81 - 1,24) | 0,98 | 0,99 (0,80 - 1,21) | 0,89 | 1,23 (1,01 - 1,51) | 0,042 |
| **Systemic steroids** | 1,29 (0,82 - 2,03) | 0,26 | 1,39 (1,21 - 1,60) | <0,001 | 1,54 (1,36 - 1,76) | <0,001 | 1,17 (1,07 - 1,29) | <0,001 |
| **Chemotherapy** | 1,95 (0,79 - 4,83) | 0,15 | 1,64 (1,22 - 2,2) | <0,001 | 1,42 (1,13 - 1,79) | 0,003 | 1,26 (1,07 - 1,48) | 0,006 |
| **Iron supplements, erythropoietic stimulating agents, vitamin B12, folic acid** | 1,39 (0,88 - 2,19) | 0,16 | 1,45 (1,19 - 1,77) | <0,001 | 1,21 (1,02 - 1,44) | 0,033 | 1,20 (1,08 - 1,32) | <0,001 |
| **Antacids, incl. antihistamines** | 1,02 (0,66 - 1,59) | 0,92 | 0,94 (0,82 - 1,07) | 0,31 | 0,86 (0,76 - 0,98) | 0,019 | 1,00 (0,92 - 1,08) | 0,92 |
| **Vitamin D and other vitamin supplements** | 0,82 (0,53 - 1,26) | 0,36 | 1,23 (1,07 - 1,41) | 0,003 | 0,91 (0,79 - 1,04) | 0,18 | 0,97 (0,89 - 1,05) | 0,47 |
| **NSAID and other anti-inflammatory drugs** | 0,93 (0,69 - 1,27) | 0,66 | 1,04 (0,93 - 1,15) | 0,49 | 0,91 (0,81 - 1,01) | 0,082 | 1,12 (1,04 - 1,20) | 0,004 |
| **Gout medications** | 0,77 (0,19 - 3,03) | 0,7 | 1,03 (0,8 - 1,31) | 0,84 | 1,05 (0,85 - 1,29) | 0,65 | 1,07 (0,95 - 1,20) | 0,25 |
| **Antiepileptics** | 1,03 (0,65 - 1,63) | 0,9 | 1,38 (1,19 - 1,61) | <0,001 | 1,35 (1,17 - 1,54) | <0,001 | 1,16 (1,07 - 1,27) | <0,001 |
| **Antipsychotics** | 1,62 (1,16 - 2,26) | 0,005 | 1,25 (1,11 - 1,40) | <0,001 | 1,15 (1,03 - 1,29) | 0,011 | 1,14 (1,05 - 1,22) | <0,001 |
| **Inhaled anti-obstructive drugs** | 1,09 (0,77 - 1,54) | 0,64 | 1,09 (0,96 - 1,23) | 0,17 | 1,11 (0,98 - 1,24) | 0,093 | 1,13 (1,04 - 1,23) | 0,005 |
| **Inhaled steroids** | 1,18 (0,62 - 2,23) | 0,62 | 0,98 (0,78 - 1,23) | 0,83 | 0,95 (0,76 - 1,20) | 0,68 | 0,82 (0,68 - 1,00) | 0,053 |
| **Systemic antihistamines** | 0,69 (0,39 - 1,22) | 0,2 | 0,95 (0,79 - 1,15) | 0,61 | 0,97 (0,80 - 1,19) | 0,8 | 1,03 (0,92 - 1,16) | 0,61 |

HR, hazard ratio; CI, confidence interval.

**Figure S3: Hazard ratios and 95% confidence intervals for medication groups for the outcome hospitalization for the subgroup of patients surviving the index COVID-hospital stay.** For details on hazard ratios, confidence limits, and p-values, see Table S13. A hazard ratio >1 indicates a larger risk for all-cause death for patients receiving a drug in the corresponding medication group.

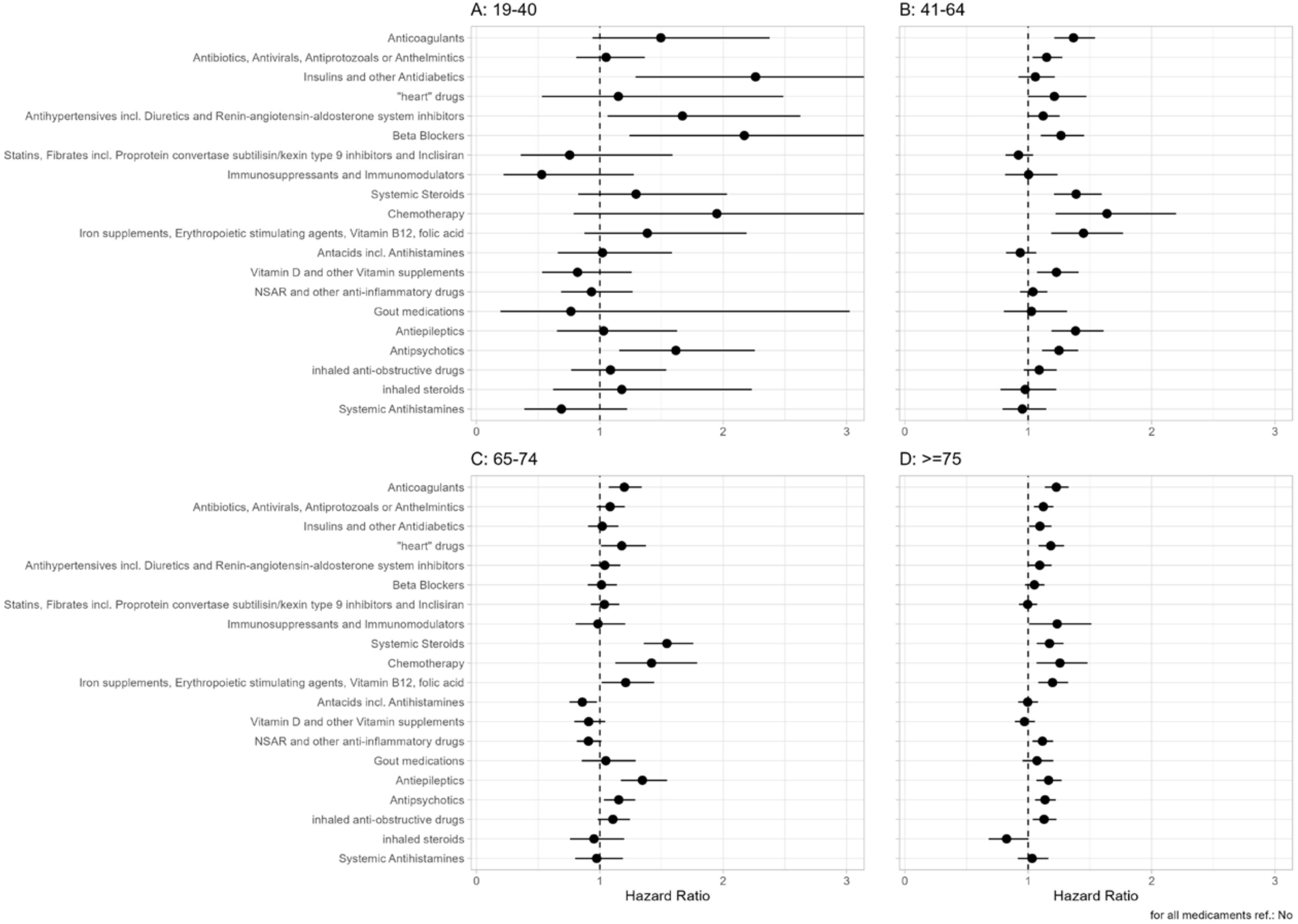

### COVID patients compared to control

The following two **Figures S4** and **S5** evaluate the role of pharmacy in all-cause death in the control group. Note that corresponding plots for the patient cohort can be found in the manuscript (Figure 2) and in Figure S1. **Figures S6** gives Kaplan-Meier curves of medication groups associated with poor survival in the control cohort. Note that corresponding plots for the patient cohort can be found in the manuscript (Figure 2), Figure S1 and Figure 4 (manuscript).

**Figure S6** shows Kaplan-Meier curves of 6 medication groups associated with poor survival in the control cohort. Note that the corresponding plot for the patient cohort can be found in the manuscript (Figure 4).

**Figure S4: Kaplan-Meier curves for the control population by age group.** Corresponding 95% confidence intervals are given for pharmacy groups. Curves are shown separately for patients with drugs in 0-1 (green), 2-5 (orange), 6-10 (blue) and ≥11 (red) medication groups.

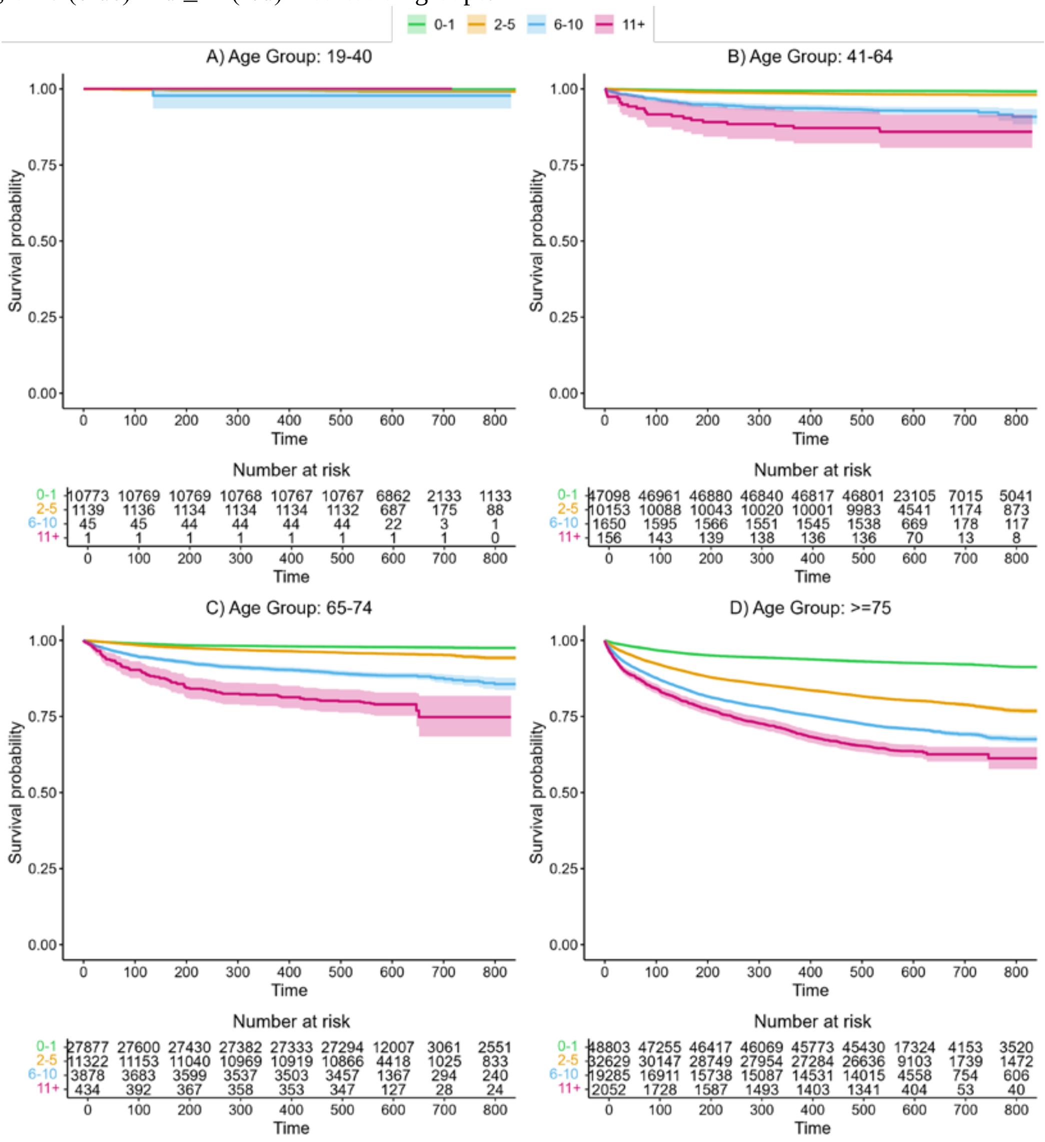

**Figure S5: Kaplan-Meier curves for the control population by age group.** Corresponding 95% confidence intervals are given for pharmacy (No: 0 vs. Yes: >1). Curves are shown separately for patients with drugs in 0 medication groups (green) and at least one medication group (orange).

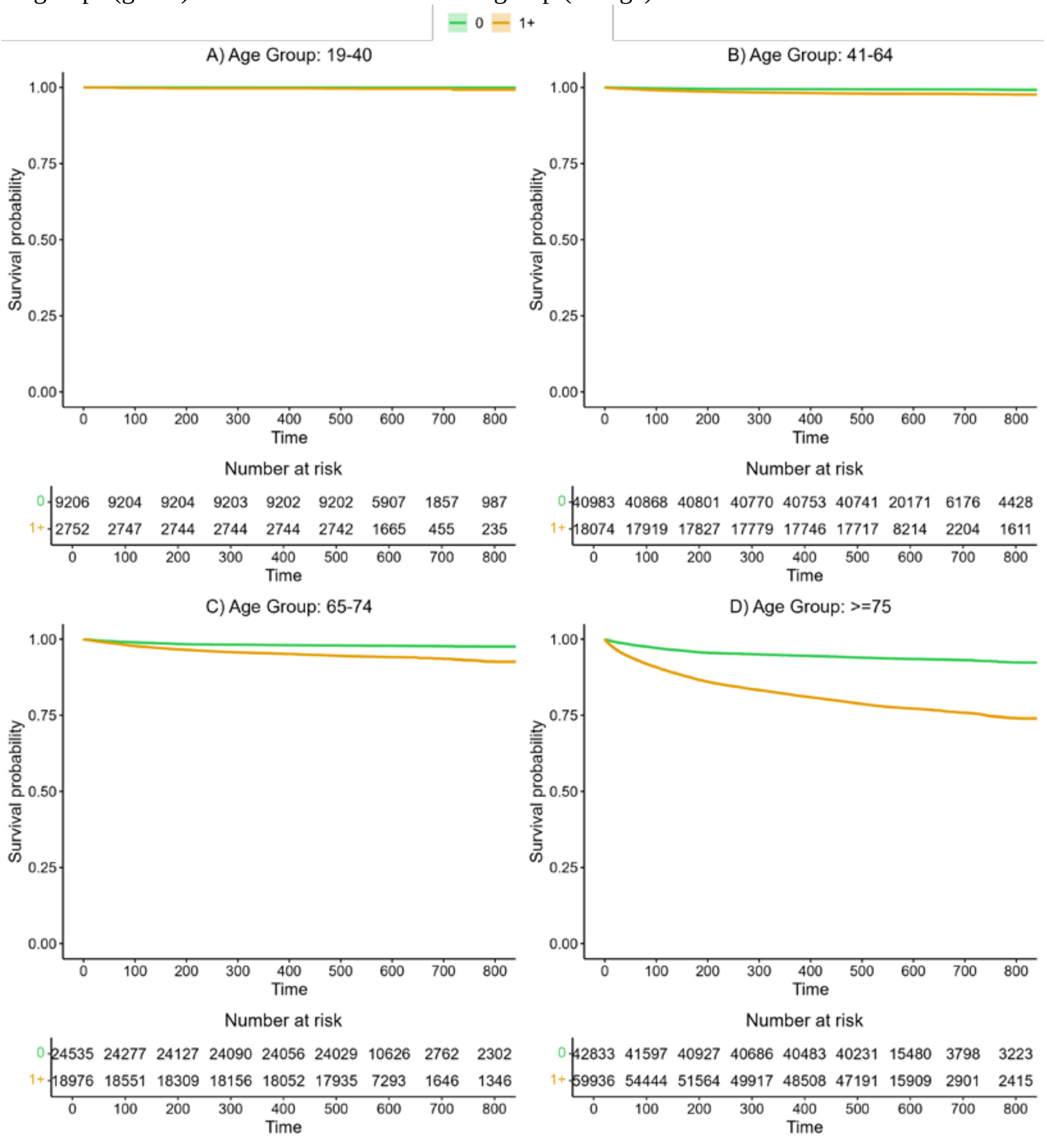

**Figure S6: Kaplan-Meier curves of medication groups associated with poor survival in the control cohort.**

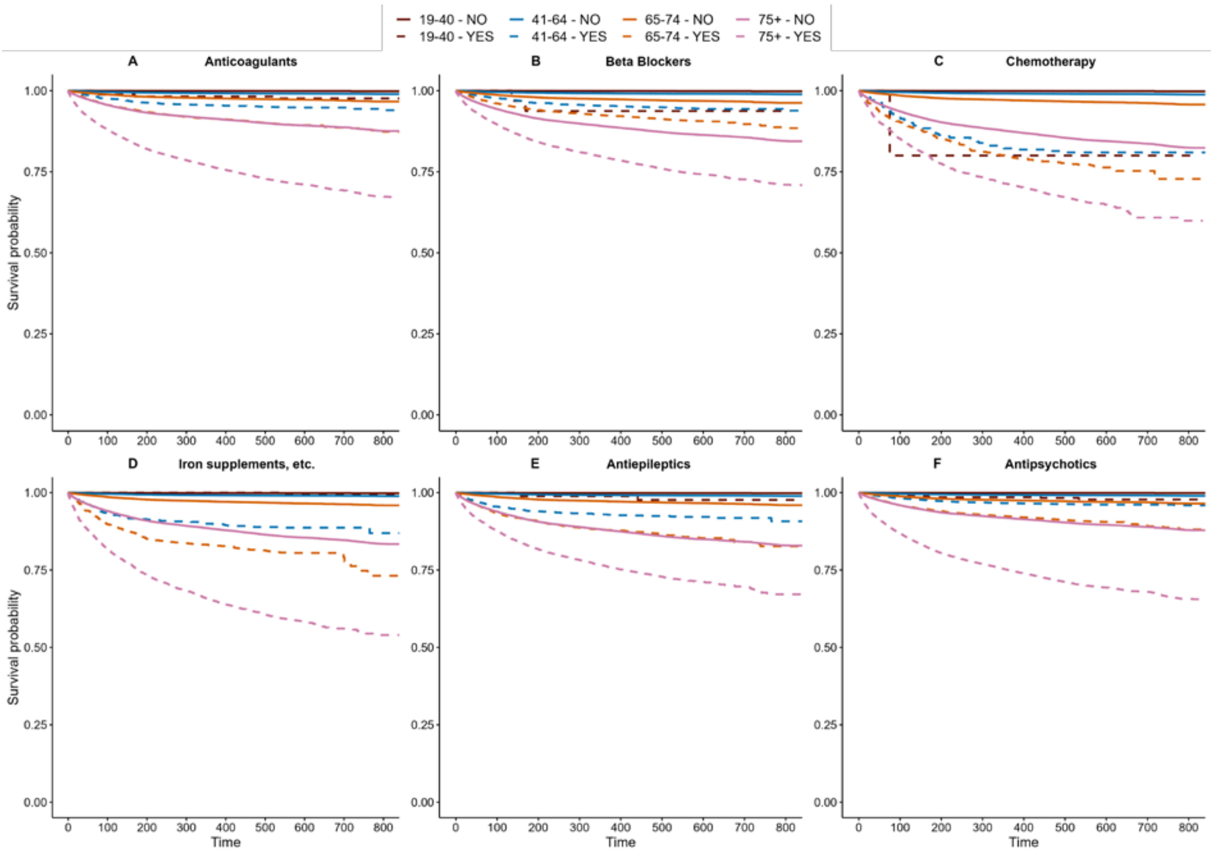

To evaluate the additional group effect of COVID on all-cause mortality, multivariable Cox regression models including sex, age, polypharmacy, half-year, and medicament groups with clustering variable region were calculated for the propensity score-matched patients and controls (propensity score-matching based on medication groups). Due to the too small number of events after propensity score matching, this model could not be evaluated within the 19-40 years age group.

A similar model was calculated for all-cause mortality for the subgroup of patients surviving the hospital stay.

A competing risk model with competing risk death was calculated for the outcome hospitalization. The model accounted for the same confounding factors as the other two models. **Table S14** shows the group effects of the multivariable models separately for age groups.

**Table S14: Group effects (COVID-patients vs. controls) of the multivariable models of the three outcome parameters by age group**

| | **All-cause mortality** | | **All-cause mortality after surviving hospital stay** | | **Hospitalization** | |
|---|---|---|---|---|---|---|
| **Age group** | **HR (CI)** | **p-value** | **HR (CI)** | **p-value** | **HR (CI)** | **p-value** |
| **19-40 years** | NA | NA | NA | NA | 3,66 (2,88 - 4,65) | <0,001 |
| **41-64 years** | 3,24 (2,68 - 3,93) | <0,001 | 1,65 (1,38 - 1,97) | <0,001 | 2,52 (2,35 - 2,71) | <0,001 |
| **65-74 years** | 3,45 (3,04 - 3,92) | <0,001 | 1,55 (1,36 - 1,76) | <0,001 | 1,99 (1,86 - 2,13) | <0,001 |
| **≥75 years** | 2,39 (2,14 - 2,66) | <0,001 | 1,08 (0,99 - 1,17) | 0,078 | 1,58 (1,52 - 1,65) | <0,001 |

HR, hazard ratio; CI, confidence interval; NA, “not available” could not calculated due to low event-rate.

**Figure S7: Kaplan-Meier curves for propensity score-matched COVID patients and controls.** (A,B) All-cause mortality, (C,D) all-cause mortality after COVID-hospital survival, and (E,F) hospitalization.

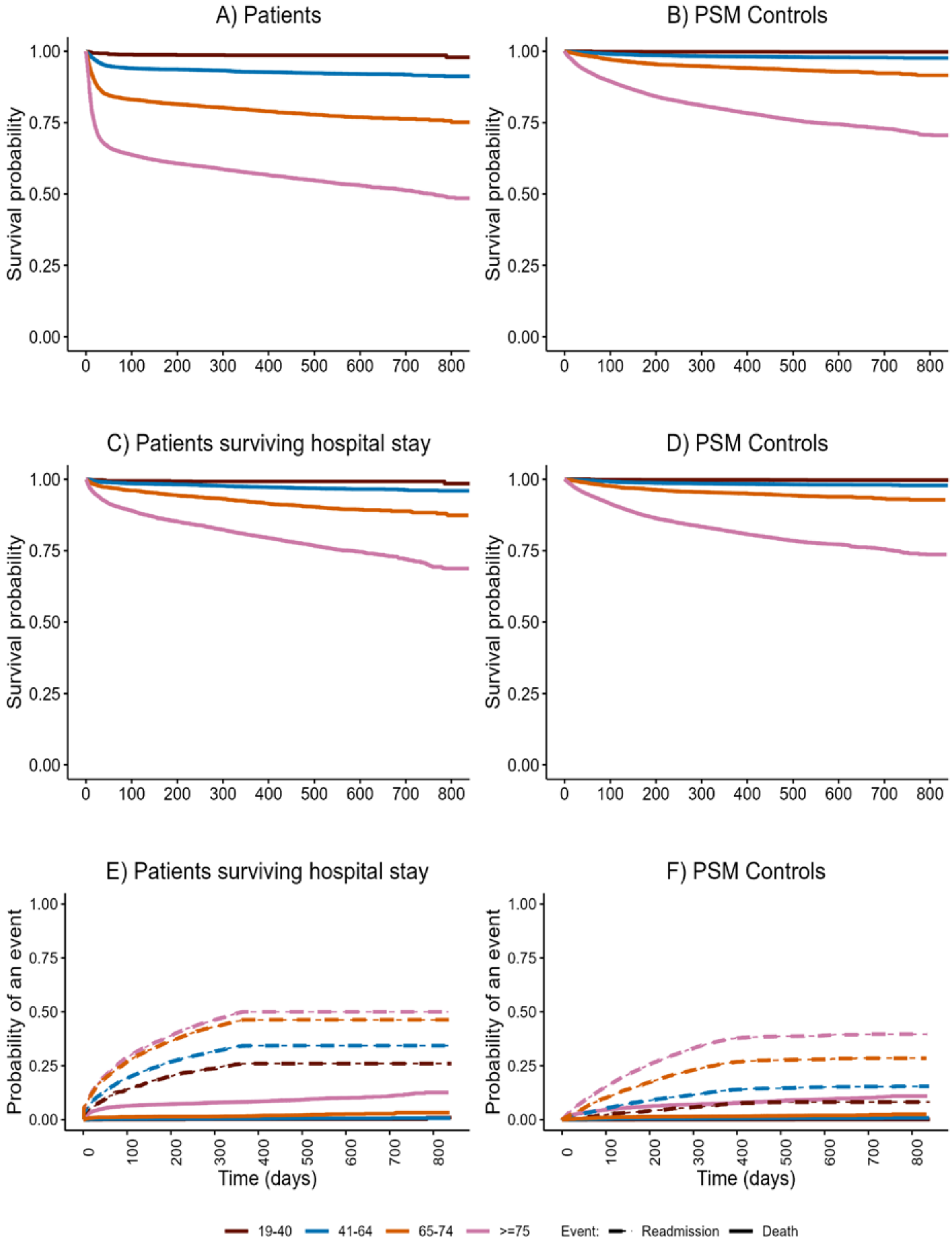
